# Long-term negative divergence in mortality at ages 25-49 years between the United Kingdom and 21 peer countries between 1990 and 2019

**DOI:** 10.1101/2025.05.02.25325864

**Authors:** David A Leon, Dmitry Jdanov, Naomi Medina-Jaudes, Inna Danilova, Vladimir M Shkolnikov

## Abstract

**Background:** The poor performance of the UK in reducing mortality compared to many other high-income countries following the 2008 financial crisis have been extensively studied, with particular attention to deaths of despair at working ages. However, longer-term trends in the differences in working-age mortality between the UK and peer countries have not been systematically investigated.

**Methods:** We compared trends (1990-2019) in age-standardised mortality rates at age 25-49 years in the UK and its constituent parts (England and its 9 standard regions, Wales, Scotland, Northern Ireland) with those of 21 peer countries.

**Findings:** Between 1990 and 2019 the UK went from having relatively low mortality rates at age 25-49 years compared to its peers to having one of the highest. This reflects both the better progress made by many other countries in reducing mortality rates as well as an absolute increase in the UK from 2013. Against the counter-factual that rates in the UK followed the median of the comparator countries (2001-2019) this resulted in 3.1 million excess years of life lost. The divergence in mortality of the UK with its peers was apparent from 1990 and was observed for all constituent parts of the UK and English regions. External cause mortality accounted for much of the divergence in rates between 2001 and 2019 (69% women; 78% men), as did the overlapping categories of drug-related deaths (42%; 28%) and suicides (17%; 20%). Alcohol-related deaths made only a small contribution.

**Interpretation:** The divergence in mortality rates at ages 25-49 years in the UK from peer countries was already apparent from 1990, pre-dating the austerity policies two decades later. Nevertheless, austerity may well have exacerbated this longer-term deterioration in the UKs position. The fact that all areas of the UK showed deterioration relative to peer countries indicates that this is a national problem.

## Introduction

Comparing mortality rates between peer countries can generate public health insights.^1^ Progress in any one country can be placed in the context of what others have already achieved. It can also provide warning signals about adverse trends relative to peers that would be missed by considering national mortality trends in isolation. Using this approach, we have previously found that between the mid-2000s and 2016 working-age adults in England and Wales developed exceptionally high mortality compared to a group of other high-income countries.^2^ The emergence of this hotspot is notable for three reasons. Firstly, this is an age group that historically had lower than average mortality rates in the UK compared to peer countries. Secondly, this age group has been identified as potentially being affected by “deaths of despair” consequent upon austerity in the UK following the 2008 financial crisis.^3,4^ Thirdly, any deaths at this relatively young age have a disproportionate impact on years of life lost compared to deaths at older ages.

This paper explores this phenomenon in unprecedented detail. We extend our original analyses of all-cause mortality in England and Wales^2^ to cover all of the UK up to 2019. We determine whether the UK remains an outlier compared to peers, quantifying the public health impact of this in terms of years of life lost. We then assess the extent to which these negative trends relative to peers have affected all parts of the UK (constituent countries and English regions). Finally, we identify which causes of death have contributed to the UK’s divergence from peer countries, including the “deaths of despair” that have been linked to adverse trends in working age mortality in the USA^5^ and elsewhere.^6^ Our analyses will contribute to understanding the factors underlying these negative trends and through this help identify priorities for action in the UK.

## Methods

Data on all-cause mortality (by single-year and 5-year ages, sex and year) for the UK as a whole and for the 21 comparator high-income countries plus the corresponding population exposure were taken from the Human Mortality Database.^7^ For the most recent years (2022-23), annual rates were estimated using the weekly mortality data from the Short Term Mortality Fluctuations data series of the HMD (STMF@HMD) as described elsewhere.^8^ Mortality and population data used in analyses of all-cause mortality (1990-2019) for England, Wales, and the 9 standard regions were provided by the Office for National Statistics (ONS).^9^ Data for Scotland and Northern Ireland and the corresponding population exposure was taken from the Human Mortality Database. The populations and data sources for the all-cause mortality analysis are listed in Table SM1 of Supplementary Methods.

Data on cause-specific mortality (by 5-year age group, sex, year and cause) for England, Wales and the 9 standard regions of England were provided by the ONS.^9^ Equivalent cause-specific death rates for Northern Ireland and Scotland were calculated using HMD population exposures while cause-specific death counts were provided by the Northern Ireland Statistics Agency and National Records of Scotland. The WHO Mortality Database^10^ was used as the source of data for the 21 comparator countries on cause-specific mortality coded to ICD10. Death counts by cause were missing for some countries and in specific years, as summarised in Supplementary Methods Figure SM1.

Deaths at ages 25-49 years have a distinct cause profile which differs by sex (see Supplementary Results Table S1). Among women in 2019, the three largest ICD10 chapter were malignant neoplasms (36% of total deaths) followed by external causes (21%) and circulatory diseases (13%). Among men the same three ICD10 chapters dominated with external causes being the largest (38%) followed by circulatory diseases (18%) and then all malignant neoplasms (17%). In our cause specific analyses we focus on these three chapters that in 2019 accounted for 70% of all deaths among women and 73% of deaths among men. In addition we also pay attention to three cause sub-groups that make up the “deaths of despair”.^5^ These are deaths from suicide and undetermined intent, alcohol-related causes and drug-related causes. The ICD10 codes used to define each of these three cause aggregates are described in Supplementary Methods Table SM2. Discussion of the validity of cause of death data, with a focus on potential inconsistencies induced by ICD10 updates, is provided in the Supplementary Methods.

We compared mortality in the UK with the median death rates across the 21 comparator high-income countries taking age into account. Further details about the calculation of the comparator medians are provided in Supplementary Methods. We computed sex-specific age-standardized deaths rates (ASDRs) in the age group 25-49 years for selected causes of death and all causes combined using the European standard population from 2013. The confidence intervals for cause-specific ASDRs were computed using Keyfitz’ formula.^11^

To quantify the public health impact of divergence of mortality rates in the UK from the comparator median levels, we calculated excess deaths and years of life lost as observed against the counter-factual of rates in the UK being the same as for the comparator median. Further details are given in the Supplementary Methods.

We restricted our main analyses of all-cause mortality to 1990-2019. For cause-specific analyses we took a narrower period (2001-19). Starting in 2001 avoids the uncertainties inherent in creating consistent cause of death categories across different revisions of the

International Classification of Diseases (ICD)^12^ as all countries from this year onwards provided data coded to ICD10. Choosing 2019 as our final year was dictated by the fact that for 2020 onwards a substantial proportion of our comparator countries have not yet made these data available to the WHO.

To examine geographic patterns of the mortality excess within the UK, we built geographic maps of the ratio of UK ASDRs to the median value for the years 1990, 2000, 2010, and 2019. Data handling, statistical analyses and mapping used Microsoft Excel, Stata-17 and R-studio.^13^

## Results

Trends (1990-2019) in UK mortality rates relative to the comparator median (expressed as rate ratios) are shown in Figure 1 (see Supplementary Figure S1 for a version extended to 2023). The age-group 25-49 stands out as showing the most pronounced change over time. In 1990 UK mortality rates among these young adults were at the level of the comparator median (women) or appreciably below it (men). Since then there was been a progressive deterioration in the UK’s position for this age group. The ratio of the UK to comparator median rates at age 25-49 years became the highest of any age-group for women from 2010 and from 2006 for men. An alternative and more granular visualisation of these age effects for the longer period 1970-2023 is provided in Supplementary Figure S2.

**Figure 1.**
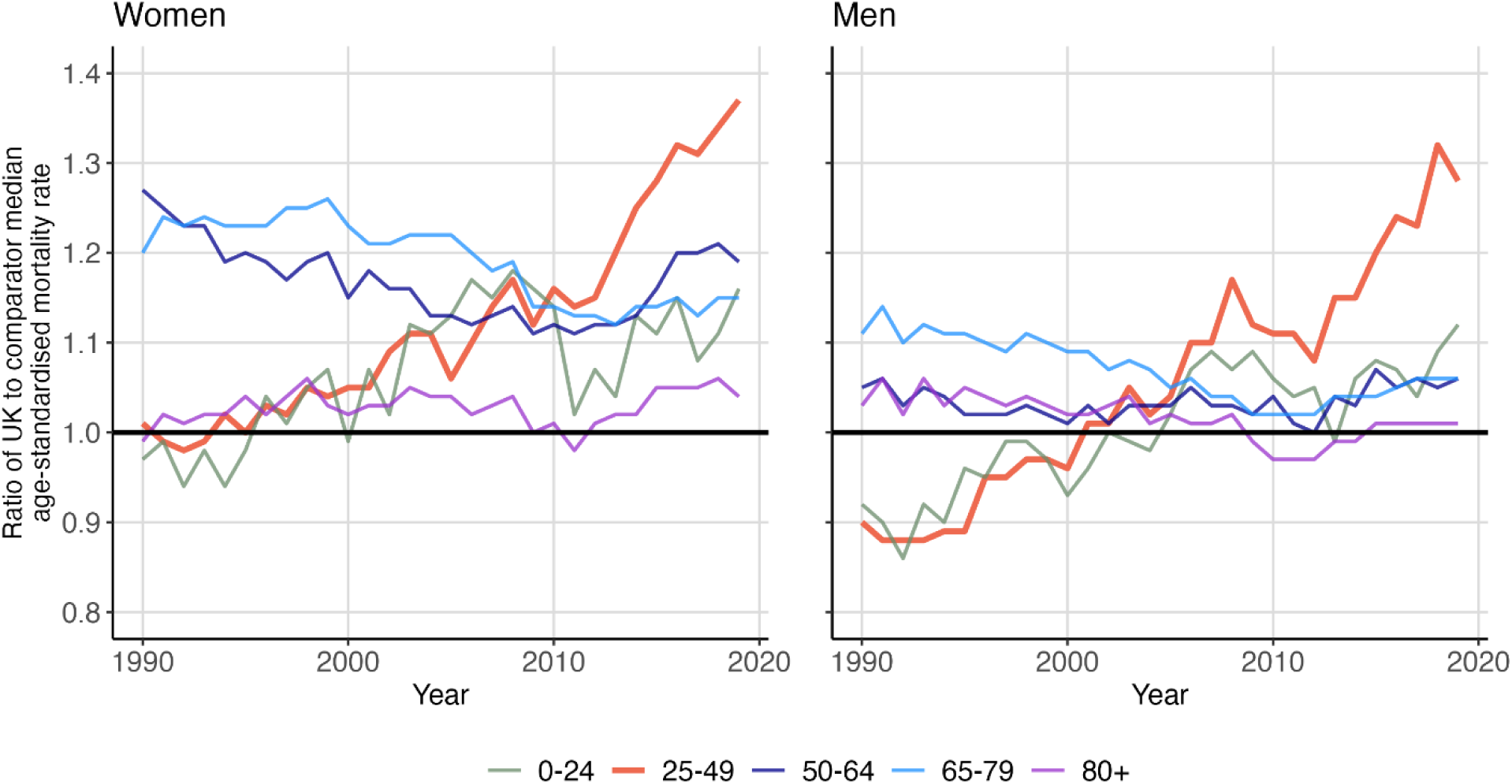
Trends (1990-2019) in ratio of all-cause age-standardised mortality rates in the UK relative to the comparator median of 21 peer countries by age-group and sex.

Trends in mortality rates for the UK and the comparator median at ages 25-49 years are shown superimposed over trends for each of the 21 individual comparator countries in Figure 2 (see also Supplementary Table S2 and Supplementary Figure S3 for the years 1990-2023). This shows the deterioration of the UK compared to the majority of the comparator countries. For males in particular, the UK goes from having one of the lowest rates in 1990 to having one of the highest in 2019. In 1990 mortality rates among men in the UK were ranked 18^th^ out of all 22 countries we consider, moving to 14^th^ in 2000, 6^th^ in 2010 and 2^nd^ (after the USA) in 2019. For women the progress in rank order in the same years was 12^th^, 10^th^, 6^th^ and 2^nd^.

**Figure 2.**
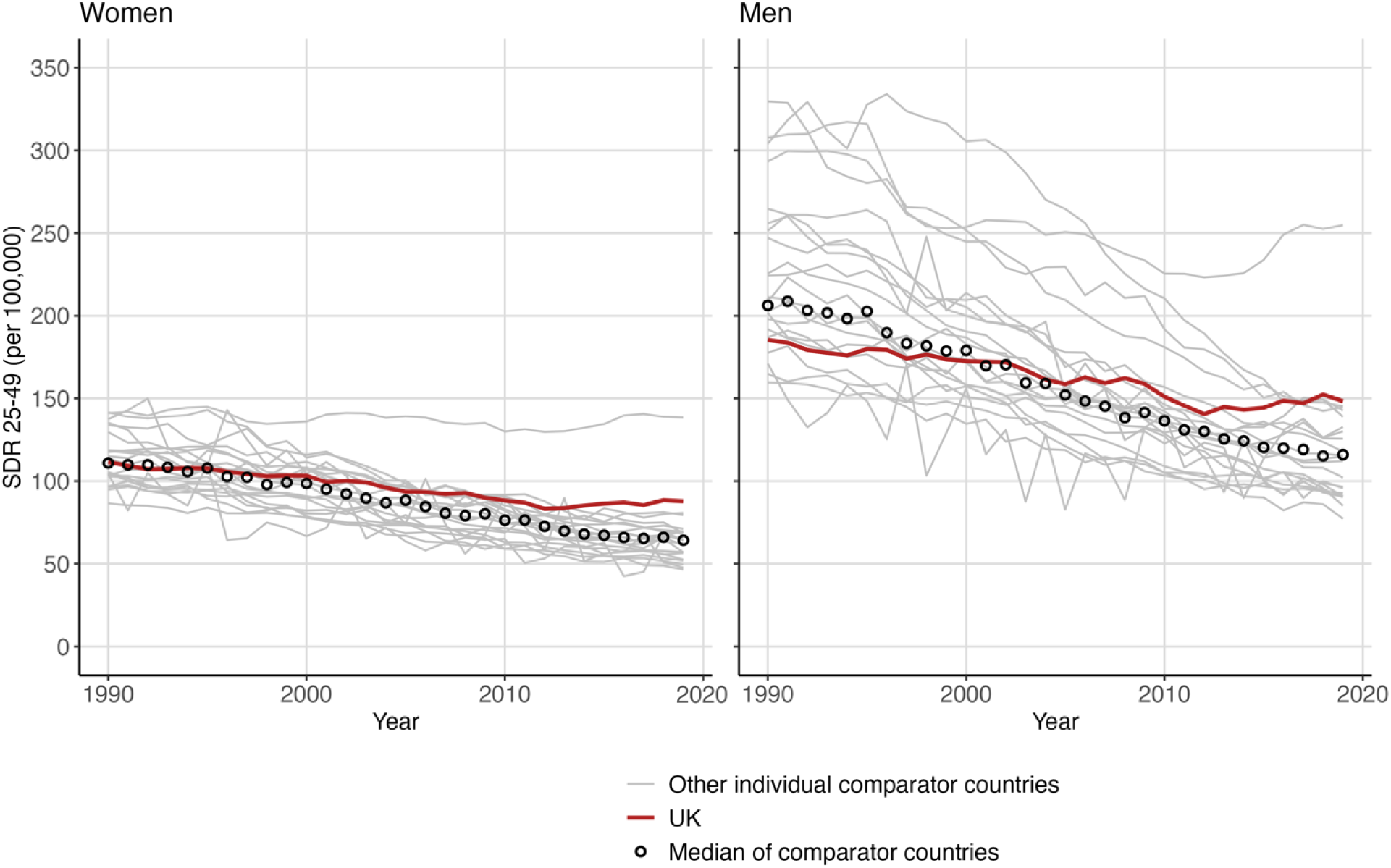
Trends (1990-2019) in age-standardised mortality rates per 100,000 at ages 25-49 for the UK and 21 peer countries plus the comparator median by sex.

The deterioration of the UK’s mortality at age 25-49 years relative to the median is the result of several factors. Firstly, the comparator median showed a steady decline over the entire period 1990-2019. As shown in Supplementary Figure S4, this downward trend in the median is broadly reflected in a similar decline in each of the individual comparator countries with the exception of the USA and Canada. Secondly, compared to the comparator median the UK showed a shallower decline until 2012, after which the UK rates started to increase. It is these contrasting trends that resulted in a reversal of the advantage that males in the UK had in 1990, with the cross-over in rates between the UK and the comparator median being particularly striking.

The public health impact of these negative relative trends is shown in Figure 3, expressed in terms of the annual number of excess deaths at ages 25-49 years in the UK, relative to the counter-factual of rates in each year being equal to the comparator median. For women, the number of annual excess deaths showed a progressive increase from the mid-1990s onwards. For men, the observed deaths were lower than expected in 1990. However, this initial advantage was eroded and reversed by 2000. Between 2001 and 2019 the cumulative total number of excess deaths was 26,733 for women and 33,345 for men. This corresponds to 1.36 million years of life lost for women and 1.70 million for men

**Figure 3.**
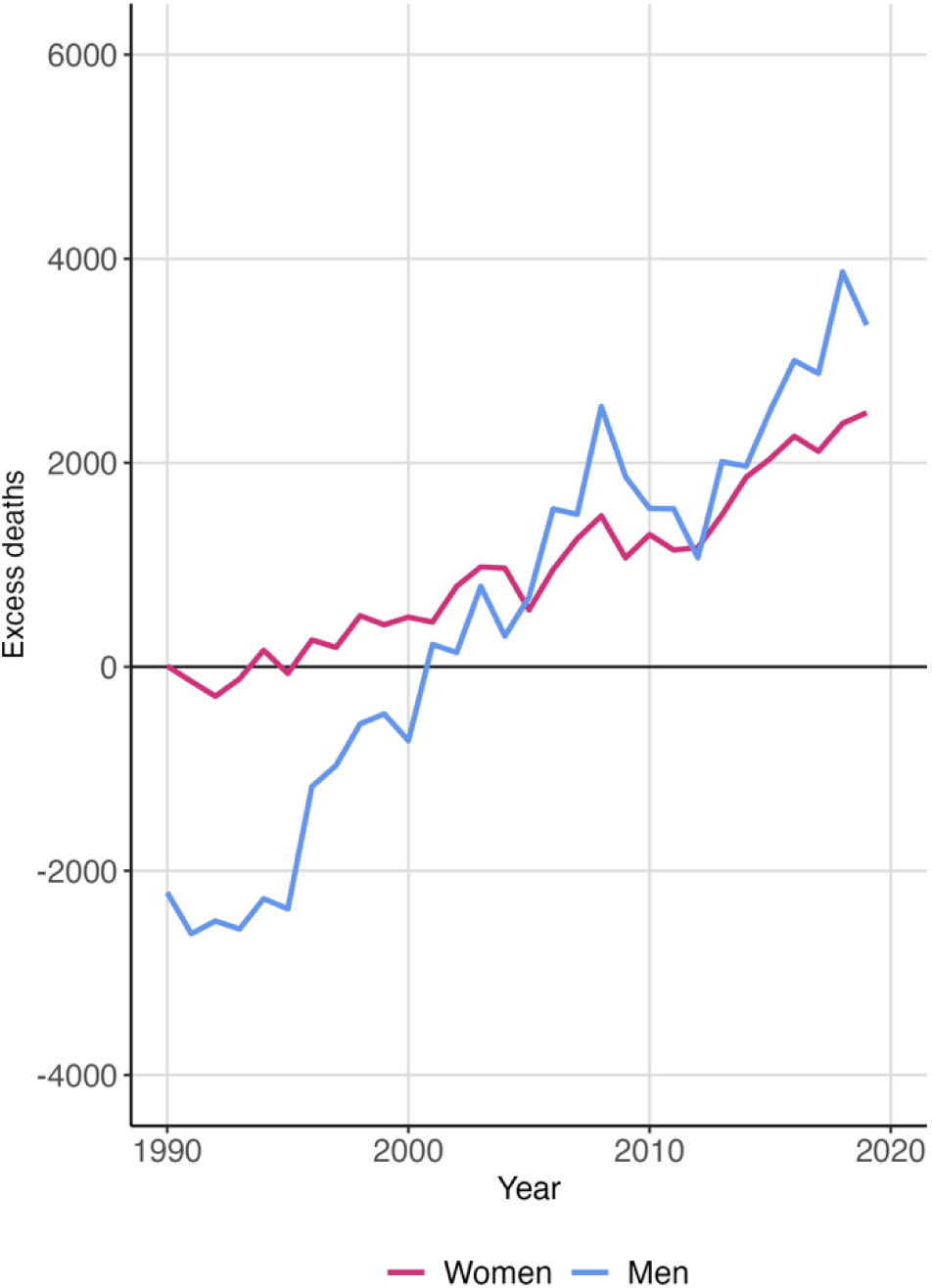
Excess annual deaths in the UK relative to the comparator median of 21 peer countries at age 25-49 years by sex and year (1990-2019).

Do the negative trends for UK mortality at ages 25-49 years relative to its peers apply to all parts of the UK? Figure 4 shows trends in each of the 4 constituent parts of the UK plus the 9 English regions along with the comparator median. The reversal over time of the initial advantage for men compared to the comparator median is seen for all parts of the UK. By 2019 all areas had rates that were higher than the median with the exception of London for men which showed a dramatic improvement across the entire period. In 1990 rates for men in London were only second to that of Scotland, yet by 2019 they were the lowest of any UK area and below that of the comparator median. Annual all-cause mortality rates (1990-2019) for each UK area are given in Supplementary Table S3.

**Figure 4.**
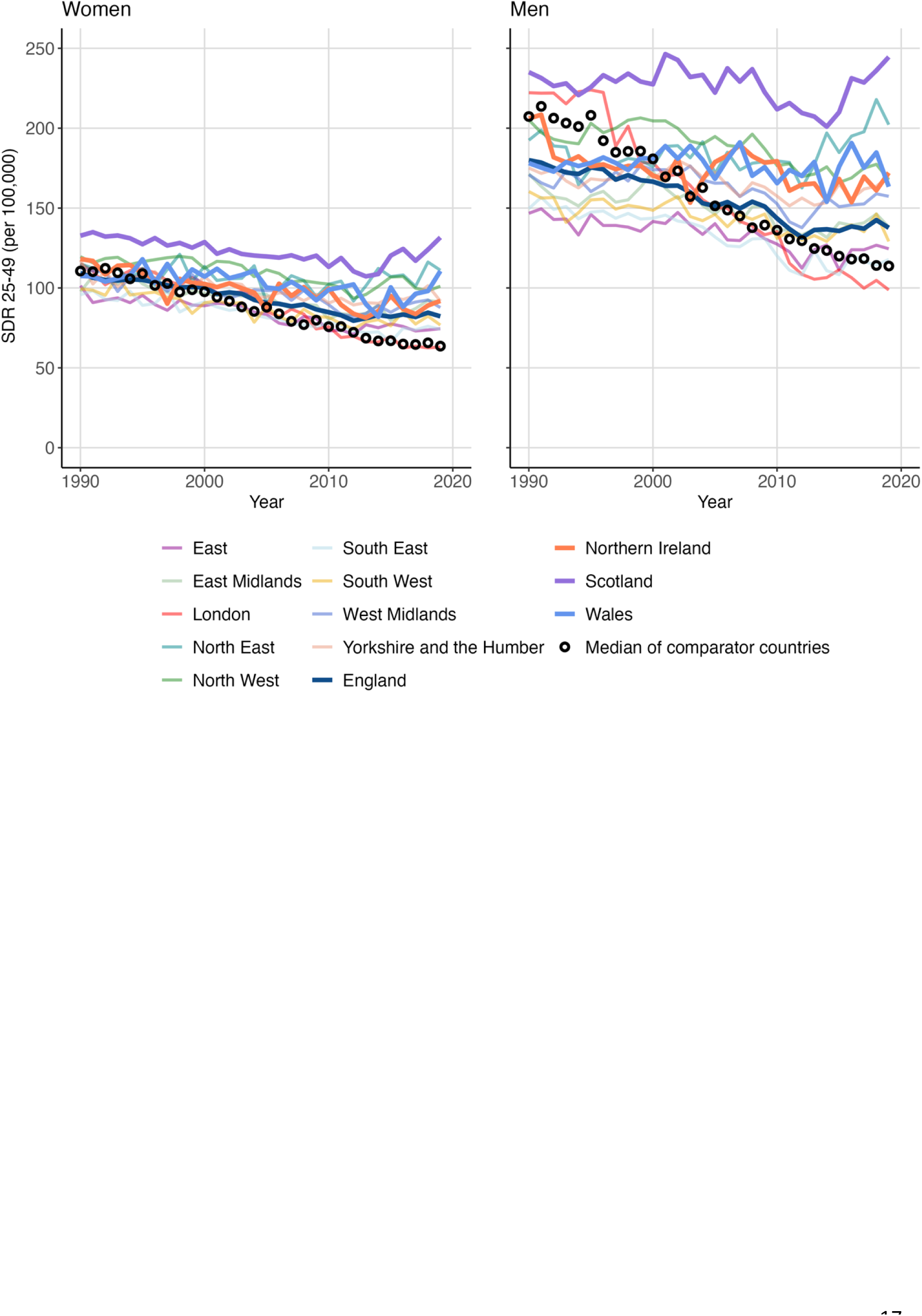
Trends (1990-2019) in age-standardised all-cause mortality rates per 100,000 at ages 25-49 years in the 9 regions of England, Wales, Scotland and Northern Ireland, plus the comparator median of 21 peer countries by sex

The spatial patterns of mortality within the UK relative to that of the comparator median (expressed as rate ratios) are mapped out in Figure 5 for four selected years 1990 to 2019. This shows the predictable North-South gradient. All southern regions had mortality ratios below one for both men and women in 1990 and 2000, with the notable exception of London, which was above one. However, by 2019, the rate ratios in all areas apart from London were above one, with Scotland, Wales (women), and the North East of England being the highest. The trends over time for males and females were similar.

**Figure 5.**
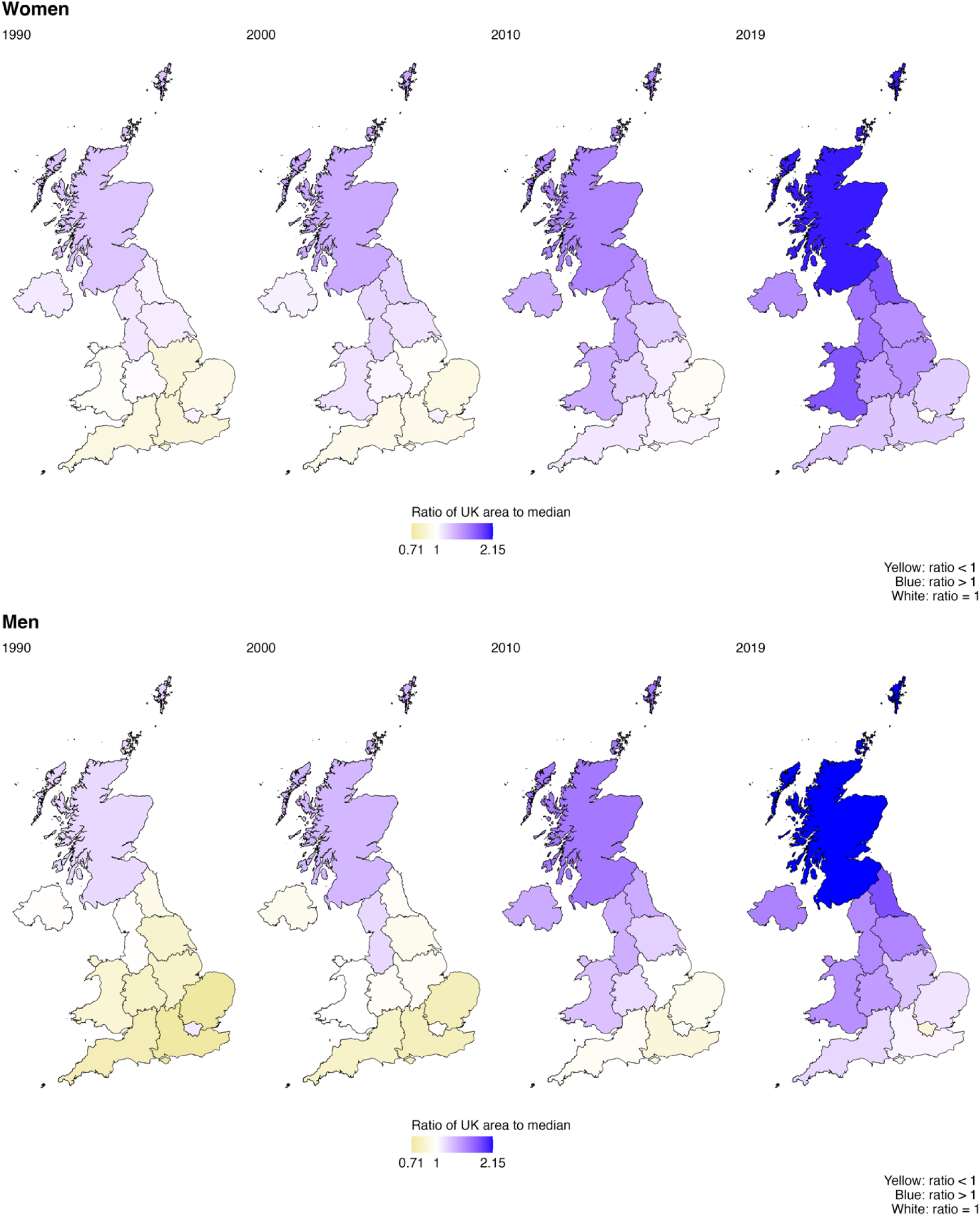
Ratio of all-cause age-standardised mortality rate at ages 25-49 years in each UK area to the comparator median of 21 peer countries for selected years by sex.

We now turn to examine the contribution of causes of death to these negative relative trends. Figure 6 shows for each cause the ratio of mortality rates in the UK to the comparator median level for the period 2001-2019. Of the three ICD10 chapters examined (solid lines), the rate ratios for deaths from external causes show the most marked increases. In 2001, they were below one (UK rates lower than the median), but then started to increase for women from 2008 and for men from 2011. By 2019, UK external cause mortality rates were over 40% higher than the comparator median for both sexes. For malignant neoplasms, the rate ratios showed a shallow increase throughout. Circulatory disease rate ratios were stable for women up until about 2013, after which they increased, while for men, there is a suggestion that they increased steadily over the entire period 2001-19.

**Figure 6.**
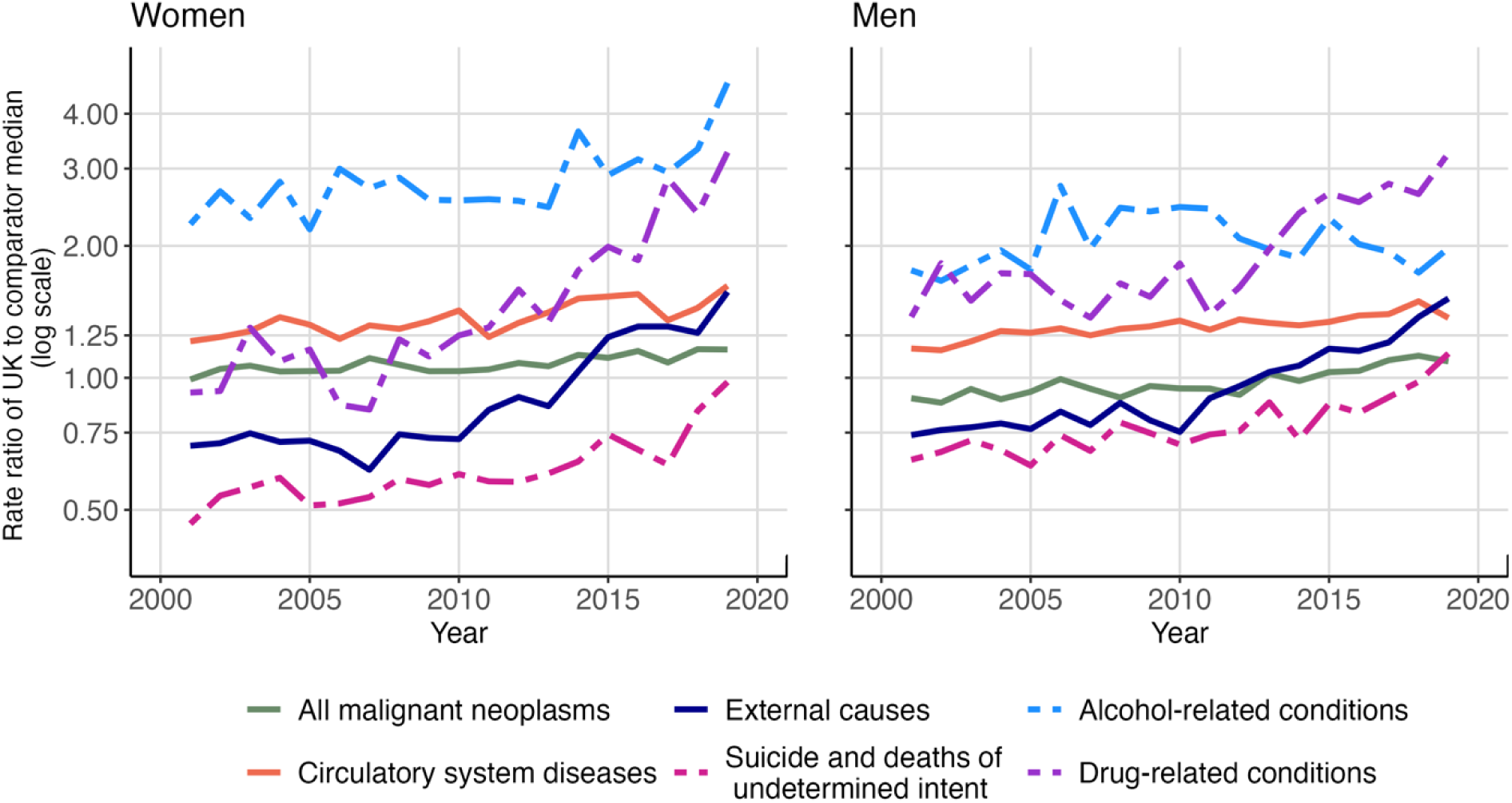
Trends (2001-2019) in ratio of age-standardised mortality rates in the UK relative to the comparator median of 21 peer countries for selected causes of death at age 25-49 years by sex.

Of the rate ratios for the three other causes (Figure 6 dashed lines) drug-related deaths showed the largest increases. For women a pronounced upward trend was evident from 2009 and for men from around 2011. Rate ratios for deaths from alcohol-related causes were the highest of any of the causes for women, and were the highest or second highest for men depending upon the year. However, whereas there was a continuous upward trend for women starting in 2013, for men the rate ratios declined from a high in 2006. Finally, the rate ratios for deaths from suicide and undetermined intent show a tendency to increase over the entire period 2001-19, although this became more pronounced in the most recent years. It is notable that other than 2018-19 for men, the rate ratios for this cause were below one i.e. rates in the UK were lower than the comparator median.

The changes over time in the cause-specific rate ratios are logically a consequence of differences in trends in UK rates compared to those of the comparator median. These can be seen in Figure 7 which shows absolute mortality rate trends (2001-2019) at age 25-49 years for the same cause of death groups. For malignant neoplasms, circulatory system diseases and external causes the comparator medians all showed declines throughout. For each of these causes the trends were less positive for the UK, with rates declining more slowly than the median after 2010 (malignant neoplasms), plateauing after 2010 (circulatory system diseases) or definitively increasing from 2010 (external causes). The difference in trends between the UK and the comparator median were particularly marked for external causes, with a crossover in 2012 for both sexes, with the UK moving from having a mortality advantage to having a disadvantage relative to the comparator median.

**Figure 7.**
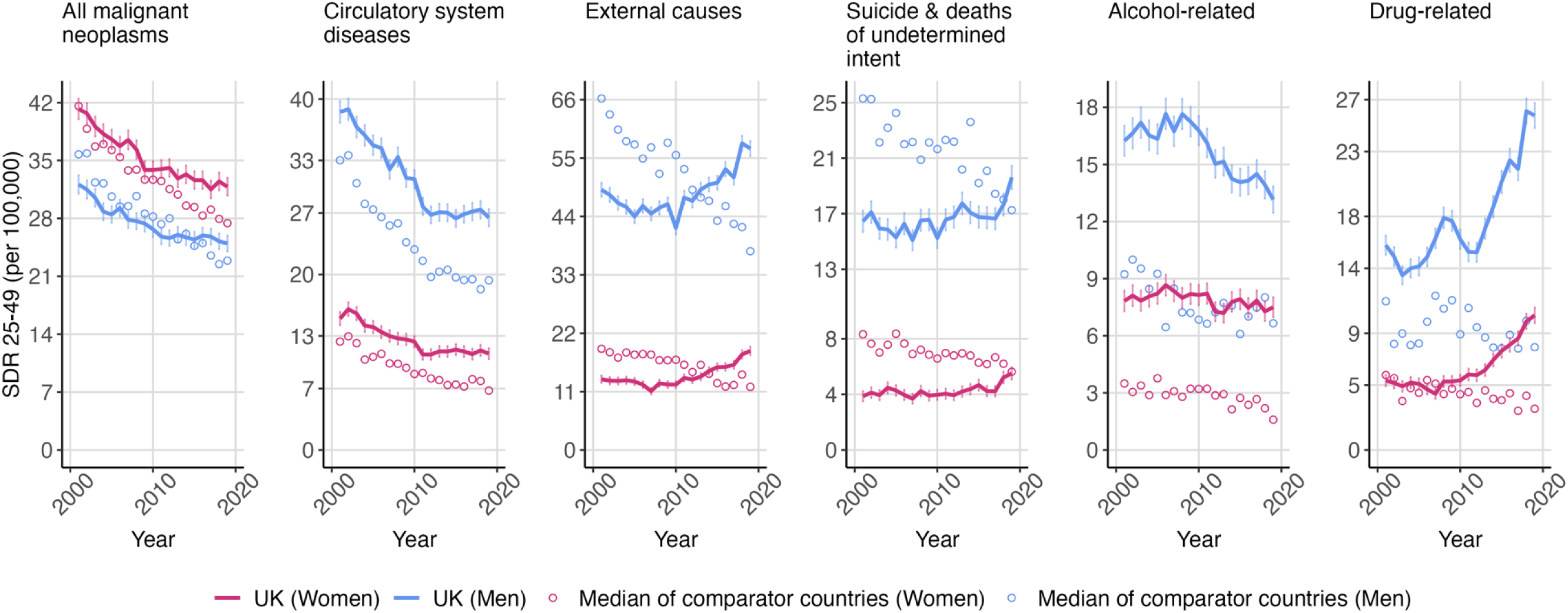
Trends (2001-2019) in age-standardised mortality rates per 100,000 (95% confidence intervals) for selected causes of death at ages 25-49 by sex in the UK plus the comparator median of the 21 peer countries

For the other three cause-groups in Figure 7 (alcohol-related, drug-related and suicides and undetermined intent) among women the comparator median rates declined. In contrast in the UK rates for suicide and undetermined intent and drug-related deaths increased after 2010. Alcohol-related deaths for UK women showed some evidence of decline, but this was not as steep as seen from 2012 for the comparator median. Among men the median rates for all suicide and undetermined cause declined steeply, while for the other two causes median rates tended to be lower after 2010 than in the preceding 10 years. In contrast rates in the UK for suicide and undetermined intent increased from 2017 to reach their highest level in 2019 becoming higher than the comparator median. For alcohol-related causes there was a steep decline in the UK from a peak in 2008. The most dramatic change in the UK was seen for drug-related causes which showed a steep increase from 2012.

Equivalent cause-specific mortality trends for each of the four constituent parts of the UK (England, Wales, Scotland, Northern Ireland) are shown in Supplementary Results Figure S5. The trends described in the previous paragraphs for the UK as a whole in relation to the comparator median have echoes in all of the UKs constituent parts, although for the three smallest populations (Wales, Northern Ireland and Scotland) there is considerable year to year fluctuation largely due to small numbers as reflected in wide confidence intervals.

As we have shown the total mortality gap between the UK and the comparator median rate of its 21 peer countries has steadily widened over the past 30 years. In Table 1 we quantify the contribution of each cause of death to this widening gap between 2001-3 and 2017-19. We have done this by estimating the change in the gap that would have observed if the cause of interest was deleted from total mortality. The difference between this change and the total observed change is the effect of the cause of death. It should be noted that we have added two extra causes to Table 1. The first is the sum of all other ICD chapters (other than malignant neoplasms, circulatory disease and external causes). The second is the sum of effects due to deaths from alcohol- and drug related causes plus suicide and undetermined intent (deaths of despair).

**Table 1.**
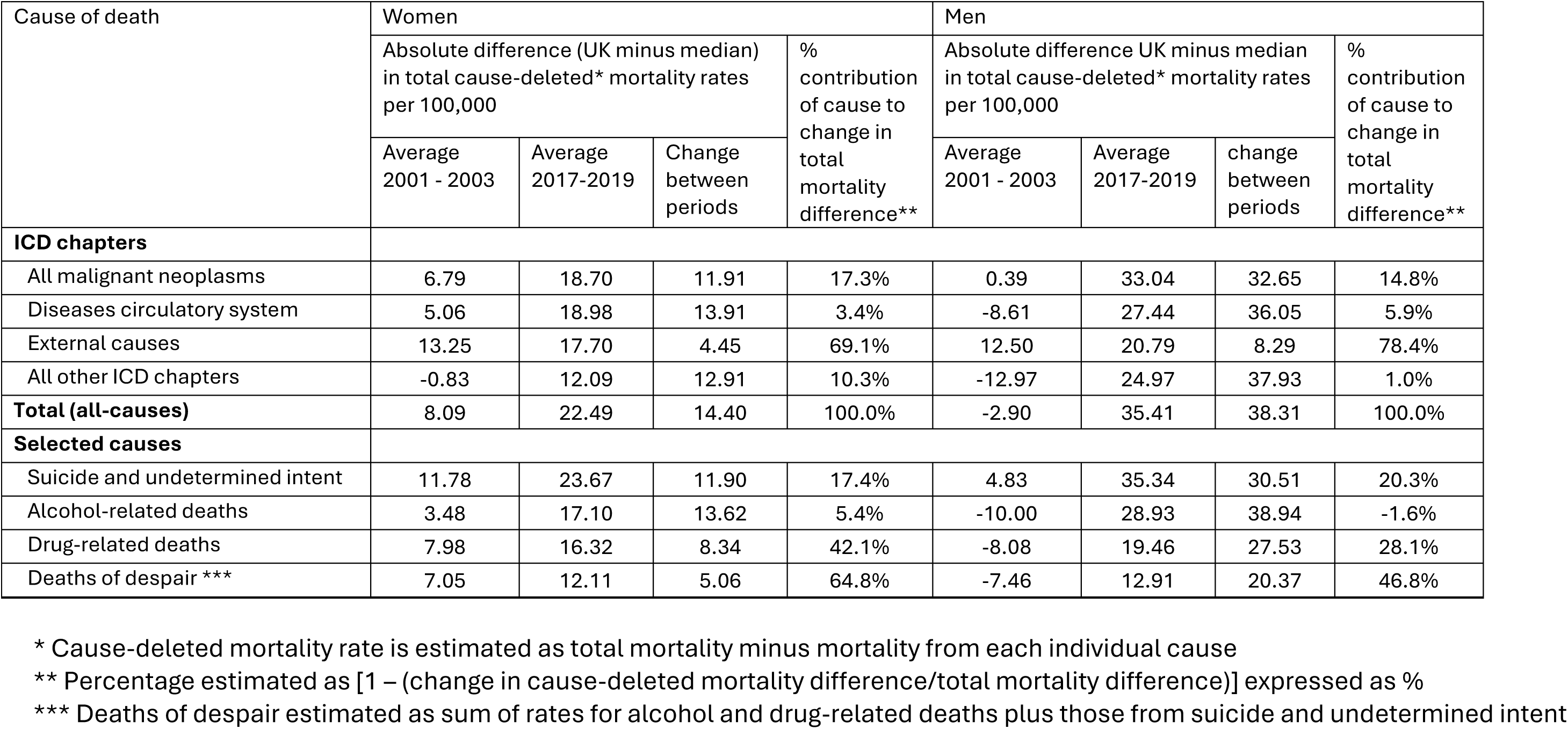
Contribution of specific causes of death to overall widening of difference in mortality between the UK and the comparator median of 21 peer countries between 2001-3 and 2017-19 at age 25-54 years by sex.

| Cause of death | Women |  |  |  | Men |  |  |  |
| --- | --- | --- | --- | --- | --- | --- | --- | --- |
|  | Absolute difference (UK minus median) in total cause-deleted* mortality rates per 100,000 |  |  | % contribution of cause to change in total mortality difference** | Absolute difference UK minus median in total cause-deleted* mortality rates per 100,000 |  |  | % contribution of cause to change in total mortality difference** |
|  | Average 2001 - 2003 | Average 2017-2019 | Change between periods |  | Average 2001 - 2003 | Average 2017-2019 | change between periods |  |
| ICD chapters |  |  |  |  |  |  |  |  |
| All malignant neoplasms | 6.79 | 18.70 | 11.91 | 17.3% | 0.39 | 33.04 | 32.65 | 14.8% |
| Diseases circulatory system | 5.06 | 18.98 | 13.91 | 3.4% | -8.61 | 27.44 | 36.05 | 5.9% |
| External causes | 13.25 | 17.70 | 4.45 | 69.1% | 12.50 | 20.79 | 8.29 | 78.4% |
| All other ICD chapters | -0.83 | 12.09 | 12.91 | 10.3% | -12.97 | 24.97 | 37.93 | 1.0% |
| Total (all-causes) | 8.09 | 22.49 | 14.40 | 100.0% | -2.90 | 35.41 | 38.31 | 100.0% |
| Selected causes |  |  |  |  |  |  |  |  |
| Suicide and undetermined intent | 11.78 | 23.67 | 11.90 | 17.4% | 4.83 | 35.34 | 30.51 | 20.3% |
| Alcohol-related deaths | 3.48 | 17.10 | 13.62 | 5.4% | -10.00 | 28.93 | 38.94 | -1.6% |
| Drug-related deaths | 7.98 | 16.32 | 8.34 | 42.1% | -8.08 | 19.46 | 27.53 | 28.1% |
| Deaths of despair *** | 7.05 | 12.11 | 5.06 | 64.8% | -7.46 | 12.91 | 20.37 | 46.8% |

\* Cause-deleted mortality rate is estimated as total mortality minus mortality from each individual cause
\*\* Percentage estimated as $[1 - (\text{change in cause-deleted mortality difference} / \text{total mortality difference})]$ expressed as %
\*\*\* Deaths of despair estimated as sum of rates for alcohol and drug-related deaths plus those from suicide and undetermined intent

For both sexes the largest percentage contributor was external causes : 69% for women and 78% for men. Of the other chapters malignant neoplasms made the next largest contribution: 17% (women) and 15% (men). Diseases of the circulatory system made only a small contribution : of 3% (women) and 6% (men). Deaths of despair contributed 65% for women and 47% for men. This big difference between the sexes is mainly due to the much larger contribution of drug-related deaths for women (42%) than for men (28%). It is notable that suicide and deaths from undetermined intent accounted for an appreciable percentage of the gap in both sexes : 17% (women) and 20% (men). Alcohol made only a small contribution.

## Discussion

We have shown that over the past 30 years mortality among working age adults under 50 has undergone a substantial and negative transformation relative to other high income countries. Among men in particular, the UK has gone from having one of the lowest mortality rates compared to its peers to having one of the highest. By 2019 it was second only to the USA. This reflects both the better progress made by many other countries in reducing mortality rates at this age (25-49 years) as well as an absolute increase in mortality rates in all parts of the UK from around 2013. The divergence in rates between 2001 and 2019 has been principally driven by external cause mortality as a whole, and the overlapping categories of drug-related deaths and suicides, all of which increased in the UK after 2010. Surprisingly, divergence in mortality from malignant neoplasms also played a significant role.

We have identified only one other study that addressed a similar question to ours although it focussed on Scotland with the rest of the UK being treated as a single entity^14^ It compared all-cause mortality rates by single years of age and calendar period with the mean for 8 Western European countries. Like us, they noted the reversal of the UK’s international advantage over time among younger adults. However, this study only went up to 2010 and did not explore this reversal in any depth.

More generally, there has been increasing concern about negative mortality trends among younger adults in high income countries, most notably the USA.^15,16^ Several international comparative studies of mortality rates ^17–19^ and life-expectancy^20^ have variously highlighted faltering progress or negative trends in all-cause mortality among working-age adults in the UK, USA, Canada, Australia, as well as in Sweden and Poland. A number of these studies have attempted to isolate the specific causes that underlie these negative trends. Deaths of despair, or specific components of this group of causes, have been highlighted as important contributors to the increasing mortality seen in middle-age or younger adults in the USA, UK, Canada.^6,17,19^ Overall, consistent with our findings, these investigations have identified the UK, or component parts of it, as being an international outlier in terms of rising young-adult mortality and the contribution of components of deaths of despair to this.

What does our study add to the existing literature? We have quantified the substantial public health impact of the UK’s failure to reduce adult mortality under the age of 50 years at the same rate as the central tendency (median) of many other high income countries: between 2001 and 2019 this resulted in 3.1 million years of life lost between. Most importantly we have also quantified the contribution of specific causes of death to this divergence. Our findings of an appreciable contribution from drug-related deaths aligns with the results of other studies,^6,19^ however our finding that this cause contributes more to the divergence with other countries among women (42%) than men (28%) in striking and novel.

The notion “deaths of despair”^5^ encapsulates a wide range of potential causal mechanisms including absolute and relative deprivation and associated states of hopelessness which drive people to behaviours that are self-harming and potentially fatal and result in deaths that are caused by drugs, alcohol or suicide. More recent work, however, has brought into question the coherence of how this concept of deaths of despair is assumed to be driven by a common set of psychosocial factors.^6,21,22^ Our results support this to the extent that we find alcohol-related causes have made little contribution to the divergence of the UK with its peers unlike drug-related deaths and suicides.

Moving beyond the deaths of despair we have found that divergence in mortality rates from malignant neoplasms also make an important contribution to the total divergence (around 15% for women and men). Cancers have not previously received little attention in this context. Finally, with respect to cause, we have been able to demonstrate that deaths from circulatory system diseases play only a very minor role in the divergence of adult mortality under age 50.

Our study is the first to consider mortality in the UK as a whole and its constituent parts relative to peer countries. What is striking is that the negative relative trends seen for the UK as a whole are seen across the UK, including each of the English standard regions. The magnitude of the relative difference between UK areas and the comparator median followed the well-known North-South pattern of geographic inequalities across the UK. While England showed the smallest differences with the comparator median, Scotland showed much larger differences with the comparator median, with Wales and Northern Ireland being intermediate. However, within England itself, there was a clear North-South gradient, with notably high rates relative to the comparator median in the North East of England. By 2019, London was the only part of the UK that has lower mortality at age 25-49 years than the comparator median. Thus, the impact and challenge of these trends is a truly national issue that speaks to the urgent need to improve the life chances of everyone in the UK.

Much of the literature concerning negative trends in mortality in the UK and other countries has focussed on the health effects of government austerity following the 2008 economic crisis.^3,23,24^ This cannot be the whole explanation for the divergence of the UK with its peers as we have shown that this was happening across the entire period from 1990 onwards. In the twenty years from 1990 to 2010 the median rate for the 21 comparator peer countries declined at a steeper rate than the UK. Our cause-specific analyses show that this was occurring from 2001 onwards for all malignant neoplasms, external causes as a whole and deaths from suicide and undetermined intent. This longer-term negative trend in the UK’s international position, that pre-date the 2008 financial crisis, has been largely overlooked and is not understood. Nevertheless, our study does show that after the 2008 financial crisis the underlying long-term divergence in mortality at ages 25-49 years accelerated not least because of sharp increases in mortality from drug-related causes and suicides from around 2010. As argued by others,^25^ this may indeed be in part driven by the effects of austerity in the UK,^26,27^ which did not perturb the overall downward trends in the median of the 21 comparator countries. This is despite the fact that there is some evidence that the 2008 financial crisis did adversely affect health and mortality in a number of high income countries.^3,23,28,29^

The main weakness of this study, and others which attempt to compare cause-specific mortality across countries and time, is that there may be changes and inconsistencies in how cause is certified and coded^30^ (see Supplementary Methods for more details). While the ICD codes we used to construct the various cause of death categories have been consistently applied across all countries included in these analyses, the criteria used by certifying and coding authorities in each country may vary. Further work in this area is warranted.

There is now an urgent need to understand how and why many peer countries made much better progress from 1990 than the UK in reducing mortality from a range of causes including all malignant neoplasms. Attention should also be given to understanding the atypical, positive trajectory of male adult mortality under the age of 50 years in London. How far does this reflect changes in the socio-economic profile of London over time, the availability of services and standard of health care or biased estimates of population or the healthy-migrant effect?

In conclusion, mortality rates among younger adults in all parts of the UK have diverged from the general positive tendency seen for most peer countries. This has resulted in a substantial burden of excess deaths and years of life lost. By their very nature, the central role of external causes and of drug-related causes (among men) contributing to these trends indicates that these deaths were potentially avoidable. The divergence in mortality rates at ages 25-49 years in the UK from peer countries was already apparent from 1990 pre-dating the imposition of government austerity policies. Nevertheless, austerity may well have contributed to this longer-term deterioration in the UK’s position. The fact that all areas of the UK showed a similar deterioration in position relative to peer countries underlines the fact that this is a truly national problem.

## Data Availability

All data produced in the present study are available upon reasonable request to the authors

## Authors’ Contributions

DAL, VMS and DAJ conceived of the study and contributed to study design. DAL and DAJ acquired the data. DAJ and VMS undertook the statistical analyses. NM-D generated the tables and figures and ID contributed to the analysis of cause-specific trends. DAL led the drafting of the manuscript. All authors contributed to interpretation of results and drafting, revision, and approval of the manuscript.

## Acknowledgement

We would like to thank Pablo Perel for his helpful comments on an earlier draft of this manuscript.

## Ethics committee approval

The analyses reported in this paper fall within the larger study *Trends in life expectancy, age- and cause-specific mortality in the UK, 1970–2022, in comparison with a set of 22 high-income countries: an analysis of vital statistics data* that received ethical approval from the London School of Hygiene & Tropical Medicine’s ethics committee on 13 September 2023 Ref: 29803

---

**Figure S1.**
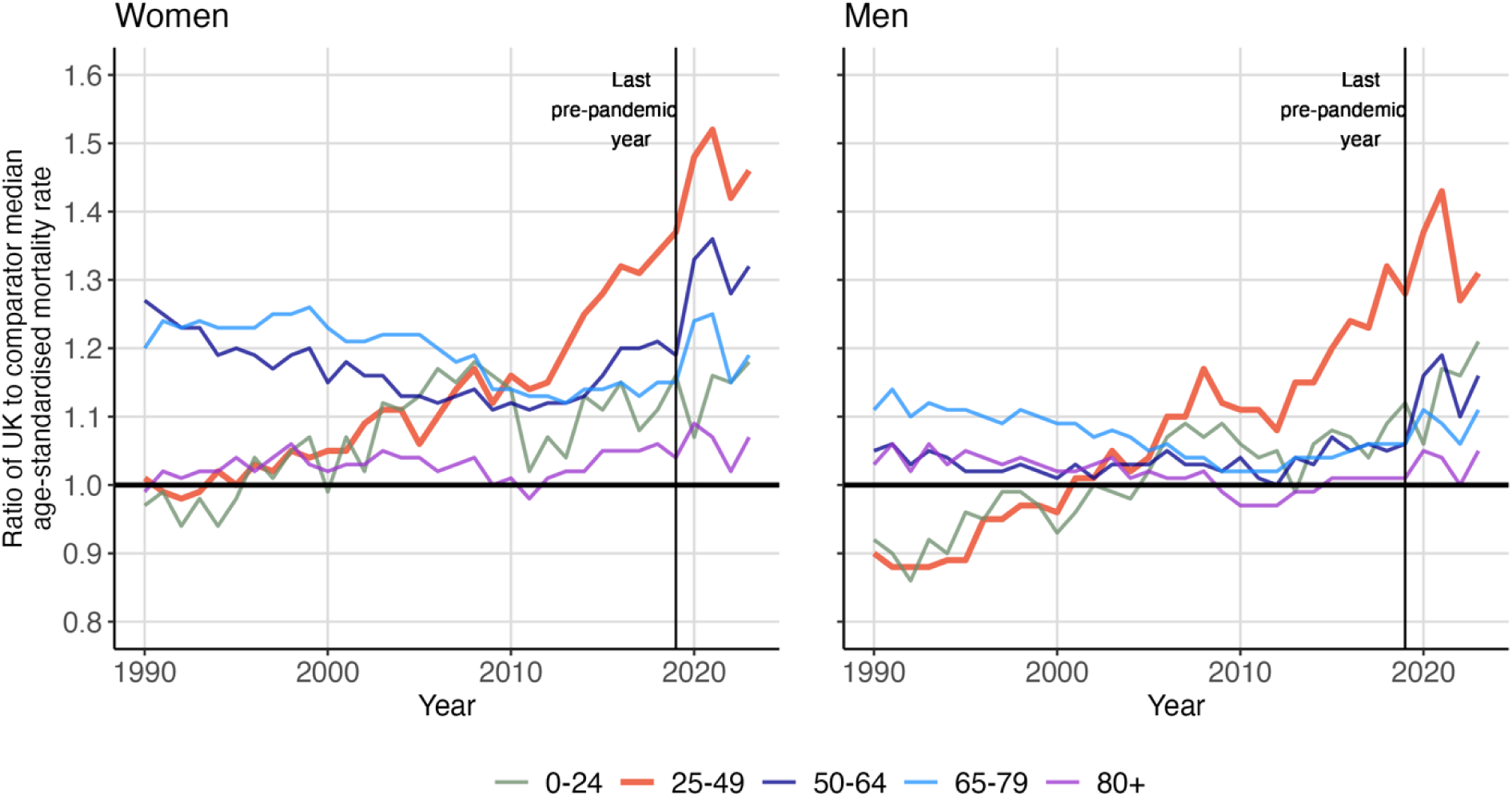
Trends (1990-2023) in ratio of all-cause age-standardised mortality rates in the UK relative to the comparator median of 21 peer countries by age-group and sex.

**Figure S2.**
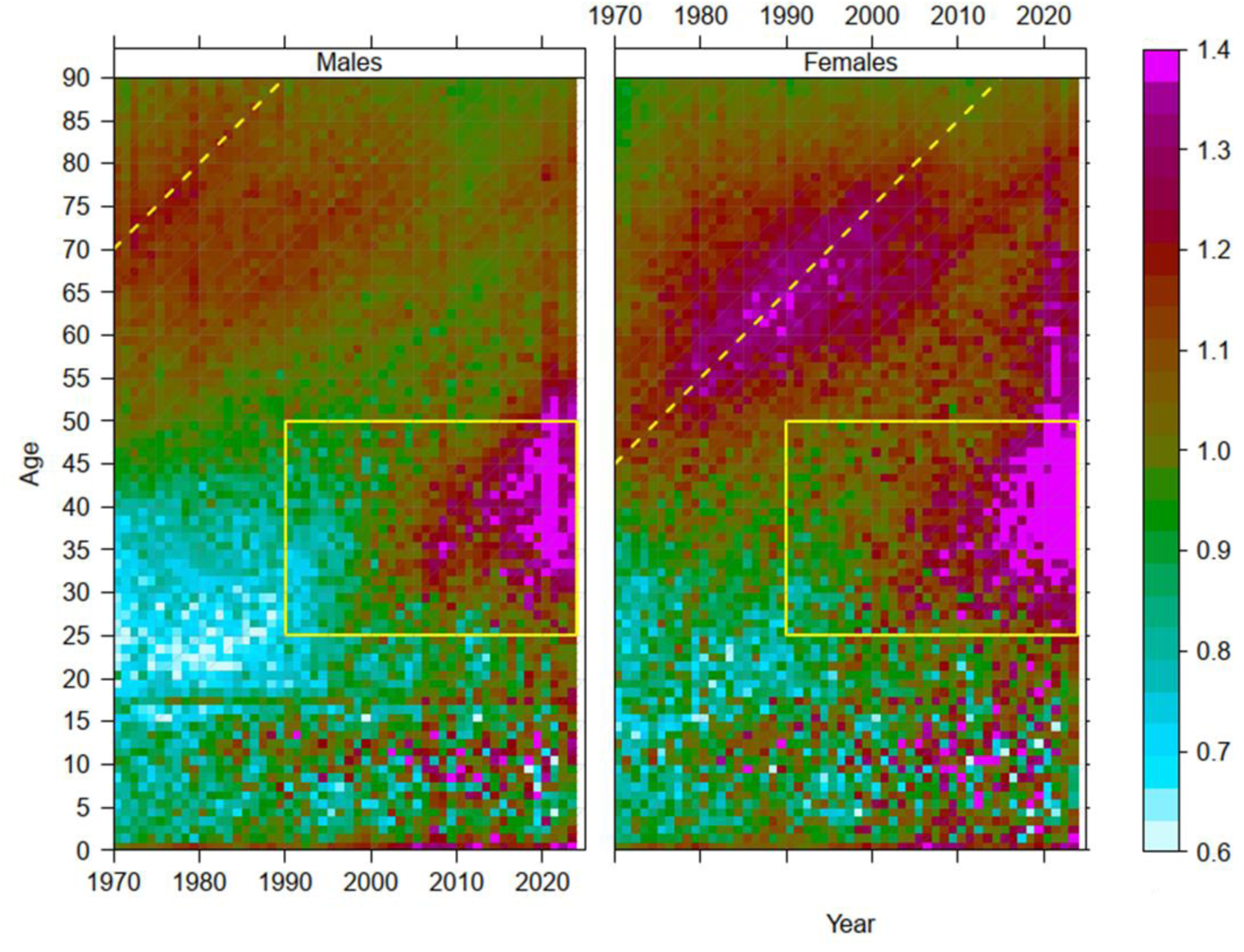
Mortality rate ratios in the UK relative to the median for 21 comparator countries by age, year, and sex for 1970-2023. Notes. The rectangle delineates the range of ages 25 to 49 years within the period 1990-2023. The diagonal lines correspond to the male and female cohorts of maximal smoking prevalence born in 1900 and 1925, respectively. For more details see Leon, DA et al. Trends in life expectancy and age-specific mortality in England and Wales, 1970–2016, in comparison with a set of 22 high-income countries: an analysis of vital statistics data. The Lancet Public Health, Volume 4, Issue 11, e575 - e582

**Figure S3.**
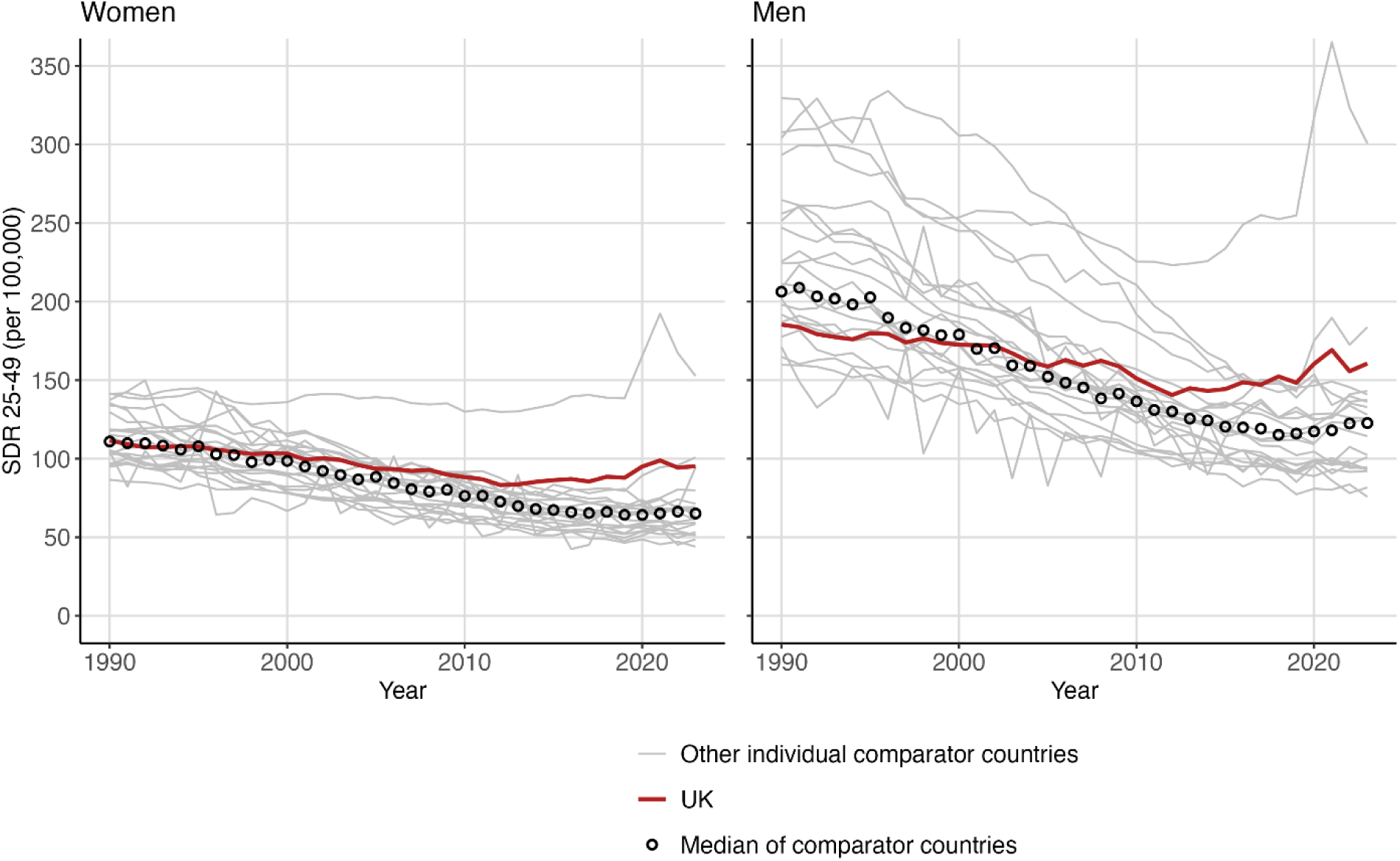
Trends (1990-2023) in age-standardised mortality rates per 100,000 at ages 25-49 for the UK and 21 peer countries plus the comparator median by sex

**Figure S4.**
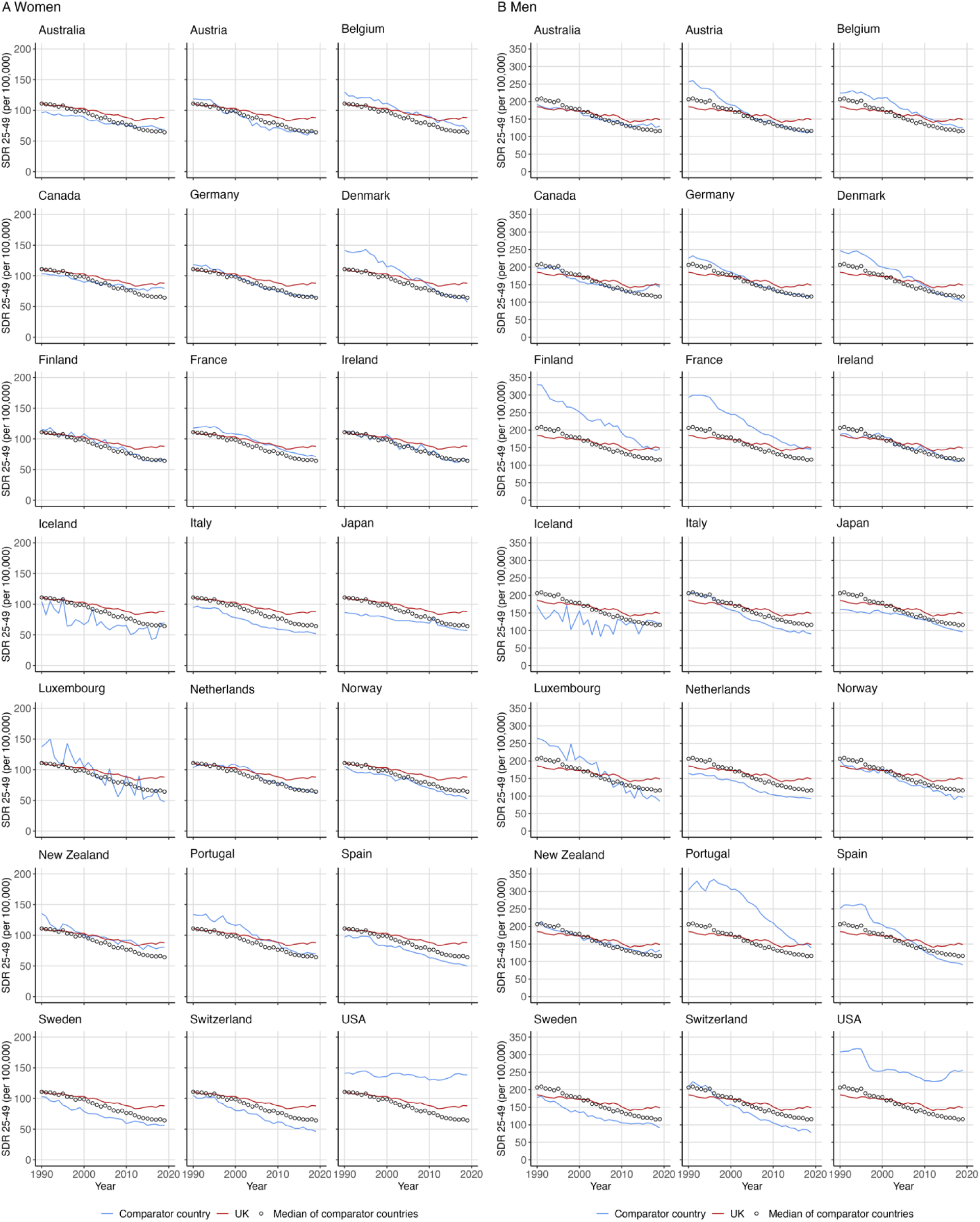
Age-standardised all-cause mortality rates per 100,000 by year (1990-2019) at ages 25-49 years for women (A) and men (B) in the UK plus the comparator median for each of the other 21 comparator countries

**Figure S5.**
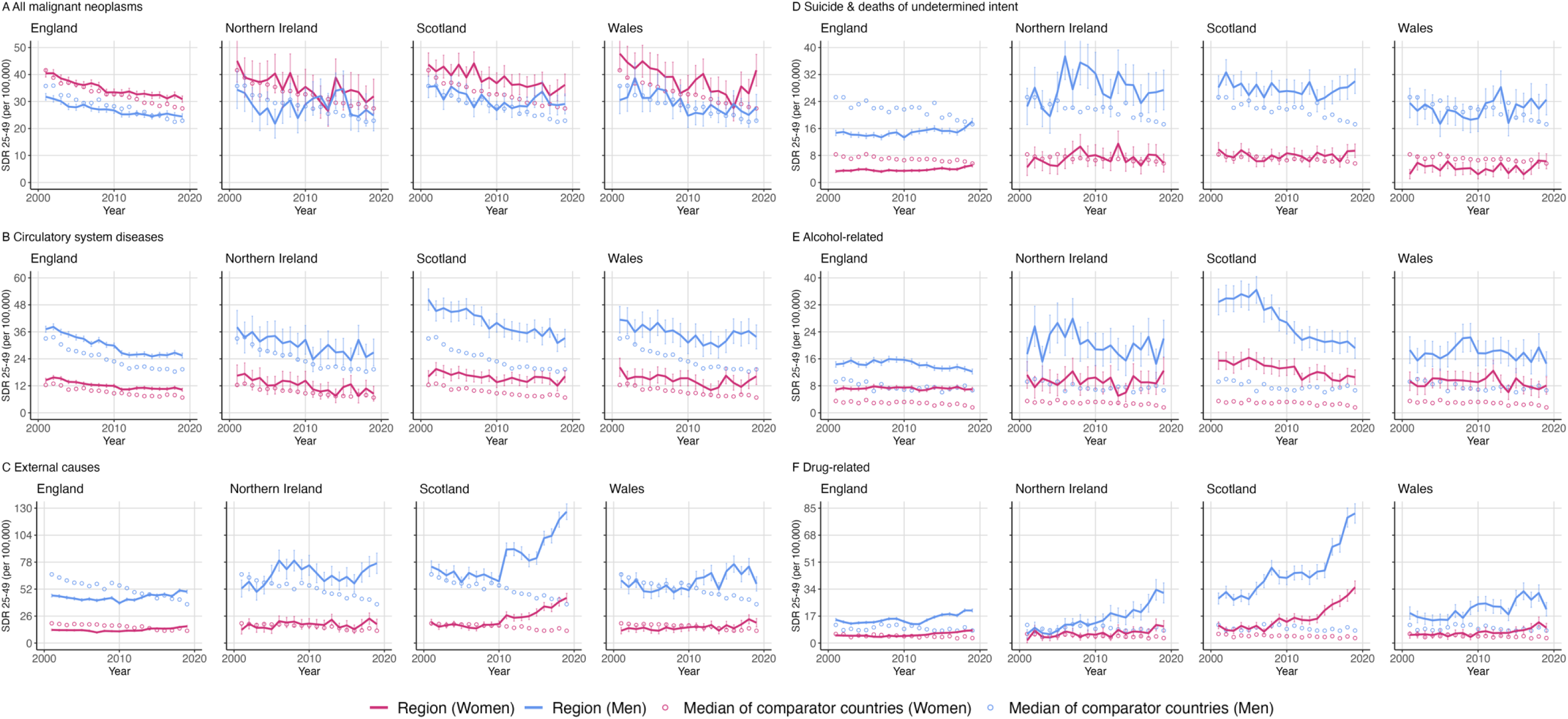
Age-standardised mortality rates per 100,000 with 95% confidence intervals for selected causes of death by year (2001-2019) for ages 25-49 by sex in the four constituent parts of the UK plus the median for the 21 comparator countries

**Table S1.**
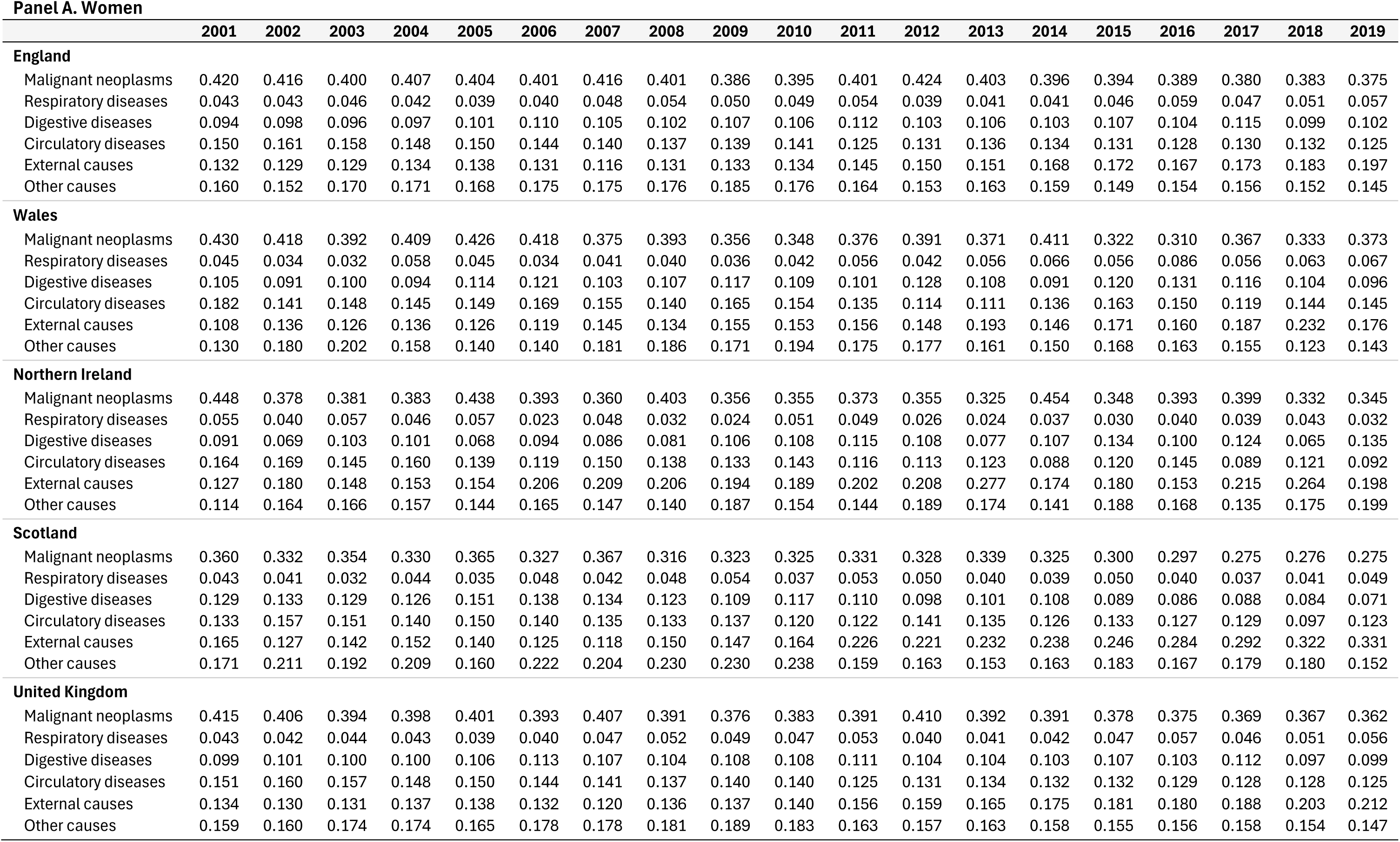
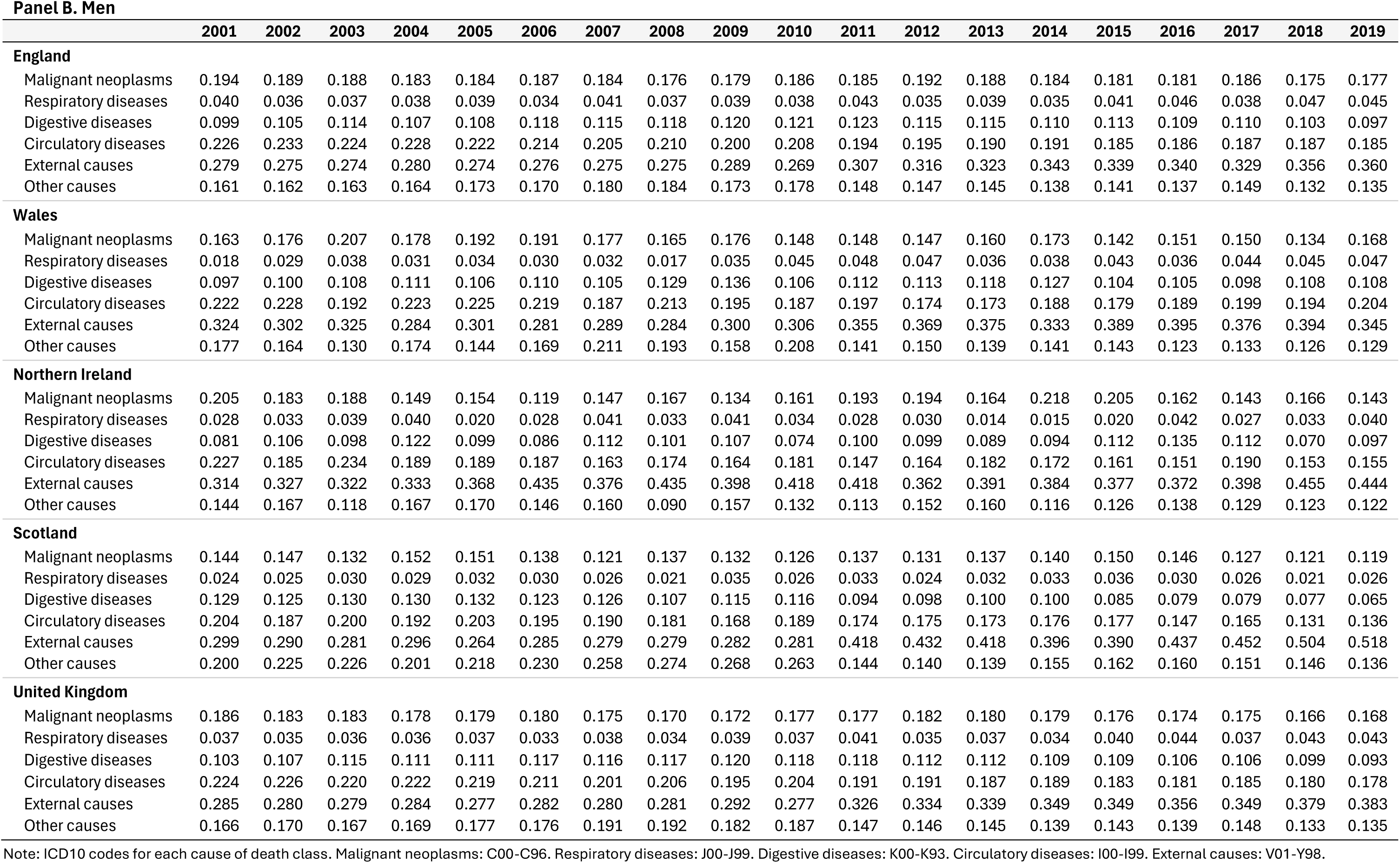
Cause-specific age-standardised mortality rates per 100,000 at ages 25-49 years as a proportion of all-cause mortality rates by year (2001-2019) for women (A) and men (B) in England, Wales, Northern Ireland, Scotland and the UK.

**Panel A. Women**
|  | 2001 | 2002 | 2003 | 2004 | 2005 | 2006 | 2007 | 2008 | 2009 | 2010 | 2011 | 2012 | 2013 | 2014 | 2015 | 2016 | 2017 | 2018 | 2019 |
| --- | --- | --- | --- | --- | --- | --- | --- | --- | --- | --- | --- | --- | --- | --- | --- | --- | --- | --- | --- |
| **England** |  |  |  |  |  |  |  |  |  |  |  |  |  |  |  |  |  |  |  |
| Malignant neoplasms | 0.420 | 0.416 | 0.400 | 0.407 | 0.404 | 0.401 | 0.416 | 0.401 | 0.386 | 0.395 | 0.401 | 0.424 | 0.403 | 0.396 | 0.394 | 0.389 | 0.380 | 0.383 | 0.375 |
| Respiratory diseases | 0.043 | 0.043 | 0.046 | 0.042 | 0.039 | 0.040 | 0.048 | 0.054 | 0.050 | 0.049 | 0.054 | 0.039 | 0.041 | 0.041 | 0.046 | 0.059 | 0.047 | 0.051 | 0.057 |
| Digestive diseases | 0.094 | 0.098 | 0.096 | 0.097 | 0.101 | 0.110 | 0.105 | 0.102 | 0.107 | 0.106 | 0.112 | 0.103 | 0.106 | 0.103 | 0.107 | 0.104 | 0.115 | 0.099 | 0.102 |
| Circulatory diseases | 0.150 | 0.161 | 0.158 | 0.148 | 0.150 | 0.144 | 0.140 | 0.137 | 0.139 | 0.141 | 0.125 | 0.131 | 0.136 | 0.134 | 0.131 | 0.128 | 0.130 | 0.132 | 0.125 |
| External causes | 0.132 | 0.129 | 0.129 | 0.134 | 0.138 | 0.131 | 0.116 | 0.131 | 0.133 | 0.134 | 0.145 | 0.150 | 0.151 | 0.168 | 0.172 | 0.167 | 0.173 | 0.183 | 0.197 |
| Other causes | 0.160 | 0.152 | 0.170 | 0.171 | 0.168 | 0.175 | 0.175 | 0.176 | 0.185 | 0.176 | 0.164 | 0.153 | 0.163 | 0.159 | 0.149 | 0.154 | 0.156 | 0.152 | 0.145 |
| **Wales** |  |  |  |  |  |  |  |  |  |  |  |  |  |  |  |  |  |  |  |
| Malignant neoplasms | 0.430 | 0.418 | 0.392 | 0.409 | 0.426 | 0.418 | 0.375 | 0.393 | 0.356 | 0.348 | 0.376 | 0.391 | 0.371 | 0.411 | 0.322 | 0.310 | 0.367 | 0.333 | 0.373 |
| Respiratory diseases | 0.045 | 0.034 | 0.032 | 0.058 | 0.045 | 0.034 | 0.041 | 0.040 | 0.036 | 0.042 | 0.056 | 0.042 | 0.056 | 0.066 | 0.056 | 0.086 | 0.056 | 0.063 | 0.067 |
| Digestive diseases | 0.105 | 0.091 | 0.100 | 0.094 | 0.114 | 0.121 | 0.103 | 0.107 | 0.117 | 0.109 | 0.101 | 0.128 | 0.108 | 0.091 | 0.120 | 0.131 | 0.116 | 0.104 | 0.096 |
| Circulatory diseases | 0.182 | 0.141 | 0.148 | 0.145 | 0.149 | 0.169 | 0.155 | 0.140 | 0.165 | 0.154 | 0.135 | 0.114 | 0.111 | 0.136 | 0.163 | 0.150 | 0.119 | 0.144 | 0.145 |
| External causes | 0.108 | 0.136 | 0.126 | 0.136 | 0.126 | 0.119 | 0.145 | 0.134 | 0.155 | 0.153 | 0.156 | 0.148 | 0.193 | 0.146 | 0.171 | 0.160 | 0.187 | 0.232 | 0.176 |
| Other causes | 0.130 | 0.180 | 0.202 | 0.158 | 0.140 | 0.140 | 0.181 | 0.186 | 0.171 | 0.194 | 0.175 | 0.177 | 0.161 | 0.150 | 0.168 | 0.163 | 0.155 | 0.123 | 0.143 |
| **Northern Ireland** |  |  |  |  |  |  |  |  |  |  |  |  |  |  |  |  |  |  |  |
| Malignant neoplasms | 0.448 | 0.378 | 0.381 | 0.383 | 0.438 | 0.393 | 0.360 | 0.403 | 0.356 | 0.355 | 0.373 | 0.355 | 0.325 | 0.454 | 0.348 | 0.393 | 0.399 | 0.332 | 0.345 |
| Respiratory diseases | 0.055 | 0.040 | 0.057 | 0.046 | 0.057 | 0.023 | 0.048 | 0.032 | 0.024 | 0.051 | 0.049 | 0.026 | 0.024 | 0.037 | 0.030 | 0.040 | 0.039 | 0.043 | 0.032 |
| Digestive diseases | 0.091 | 0.069 | 0.103 | 0.101 | 0.068 | 0.094 | 0.086 | 0.081 | 0.106 | 0.108 | 0.115 | 0.108 | 0.077 | 0.107 | 0.134 | 0.100 | 0.124 | 0.065 | 0.135 |
| Circulatory diseases | 0.164 | 0.169 | 0.145 | 0.160 | 0.139 | 0.119 | 0.150 | 0.138 | 0.133 | 0.143 | 0.116 | 0.113 | 0.123 | 0.088 | 0.120 | 0.145 | 0.089 | 0.121 | 0.092 |
| External causes | 0.127 | 0.180 | 0.148 | 0.153 | 0.154 | 0.206 | 0.209 | 0.206 | 0.194 | 0.189 | 0.202 | 0.208 | 0.277 | 0.174 | 0.180 | 0.153 | 0.215 | 0.264 | 0.198 |
| Other causes | 0.114 | 0.164 | 0.166 | 0.157 | 0.144 | 0.165 | 0.147 | 0.140 | 0.187 | 0.154 | 0.144 | 0.189 | 0.174 | 0.141 | 0.188 | 0.168 | 0.135 | 0.175 | 0.199 |
| **Scotland** |  |  |  |  |  |  |  |  |  |  |  |  |  |  |  |  |  |  |  |
| Malignant neoplasms | 0.360 | 0.332 | 0.354 | 0.330 | 0.365 | 0.327 | 0.367 | 0.316 | 0.323 | 0.325 | 0.331 | 0.328 | 0.339 | 0.325 | 0.300 | 0.297 | 0.275 | 0.276 | 0.275 |
| Respiratory diseases | 0.043 | 0.041 | 0.032 | 0.044 | 0.035 | 0.048 | 0.042 | 0.048 | 0.054 | 0.037 | 0.053 | 0.050 | 0.040 | 0.039 | 0.050 | 0.040 | 0.037 | 0.041 | 0.049 |
| Digestive diseases | 0.129 | 0.133 | 0.129 | 0.126 | 0.151 | 0.138 | 0.134 | 0.123 | 0.109 | 0.117 | 0.110 | 0.098 | 0.101 | 0.108 | 0.089 | 0.086 | 0.088 | 0.084 | 0.071 |
| Circulatory diseases | 0.133 | 0.157 | 0.151 | 0.140 | 0.150 | 0.140 | 0.135 | 0.133 | 0.137 | 0.120 | 0.122 | 0.141 | 0.135 | 0.126 | 0.133 | 0.127 | 0.129 | 0.097 | 0.123 |
| External causes | 0.165 | 0.127 | 0.142 | 0.152 | 0.140 | 0.125 | 0.118 | 0.150 | 0.147 | 0.164 | 0.226 | 0.221 | 0.232 | 0.238 | 0.246 | 0.284 | 0.292 | 0.322 | 0.331 |
| Other causes | 0.171 | 0.211 | 0.192 | 0.209 | 0.160 | 0.222 | 0.204 | 0.230 | 0.230 | 0.238 | 0.159 | 0.163 | 0.153 | 0.163 | 0.183 | 0.167 | 0.179 | 0.180 | 0.152 |
| **United Kingdom** |  |  |  |  |  |  |  |  |  |  |  |  |  |  |  |  |  |  |  |
| Malignant neoplasms | 0.415 | 0.406 | 0.394 | 0.398 | 0.401 | 0.393 | 0.407 | 0.391 | 0.376 | 0.383 | 0.391 | 0.410 | 0.392 | 0.391 | 0.378 | 0.375 | 0.369 | 0.367 | 0.362 |
| Respiratory diseases | 0.043 | 0.042 | 0.044 | 0.043 | 0.039 | 0.040 | 0.047 | 0.052 | 0.049 | 0.047 | 0.053 | 0.040 | 0.041 | 0.042 | 0.047 | 0.057 | 0.046 | 0.051 | 0.056 |
| Digestive diseases | 0.099 | 0.101 | 0.100 | 0.100 | 0.106 | 0.113 | 0.107 | 0.104 | 0.108 | 0.108 | 0.111 | 0.104 | 0.104 | 0.103 | 0.107 | 0.103 | 0.112 | 0.097 | 0.099 |
| Circulatory diseases | 0.151 | 0.160 | 0.157 | 0.148 | 0.150 | 0.144 | 0.141 | 0.137 | 0.140 | 0.140 | 0.125 | 0.131 | 0.134 | 0.132 | 0.132 | 0.129 | 0.128 | 0.128 | 0.125 |
| External causes | 0.134 | 0.130 | 0.131 | 0.137 | 0.138 | 0.132 | 0.120 | 0.136 | 0.137 | 0.140 | 0.156 | 0.159 | 0.165 | 0.175 | 0.181 | 0.180 | 0.188 | 0.203 | 0.212 |
| Other causes | 0.159 | 0.160 | 0.174 | 0.174 | 0.165 | 0.178 | 0.178 | 0.181 | 0.189 | 0.183 | 0.163 | 0.157 | 0.163 | 0.158 | 0.155 | 0.156 | 0.158 | 0.154 | 0.147 |

**Table S1. (continued)** **Panel B. Men**
|  | 2001 | 2002 | 2003 | 2004 | 2005 | 2006 | 2007 | 2008 | 2009 | 2010 | 2011 | 2012 | 2013 | 2014 | 2015 | 2016 | 2017 | 2018 | 2019 |
| --- | --- | --- | --- | --- | --- | --- | --- | --- | --- | --- | --- | --- | --- | --- | --- | --- | --- | --- | --- |
| **England** |  |  |  |  |  |  |  |  |  |  |  |  |  |  |  |  |  |  |  |
| Malignant neoplasms | 0.194 | 0.189 | 0.188 | 0.183 | 0.184 | 0.187 | 0.184 | 0.176 | 0.179 | 0.186 | 0.185 | 0.192 | 0.188 | 0.184 | 0.181 | 0.181 | 0.186 | 0.175 | 0.177 |
| Respiratory diseases | 0.040 | 0.036 | 0.037 | 0.038 | 0.039 | 0.034 | 0.041 | 0.037 | 0.039 | 0.038 | 0.043 | 0.035 | 0.039 | 0.035 | 0.041 | 0.046 | 0.038 | 0.047 | 0.045 |
| Digestive diseases | 0.099 | 0.105 | 0.114 | 0.107 | 0.108 | 0.118 | 0.115 | 0.118 | 0.120 | 0.121 | 0.123 | 0.115 | 0.115 | 0.110 | 0.113 | 0.109 | 0.110 | 0.103 | 0.097 |
| Circulatory diseases | 0.226 | 0.233 | 0.224 | 0.228 | 0.222 | 0.214 | 0.205 | 0.210 | 0.200 | 0.208 | 0.194 | 0.195 | 0.190 | 0.191 | 0.185 | 0.186 | 0.187 | 0.187 | 0.185 |
| External causes | 0.279 | 0.275 | 0.274 | 0.280 | 0.274 | 0.276 | 0.275 | 0.275 | 0.289 | 0.269 | 0.307 | 0.316 | 0.323 | 0.343 | 0.339 | 0.340 | 0.329 | 0.356 | 0.360 |
| Other causes | 0.161 | 0.162 | 0.163 | 0.164 | 0.173 | 0.170 | 0.180 | 0.184 | 0.173 | 0.178 | 0.148 | 0.147 | 0.145 | 0.138 | 0.141 | 0.137 | 0.149 | 0.132 | 0.135 |
| **Wales** |  |  |  |  |  |  |  |  |  |  |  |  |  |  |  |  |  |  |  |
| Malignant neoplasms | 0.163 | 0.176 | 0.207 | 0.178 | 0.192 | 0.191 | 0.177 | 0.165 | 0.176 | 0.148 | 0.148 | 0.147 | 0.160 | 0.173 | 0.142 | 0.151 | 0.150 | 0.134 | 0.168 |
| Respiratory diseases | 0.018 | 0.029 | 0.038 | 0.031 | 0.034 | 0.030 | 0.032 | 0.017 | 0.035 | 0.045 | 0.048 | 0.047 | 0.036 | 0.038 | 0.043 | 0.036 | 0.044 | 0.045 | 0.047 |
| Digestive diseases | 0.097 | 0.100 | 0.108 | 0.111 | 0.106 | 0.110 | 0.105 | 0.129 | 0.136 | 0.106 | 0.112 | 0.113 | 0.118 | 0.127 | 0.104 | 0.105 | 0.098 | 0.108 | 0.108 |
| Circulatory diseases | 0.222 | 0.228 | 0.192 | 0.223 | 0.225 | 0.219 | 0.187 | 0.213 | 0.195 | 0.187 | 0.197 | 0.174 | 0.173 | 0.188 | 0.179 | 0.189 | 0.199 | 0.194 | 0.204 |
| External causes | 0.324 | 0.302 | 0.325 | 0.284 | 0.301 | 0.281 | 0.289 | 0.284 | 0.300 | 0.306 | 0.355 | 0.369 | 0.375 | 0.333 | 0.389 | 0.395 | 0.376 | 0.394 | 0.345 |
| Other causes | 0.177 | 0.164 | 0.130 | 0.174 | 0.144 | 0.169 | 0.211 | 0.193 | 0.158 | 0.208 | 0.141 | 0.150 | 0.139 | 0.141 | 0.143 | 0.123 | 0.133 | 0.126 | 0.129 |
| **Northern Ireland** |  |  |  |  |  |  |  |  |  |  |  |  |  |  |  |  |  |  |  |
| Malignant neoplasms | 0.205 | 0.183 | 0.188 | 0.149 | 0.154 | 0.119 | 0.147 | 0.167 | 0.134 | 0.161 | 0.193 | 0.194 | 0.164 | 0.218 | 0.205 | 0.162 | 0.143 | 0.166 | 0.143 |
| Respiratory diseases | 0.028 | 0.033 | 0.039 | 0.040 | 0.020 | 0.028 | 0.041 | 0.033 | 0.041 | 0.034 | 0.028 | 0.030 | 0.014 | 0.015 | 0.020 | 0.042 | 0.027 | 0.033 | 0.040 |
| Digestive diseases | 0.081 | 0.106 | 0.098 | 0.122 | 0.099 | 0.086 | 0.112 | 0.101 | 0.107 | 0.074 | 0.100 | 0.099 | 0.089 | 0.094 | 0.112 | 0.135 | 0.112 | 0.070 | 0.097 |
| Circulatory diseases | 0.227 | 0.185 | 0.234 | 0.189 | 0.189 | 0.187 | 0.163 | 0.174 | 0.164 | 0.181 | 0.147 | 0.164 | 0.182 | 0.172 | 0.161 | 0.151 | 0.190 | 0.153 | 0.155 |
| External causes | 0.314 | 0.327 | 0.322 | 0.333 | 0.368 | 0.435 | 0.376 | 0.435 | 0.398 | 0.418 | 0.418 | 0.362 | 0.391 | 0.384 | 0.377 | 0.372 | 0.398 | 0.455 | 0.444 |
| Other causes | 0.144 | 0.167 | 0.118 | 0.167 | 0.170 | 0.146 | 0.160 | 0.090 | 0.157 | 0.132 | 0.113 | 0.152 | 0.160 | 0.116 | 0.126 | 0.138 | 0.129 | 0.123 | 0.122 |
| **Scotland** |  |  |  |  |  |  |  |  |  |  |  |  |  |  |  |  |  |  |  |
| Malignant neoplasms | 0.144 | 0.147 | 0.132 | 0.152 | 0.151 | 0.138 | 0.121 | 0.137 | 0.132 | 0.126 | 0.137 | 0.131 | 0.137 | 0.140 | 0.150 | 0.146 | 0.127 | 0.121 | 0.119 |
| Respiratory diseases | 0.024 | 0.025 | 0.030 | 0.029 | 0.032 | 0.030 | 0.026 | 0.021 | 0.035 | 0.026 | 0.033 | 0.024 | 0.032 | 0.033 | 0.036 | 0.030 | 0.026 | 0.021 | 0.026 |
| Digestive diseases | 0.129 | 0.125 | 0.130 | 0.130 | 0.132 | 0.123 | 0.126 | 0.107 | 0.115 | 0.116 | 0.094 | 0.098 | 0.100 | 0.100 | 0.085 | 0.079 | 0.079 | 0.077 | 0.065 |
| Circulatory diseases | 0.204 | 0.187 | 0.200 | 0.192 | 0.203 | 0.195 | 0.190 | 0.181 | 0.168 | 0.189 | 0.174 | 0.175 | 0.173 | 0.176 | 0.177 | 0.147 | 0.165 | 0.131 | 0.136 |
| External causes | 0.299 | 0.290 | 0.281 | 0.296 | 0.264 | 0.285 | 0.279 | 0.279 | 0.282 | 0.281 | 0.418 | 0.432 | 0.418 | 0.396 | 0.390 | 0.437 | 0.452 | 0.504 | 0.518 |
| Other causes | 0.200 | 0.225 | 0.226 | 0.201 | 0.218 | 0.230 | 0.258 | 0.274 | 0.268 | 0.263 | 0.144 | 0.140 | 0.139 | 0.155 | 0.162 | 0.160 | 0.151 | 0.146 | 0.136 |
| **United Kingdom** |  |  |  |  |  |  |  |  |  |  |  |  |  |  |  |  |  |  |  |
| Malignant neoplasms | 0.186 | 0.183 | 0.183 | 0.178 | 0.179 | 0.180 | 0.175 | 0.170 | 0.172 | 0.177 | 0.177 | 0.182 | 0.180 | 0.179 | 0.176 | 0.174 | 0.175 | 0.166 | 0.168 |
| Respiratory diseases | 0.037 | 0.035 | 0.036 | 0.036 | 0.037 | 0.033 | 0.038 | 0.034 | 0.039 | 0.037 | 0.041 | 0.035 | 0.037 | 0.034 | 0.040 | 0.044 | 0.037 | 0.043 | 0.043 |
| Digestive diseases | 0.103 | 0.107 | 0.115 | 0.111 | 0.111 | 0.117 | 0.116 | 0.117 | 0.120 | 0.118 | 0.118 | 0.112 | 0.112 | 0.109 | 0.109 | 0.106 | 0.106 | 0.099 | 0.093 |
| Circulatory diseases | 0.224 | 0.226 | 0.220 | 0.222 | 0.219 | 0.211 | 0.201 | 0.206 | 0.195 | 0.204 | 0.191 | 0.191 | 0.187 | 0.189 | 0.183 | 0.181 | 0.185 | 0.180 | 0.178 |
| External causes | 0.285 | 0.280 | 0.279 | 0.284 | 0.277 | 0.282 | 0.280 | 0.281 | 0.292 | 0.277 | 0.326 | 0.334 | 0.339 | 0.349 | 0.349 | 0.356 | 0.349 | 0.379 | 0.383 |
| Other causes | 0.166 | 0.170 | 0.167 | 0.169 | 0.177 | 0.176 | 0.191 | 0.192 | 0.182 | 0.187 | 0.147 | 0.146 | 0.145 | 0.139 | 0.143 | 0.139 | 0.148 | 0.133 | 0.135 |

Note: ICD10 codes for each cause of death class. Malignant neoplasms: C00-C96. Respiratory diseases: J00-J99. Digestive diseases: K00-K93. Circulatory diseases: I00-I99. External causes: V01-Y98.

**Supplementary Table S2.**
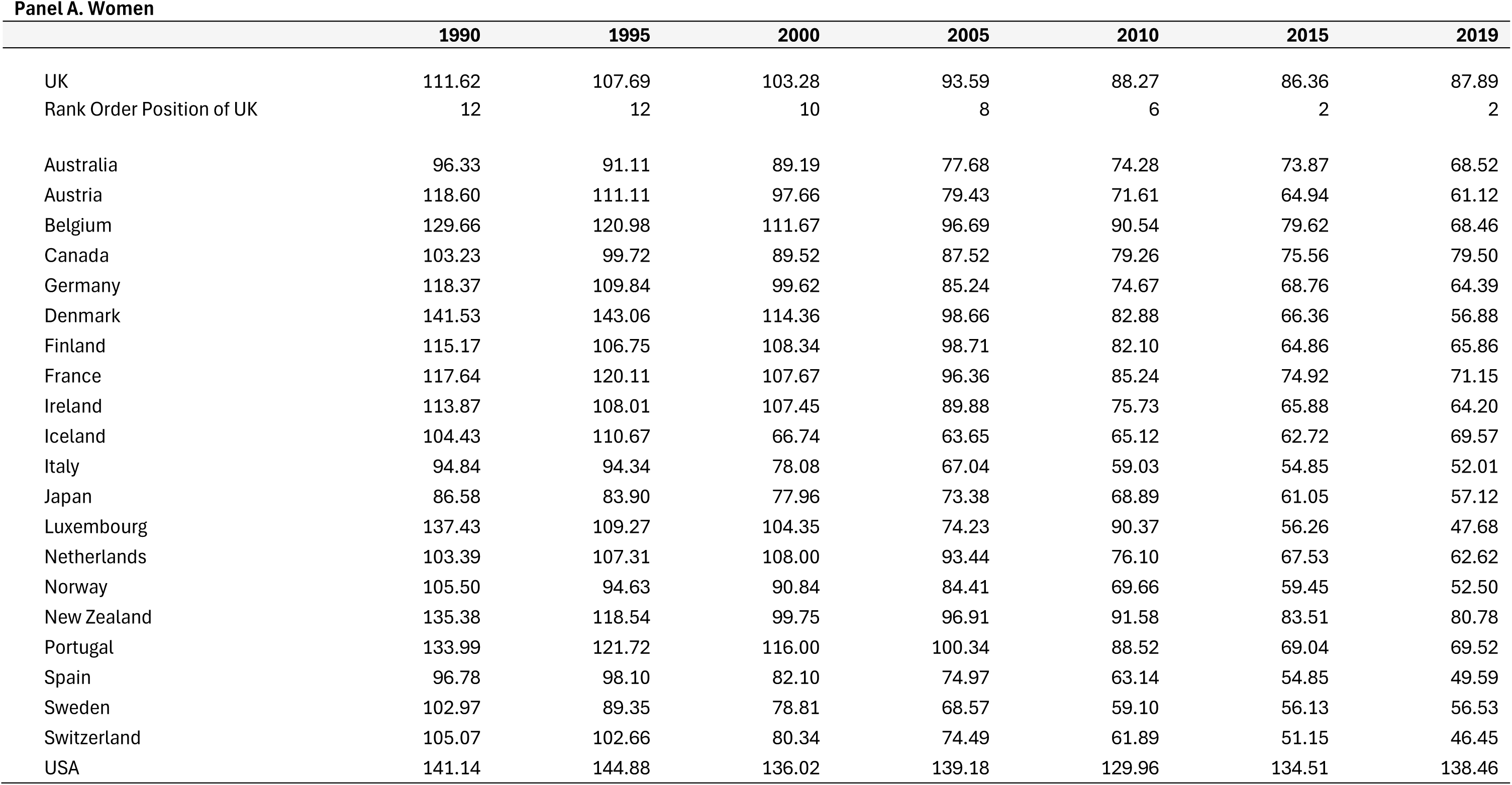
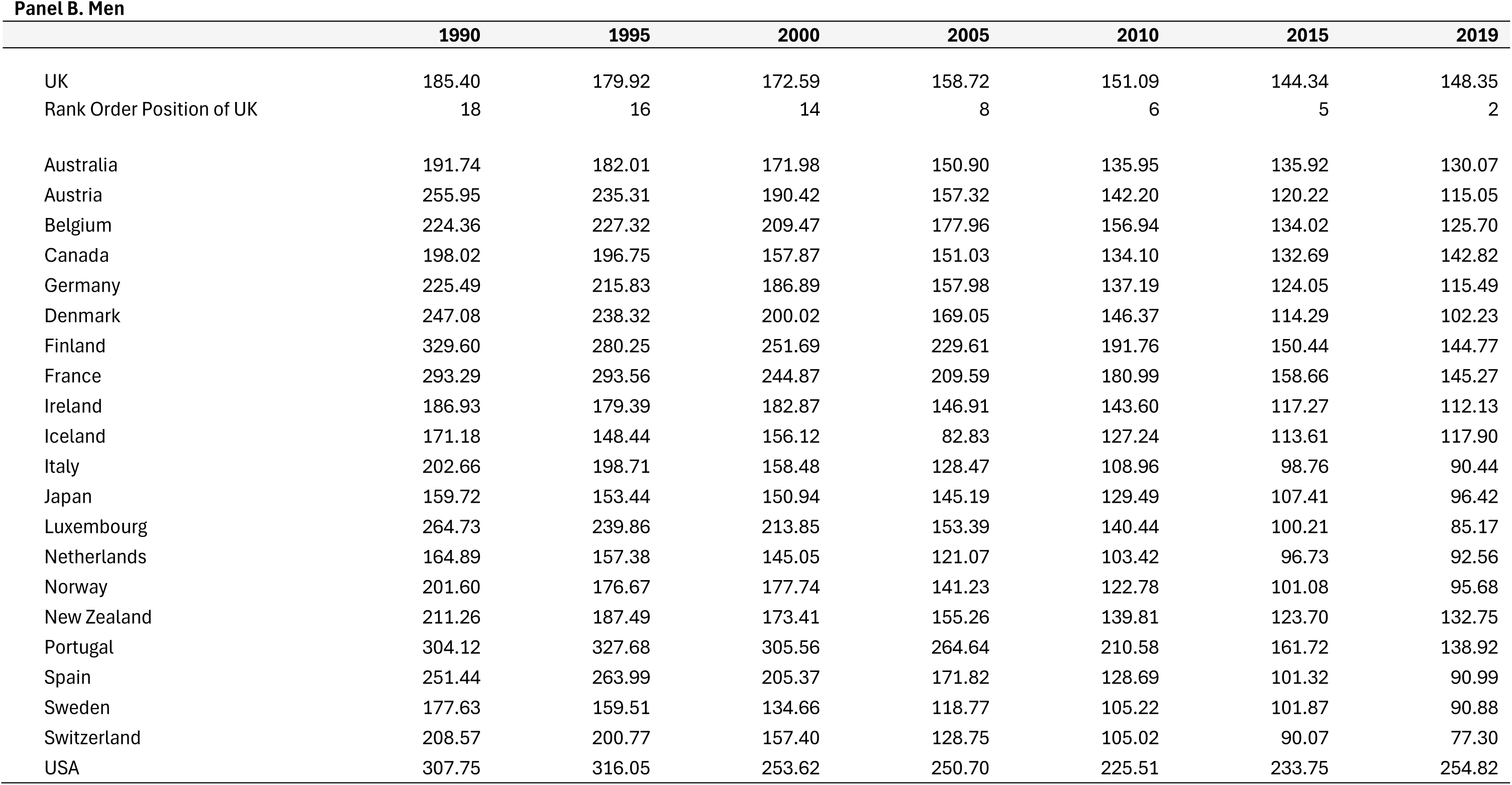
All-causes age-standardised mortality rates per 100,000 at ages 25-49 years for selected years for each of the 21 comparator countries plus the UK along with and it’s corresponding rank position, by sex

**Panel A. Women**
|  | 1990 | 1995 | 2000 | 2005 | 2010 | 2015 | 2019 |
| --- | --- | --- | --- | --- | --- | --- | --- |
| UK | 111.62 | 107.69 | 103.28 | 93.59 | 88.27 | 86.36 | 87.89 |
| Rank Order Position of UK | 12 | 12 | 10 | 8 | 6 | 2 | 2 |
| Australia | 96.33 | 91.11 | 89.19 | 77.68 | 74.28 | 73.87 | 68.52 |
| Austria | 118.60 | 111.11 | 97.66 | 79.43 | 71.61 | 64.94 | 61.12 |
| Belgium | 129.66 | 120.98 | 111.67 | 96.69 | 90.54 | 79.62 | 68.46 |
| Canada | 103.23 | 99.72 | 89.52 | 87.52 | 79.26 | 75.56 | 79.50 |
| Germany | 118.37 | 109.84 | 99.62 | 85.24 | 74.67 | 68.76 | 64.39 |
| Denmark | 141.53 | 143.06 | 114.36 | 98.66 | 82.88 | 66.36 | 56.88 |
| Finland | 115.17 | 106.75 | 108.34 | 98.71 | 82.10 | 64.86 | 65.86 |
| France | 117.64 | 120.11 | 107.67 | 96.36 | 85.24 | 74.92 | 71.15 |
| Ireland | 113.87 | 108.01 | 107.45 | 89.88 | 75.73 | 65.88 | 64.20 |
| Iceland | 104.43 | 110.67 | 66.74 | 63.65 | 65.12 | 62.72 | 69.57 |
| Italy | 94.84 | 94.34 | 78.08 | 67.04 | 59.03 | 54.85 | 52.01 |
| Japan | 86.58 | 83.90 | 77.96 | 73.38 | 68.89 | 61.05 | 57.12 |
| Luxembourg | 137.43 | 109.27 | 104.35 | 74.23 | 90.37 | 56.26 | 47.68 |
| Netherlands | 103.39 | 107.31 | 108.00 | 93.44 | 76.10 | 67.53 | 62.62 |
| Norway | 105.50 | 94.63 | 90.84 | 84.41 | 69.66 | 59.45 | 52.50 |
| New Zealand | 135.38 | 118.54 | 99.75 | 96.91 | 91.58 | 83.51 | 80.78 |
| Portugal | 133.99 | 121.72 | 116.00 | 100.34 | 88.52 | 69.04 | 69.52 |
| Spain | 96.78 | 98.10 | 82.10 | 74.97 | 63.14 | 54.85 | 49.59 |
| Sweden | 102.97 | 89.35 | 78.81 | 68.57 | 59.10 | 56.13 | 56.53 |
| Switzerland | 105.07 | 102.66 | 80.34 | 74.49 | 61.89 | 51.15 | 46.45 |
| USA | 141.14 | 144.88 | 136.02 | 139.18 | 129.96 | 134.51 | 138.46 |

**Table S2. (continued)****Panel B. Men**
|  | 1990 | 1995 | 2000 | 2005 | 2010 | 2015 | 2019 |
| --- | --- | --- | --- | --- | --- | --- | --- |
| UK | 185.40 | 179.92 | 172.59 | 158.72 | 151.09 | 144.34 | 148.35 |
| Rank Order Position of UK | 18 | 16 | 14 | 8 | 6 | 5 | 2 |
| Australia | 191.74 | 182.01 | 171.98 | 150.90 | 135.95 | 135.92 | 130.07 |
| Austria | 255.95 | 235.31 | 190.42 | 157.32 | 142.20 | 120.22 | 115.05 |
| Belgium | 224.36 | 227.32 | 209.47 | 177.96 | 156.94 | 134.02 | 125.70 |
| Canada | 198.02 | 196.75 | 157.87 | 151.03 | 134.10 | 132.69 | 142.82 |
| Germany | 225.49 | 215.83 | 186.89 | 157.98 | 137.19 | 124.05 | 115.49 |
| Denmark | 247.08 | 238.32 | 200.02 | 169.05 | 146.37 | 114.29 | 102.23 |
| Finland | 329.60 | 280.25 | 251.69 | 229.61 | 191.76 | 150.44 | 144.77 |
| France | 293.29 | 293.56 | 244.87 | 209.59 | 180.99 | 158.66 | 145.27 |
| Ireland | 186.93 | 179.39 | 182.87 | 146.91 | 143.60 | 117.27 | 112.13 |
| Iceland | 171.18 | 148.44 | 156.12 | 82.83 | 127.24 | 113.61 | 117.90 |
| Italy | 202.66 | 198.71 | 158.48 | 128.47 | 108.96 | 98.76 | 90.44 |
| Japan | 159.72 | 153.44 | 150.94 | 145.19 | 129.49 | 107.41 | 96.42 |
| Luxembourg | 264.73 | 239.86 | 213.85 | 153.39 | 140.44 | 100.21 | 85.17 |
| Netherlands | 164.89 | 157.38 | 145.05 | 121.07 | 103.42 | 96.73 | 92.56 |
| Norway | 201.60 | 176.67 | 177.74 | 141.23 | 122.78 | 101.08 | 95.68 |
| New Zealand | 211.26 | 187.49 | 173.41 | 155.26 | 139.81 | 123.70 | 132.75 |
| Portugal | 304.12 | 327.68 | 305.56 | 264.64 | 210.58 | 161.72 | 138.92 |
| Spain | 251.44 | 263.99 | 205.37 | 171.82 | 128.69 | 101.32 | 90.99 |
| Sweden | 177.63 | 159.51 | 134.66 | 118.77 | 105.22 | 101.87 | 90.88 |
| Switzerland | 208.57 | 200.77 | 157.40 | 128.75 | 105.02 | 90.07 | 77.30 |
| USA | 307.75 | 316.05 | 253.62 | 250.70 | 225.51 | 233.75 | 254.82 |

**Table S3.**
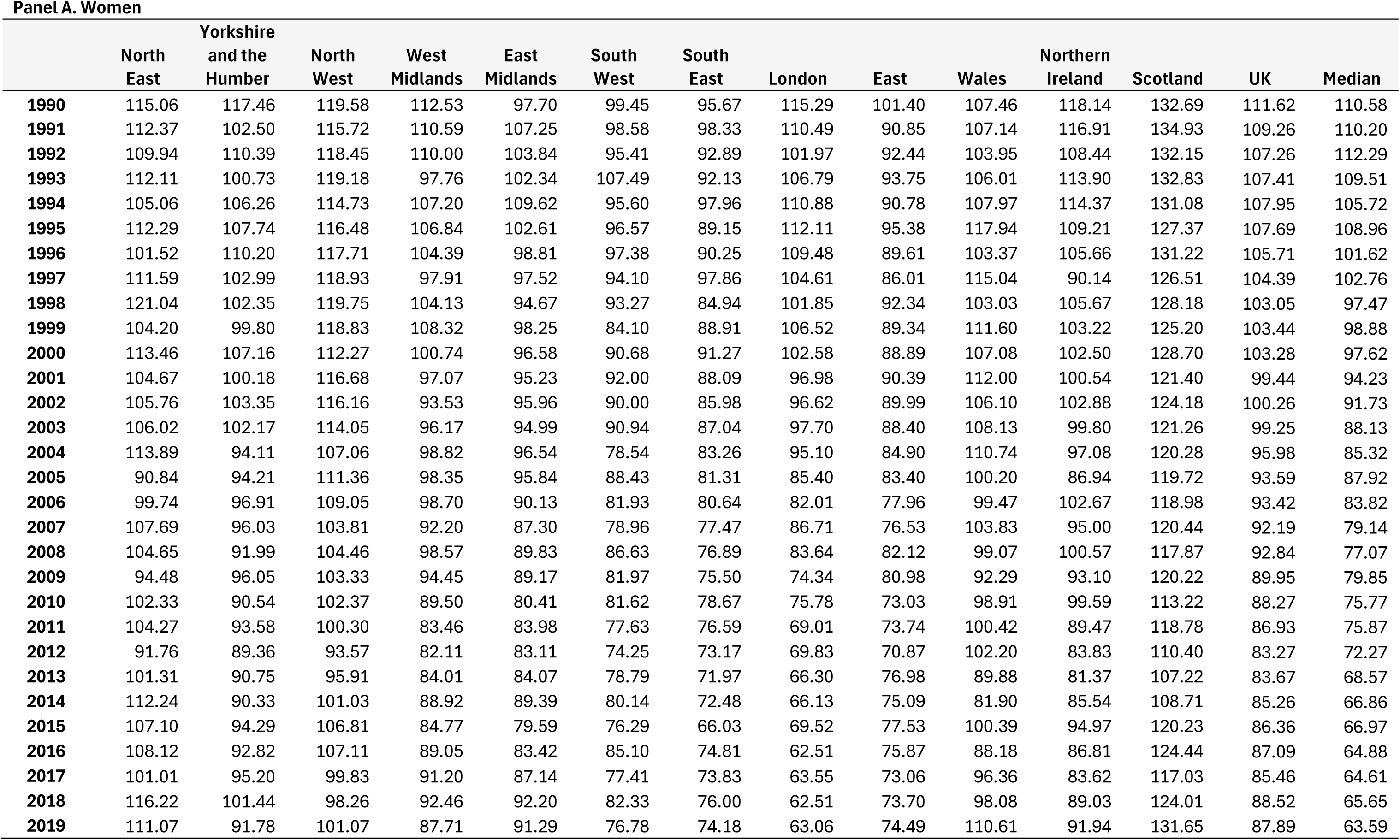
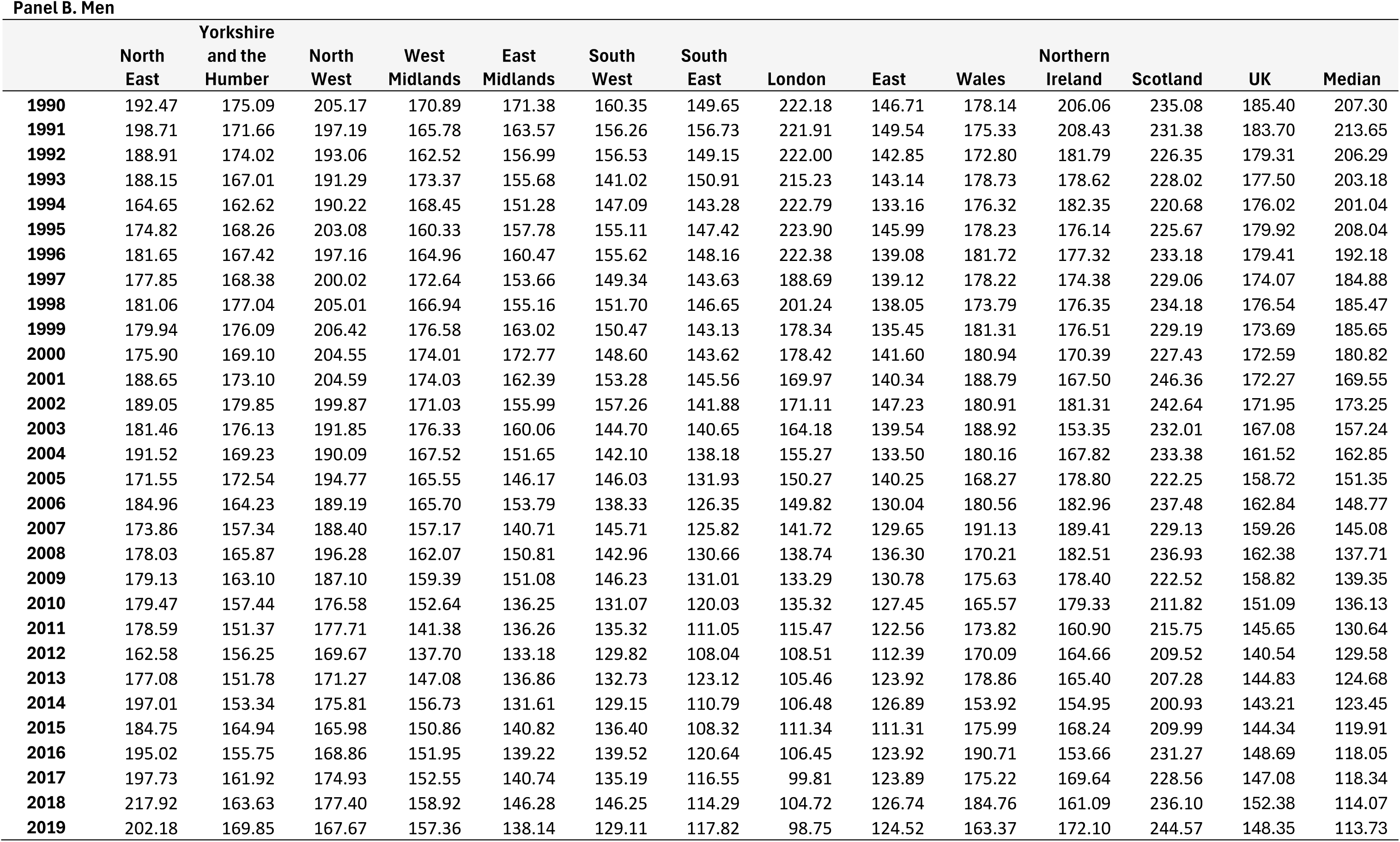
All-causes age-standardised mortality rates per 100,000 at ages 25-49 years for each area of the UK together with the UK as a whole and the comparator median of 21 peer countries by year (1990-2019) and sex.

**Panel A. Women**
|  | North<br>East | Yorkshire<br>and the<br>Humber | North<br>West | West<br>Midlands | East<br>Midlands | South<br>West | South<br>East | London | East | Wales | Northern<br>Ireland | Scotland | UK | Median |
| --- | --- | --- | --- | --- | --- | --- | --- | --- | --- | --- | --- | --- | --- | --- |
| **1990** | 115.06 | 117.46 | 119.58 | 112.53 | 97.70 | 99.45 | 95.67 | 115.29 | 101.40 | 107.46 | 118.14 | 132.69 | 111.62 | 110.58 |
| **1991** | 112.37 | 102.50 | 115.72 | 110.59 | 107.25 | 98.58 | 98.33 | 110.49 | 90.85 | 107.14 | 116.91 | 134.93 | 109.26 | 110.20 |
| **1992** | 109.94 | 110.39 | 118.45 | 110.00 | 103.84 | 95.41 | 92.89 | 101.97 | 92.44 | 103.95 | 108.44 | 132.15 | 107.26 | 112.29 |
| **1993** | 112.11 | 100.73 | 119.18 | 97.76 | 102.34 | 107.49 | 92.13 | 106.79 | 93.75 | 106.01 | 113.90 | 132.83 | 107.41 | 109.51 |
| **1994** | 105.06 | 106.26 | 114.73 | 107.20 | 109.62 | 95.60 | 97.96 | 110.88 | 90.78 | 107.97 | 114.37 | 131.08 | 107.95 | 105.72 |
| **1995** | 112.29 | 107.74 | 116.48 | 106.84 | 102.61 | 96.57 | 89.15 | 112.11 | 95.38 | 117.94 | 109.21 | 127.37 | 107.69 | 108.96 |
| **1996** | 101.52 | 110.20 | 117.71 | 104.39 | 98.81 | 97.38 | 90.25 | 109.48 | 89.61 | 103.37 | 105.66 | 131.22 | 105.71 | 101.62 |
| **1997** | 111.59 | 102.99 | 118.93 | 97.91 | 97.52 | 94.10 | 97.86 | 104.61 | 86.01 | 115.04 | 90.14 | 126.51 | 104.39 | 102.76 |
| **1998** | 121.04 | 102.35 | 119.75 | 104.13 | 94.67 | 93.27 | 84.94 | 101.85 | 92.34 | 103.03 | 105.67 | 128.18 | 103.05 | 97.47 |
| **1999** | 104.20 | 99.80 | 118.83 | 108.32 | 98.25 | 84.10 | 88.91 | 106.52 | 89.34 | 111.60 | 103.22 | 125.20 | 103.44 | 98.88 |
| **2000** | 113.46 | 107.16 | 112.27 | 100.74 | 96.58 | 90.68 | 91.27 | 102.58 | 88.89 | 107.08 | 102.50 | 128.70 | 103.28 | 97.62 |
| **2001** | 104.67 | 100.18 | 116.68 | 97.07 | 95.23 | 92.00 | 88.09 | 96.98 | 90.39 | 112.00 | 100.54 | 121.40 | 99.44 | 94.23 |
| **2002** | 105.76 | 103.35 | 116.16 | 93.53 | 95.96 | 90.00 | 85.98 | 96.62 | 89.99 | 106.10 | 102.88 | 124.18 | 100.26 | 91.73 |
| **2003** | 106.02 | 102.17 | 114.05 | 96.17 | 94.99 | 90.94 | 87.04 | 97.70 | 88.40 | 108.13 | 99.80 | 121.26 | 99.25 | 88.13 |
| **2004** | 113.89 | 94.11 | 107.06 | 98.82 | 96.54 | 78.54 | 83.26 | 95.10 | 84.90 | 110.74 | 97.08 | 120.28 | 95.98 | 85.32 |
| **2005** | 90.84 | 94.21 | 111.36 | 98.35 | 95.84 | 88.43 | 81.31 | 85.40 | 83.40 | 100.20 | 86.94 | 119.72 | 93.59 | 87.92 |
| **2006** | 99.74 | 96.91 | 109.05 | 98.70 | 90.13 | 81.93 | 80.64 | 82.01 | 77.96 | 99.47 | 102.67 | 118.98 | 93.42 | 83.82 |
| **2007** | 107.69 | 96.03 | 103.81 | 92.20 | 87.30 | 78.96 | 77.47 | 86.71 | 76.53 | 103.83 | 95.00 | 120.44 | 92.19 | 79.14 |
| **2008** | 104.65 | 91.99 | 104.46 | 98.57 | 89.83 | 86.63 | 76.89 | 83.64 | 82.12 | 99.07 | 100.57 | 117.87 | 92.84 | 77.07 |
| **2009** | 94.48 | 96.05 | 103.33 | 94.45 | 89.17 | 81.97 | 75.50 | 74.34 | 80.98 | 92.29 | 93.10 | 120.22 | 89.95 | 79.85 |
| **2010** | 102.33 | 90.54 | 102.37 | 89.50 | 80.41 | 81.62 | 78.67 | 75.78 | 73.03 | 98.91 | 99.59 | 113.22 | 88.27 | 75.77 |
| **2011** | 104.27 | 93.58 | 100.30 | 83.46 | 83.98 | 77.63 | 76.59 | 69.01 | 73.74 | 100.42 | 89.47 | 118.78 | 86.93 | 75.87 |
| **2012** | 91.76 | 89.36 | 93.57 | 82.11 | 83.11 | 74.25 | 73.17 | 69.83 | 70.87 | 102.20 | 83.83 | 110.40 | 83.27 | 72.27 |
| **2013** | 101.31 | 90.75 | 95.91 | 84.01 | 84.07 | 78.79 | 71.97 | 66.30 | 76.98 | 89.88 | 81.37 | 107.22 | 83.67 | 68.57 |
| **2014** | 112.24 | 90.33 | 101.03 | 88.92 | 89.39 | 80.14 | 72.48 | 66.13 | 75.09 | 81.90 | 85.54 | 108.71 | 85.26 | 66.86 |
| **2015** | 107.10 | 94.29 | 106.81 | 84.77 | 79.59 | 76.29 | 66.03 | 69.52 | 77.53 | 100.39 | 94.97 | 120.23 | 86.36 | 66.97 |
| **2016** | 108.12 | 92.82 | 107.11 | 89.05 | 83.42 | 85.10 | 74.81 | 62.51 | 75.87 | 88.18 | 86.81 | 124.44 | 87.09 | 64.88 |
| **2017** | 101.01 | 95.20 | 99.83 | 91.20 | 87.14 | 77.41 | 73.83 | 63.55 | 73.06 | 96.36 | 83.62 | 117.03 | 85.46 | 64.61 |
| **2018** | 116.22 | 101.44 | 98.26 | 92.46 | 92.20 | 82.33 | 76.00 | 62.51 | 73.70 | 98.08 | 89.03 | 124.01 | 88.52 | 65.65 |
| **2019** | 111.07 | 91.78 | 101.07 | 87.71 | 91.29 | 76.78 | 74.18 | 63.06 | 74.49 | 110.61 | 91.94 | 131.65 | 87.89 | 63.59 |

**Table S3. (continued)** **Panel B. Men**
|  | North<br>East | Yorkshire<br>and the<br>Humber | North<br>West | West<br>Midlands | East<br>Midlands | South<br>West | South<br>East | London | East | Wales | Northern<br>Ireland | Scotland | UK | Median |
| --- | --- | --- | --- | --- | --- | --- | --- | --- | --- | --- | --- | --- | --- | --- |
| **1990** | 192.47 | 175.09 | 205.17 | 170.89 | 171.38 | 160.35 | 149.65 | 222.18 | 146.71 | 178.14 | 206.06 | 235.08 | 185.40 | 207.30 |
| **1991** | 198.71 | 171.66 | 197.19 | 165.78 | 163.57 | 156.26 | 156.73 | 221.91 | 149.54 | 175.33 | 208.43 | 231.38 | 183.70 | 213.65 |
| **1992** | 188.91 | 174.02 | 193.06 | 162.52 | 156.99 | 156.53 | 149.15 | 222.00 | 142.85 | 172.80 | 181.79 | 226.35 | 179.31 | 206.29 |
| **1993** | 188.15 | 167.01 | 191.29 | 173.37 | 155.68 | 141.02 | 150.91 | 215.23 | 143.14 | 178.73 | 178.62 | 228.02 | 177.50 | 203.18 |
| **1994** | 164.65 | 162.62 | 190.22 | 168.45 | 151.28 | 147.09 | 143.28 | 222.79 | 133.16 | 176.32 | 182.35 | 220.68 | 176.02 | 201.04 |
| **1995** | 174.82 | 168.26 | 203.08 | 160.33 | 157.78 | 155.11 | 147.42 | 223.90 | 145.99 | 178.23 | 176.14 | 225.67 | 179.92 | 208.04 |
| **1996** | 181.65 | 167.42 | 197.16 | 164.96 | 160.47 | 155.62 | 148.16 | 222.38 | 139.08 | 181.72 | 177.32 | 233.18 | 179.41 | 192.18 |
| **1997** | 177.85 | 168.38 | 200.02 | 172.64 | 153.66 | 149.34 | 143.63 | 188.69 | 139.12 | 178.22 | 174.38 | 229.06 | 174.07 | 184.88 |
| **1998** | 181.06 | 177.04 | 205.01 | 166.94 | 155.16 | 151.70 | 146.65 | 201.24 | 138.05 | 173.79 | 176.35 | 234.18 | 176.54 | 185.47 |
| **1999** | 179.94 | 176.09 | 206.42 | 176.58 | 163.02 | 150.47 | 143.13 | 178.34 | 135.45 | 181.31 | 176.51 | 229.19 | 173.69 | 185.65 |
| **2000** | 175.90 | 169.10 | 204.55 | 174.01 | 172.77 | 148.60 | 143.62 | 178.42 | 141.60 | 180.94 | 170.39 | 227.43 | 172.59 | 180.82 |
| **2001** | 188.65 | 173.10 | 204.59 | 174.03 | 162.39 | 153.28 | 145.56 | 169.97 | 140.34 | 188.79 | 167.50 | 246.36 | 172.27 | 169.55 |
| **2002** | 189.05 | 179.85 | 199.87 | 171.03 | 155.99 | 157.26 | 141.88 | 171.11 | 147.23 | 180.91 | 181.31 | 242.64 | 171.95 | 173.25 |
| **2003** | 181.46 | 176.13 | 191.85 | 176.33 | 160.06 | 144.70 | 140.65 | 164.18 | 139.54 | 188.92 | 153.35 | 232.01 | 167.08 | 157.24 |
| **2004** | 191.52 | 169.23 | 190.09 | 167.52 | 151.65 | 142.10 | 138.18 | 155.27 | 133.50 | 180.16 | 167.82 | 233.38 | 161.52 | 162.85 |
| **2005** | 171.55 | 172.54 | 194.77 | 165.55 | 146.17 | 146.03 | 131.93 | 150.27 | 140.25 | 168.27 | 178.80 | 222.25 | 158.72 | 151.35 |
| **2006** | 184.96 | 164.23 | 189.19 | 165.70 | 153.79 | 138.33 | 126.35 | 149.82 | 130.04 | 180.56 | 182.96 | 237.48 | 162.84 | 148.77 |
| **2007** | 173.86 | 157.34 | 188.40 | 157.17 | 140.71 | 145.71 | 125.82 | 141.72 | 129.65 | 191.13 | 189.41 | 229.13 | 159.26 | 145.08 |
| **2008** | 178.03 | 165.87 | 196.28 | 162.07 | 150.81 | 142.96 | 130.66 | 138.74 | 136.30 | 170.21 | 182.51 | 236.93 | 162.38 | 137.71 |
| **2009** | 179.13 | 163.10 | 187.10 | 159.39 | 151.08 | 146.23 | 131.01 | 133.29 | 130.78 | 175.63 | 178.40 | 222.52 | 158.82 | 139.35 |
| **2010** | 179.47 | 157.44 | 176.58 | 152.64 | 136.25 | 131.07 | 120.03 | 135.32 | 127.45 | 165.57 | 179.33 | 211.82 | 151.09 | 136.13 |
| **2011** | 178.59 | 151.37 | 177.71 | 141.38 | 136.26 | 135.32 | 111.05 | 115.47 | 122.56 | 173.82 | 160.90 | 215.75 | 145.65 | 130.64 |
| **2012** | 162.58 | 156.25 | 169.67 | 137.70 | 133.18 | 129.82 | 108.04 | 108.51 | 112.39 | 170.09 | 164.66 | 209.52 | 140.54 | 129.58 |
| **2013** | 177.08 | 151.78 | 171.27 | 147.08 | 136.86 | 132.73 | 123.12 | 105.46 | 123.92 | 178.86 | 165.40 | 207.28 | 144.83 | 124.68 |
| **2014** | 197.01 | 153.34 | 175.81 | 156.73 | 131.61 | 129.15 | 110.79 | 106.48 | 126.89 | 153.92 | 154.95 | 200.93 | 143.21 | 123.45 |
| **2015** | 184.75 | 164.94 | 165.98 | 150.86 | 140.82 | 136.40 | 108.32 | 111.34 | 111.31 | 175.99 | 168.24 | 209.99 | 144.34 | 119.91 |
| **2016** | 195.02 | 155.75 | 168.86 | 151.95 | 139.22 | 139.52 | 120.64 | 106.45 | 123.92 | 190.71 | 153.66 | 231.27 | 148.69 | 118.05 |
| **2017** | 197.73 | 161.92 | 174.93 | 152.55 | 140.74 | 135.19 | 116.55 | 99.81 | 123.89 | 175.22 | 169.64 | 228.56 | 147.08 | 118.34 |
| **2018** | 217.92 | 163.63 | 177.40 | 158.92 | 146.28 | 146.25 | 114.29 | 104.72 | 126.74 | 184.76 | 161.09 | 236.10 | 152.38 | 114.07 |
| **2019** | 202.18 | 169.85 | 167.67 | 157.36 | 138.14 | 129.11 | 117.82 | 98.75 | 124.52 | 163.37 | 172.10 | 244.57 | 148.35 | 113.73 |

**Table S4.**
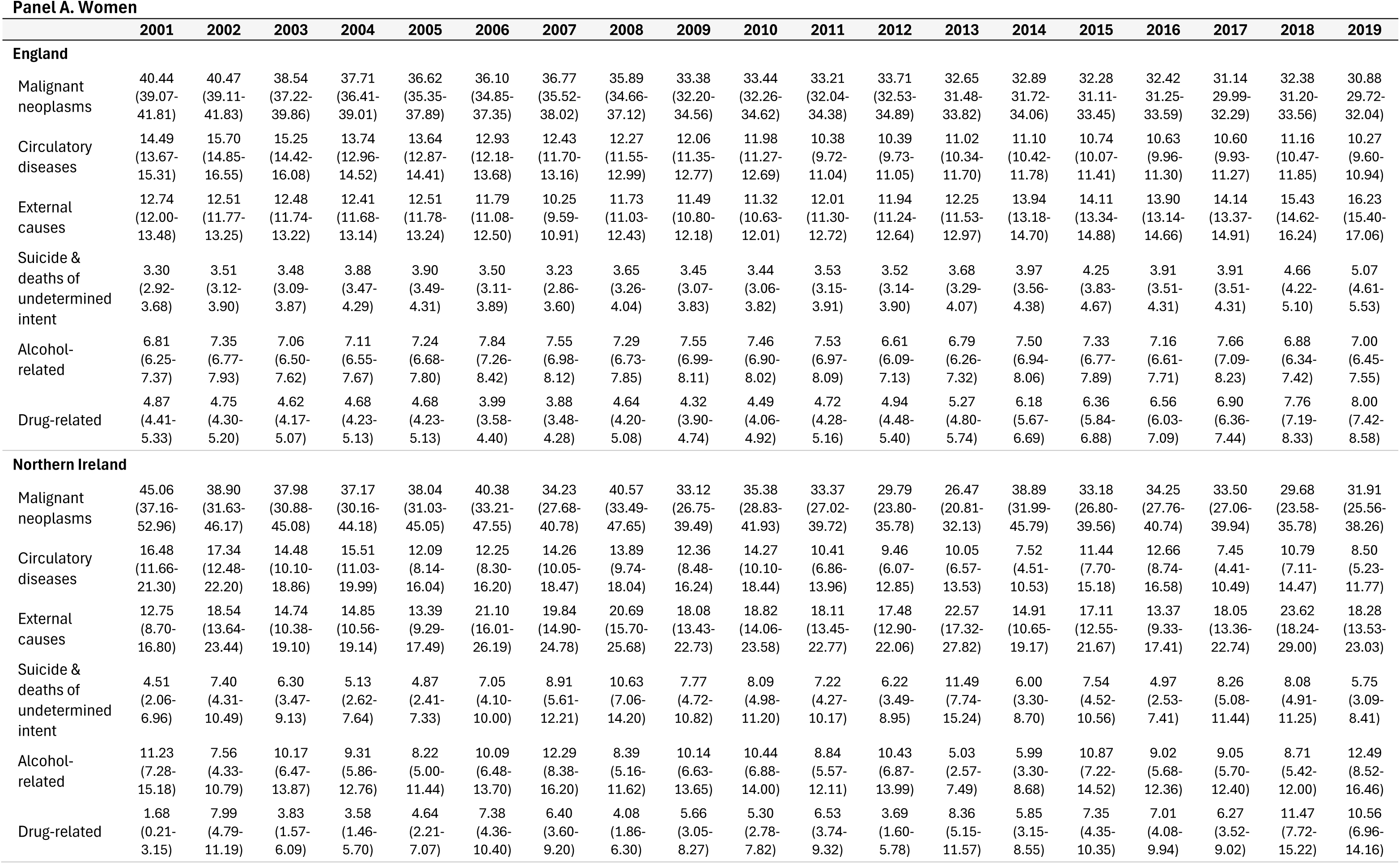
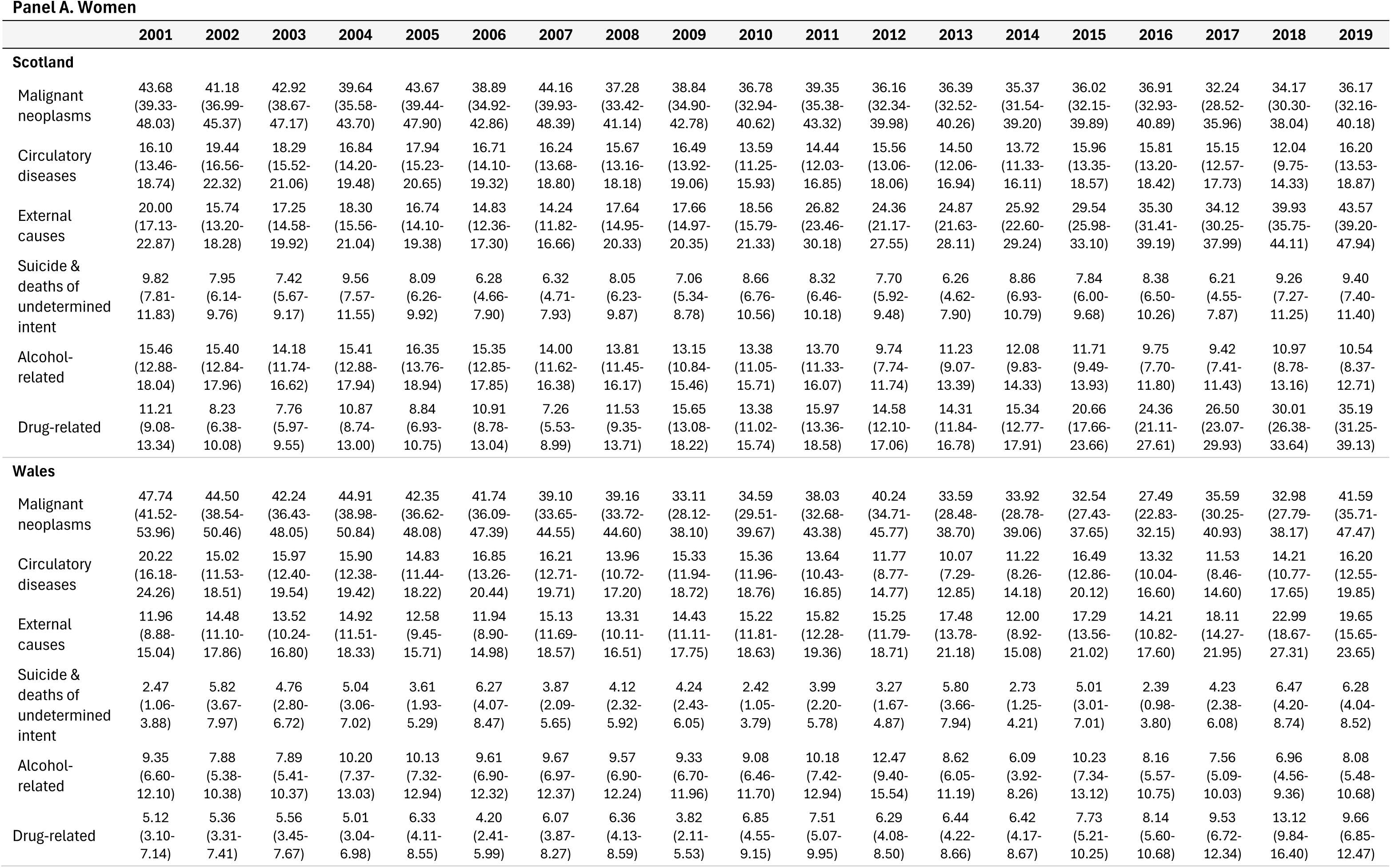
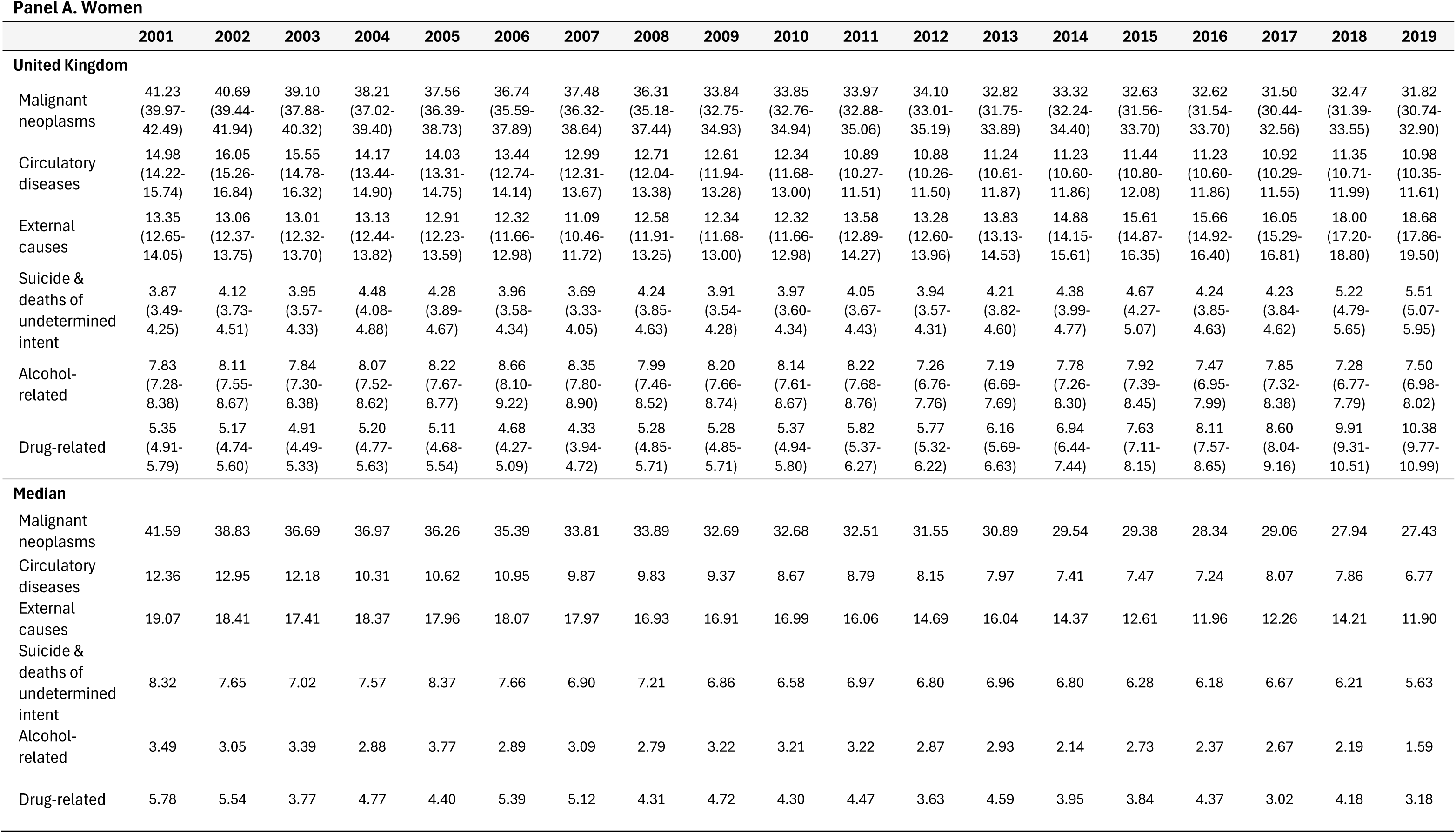
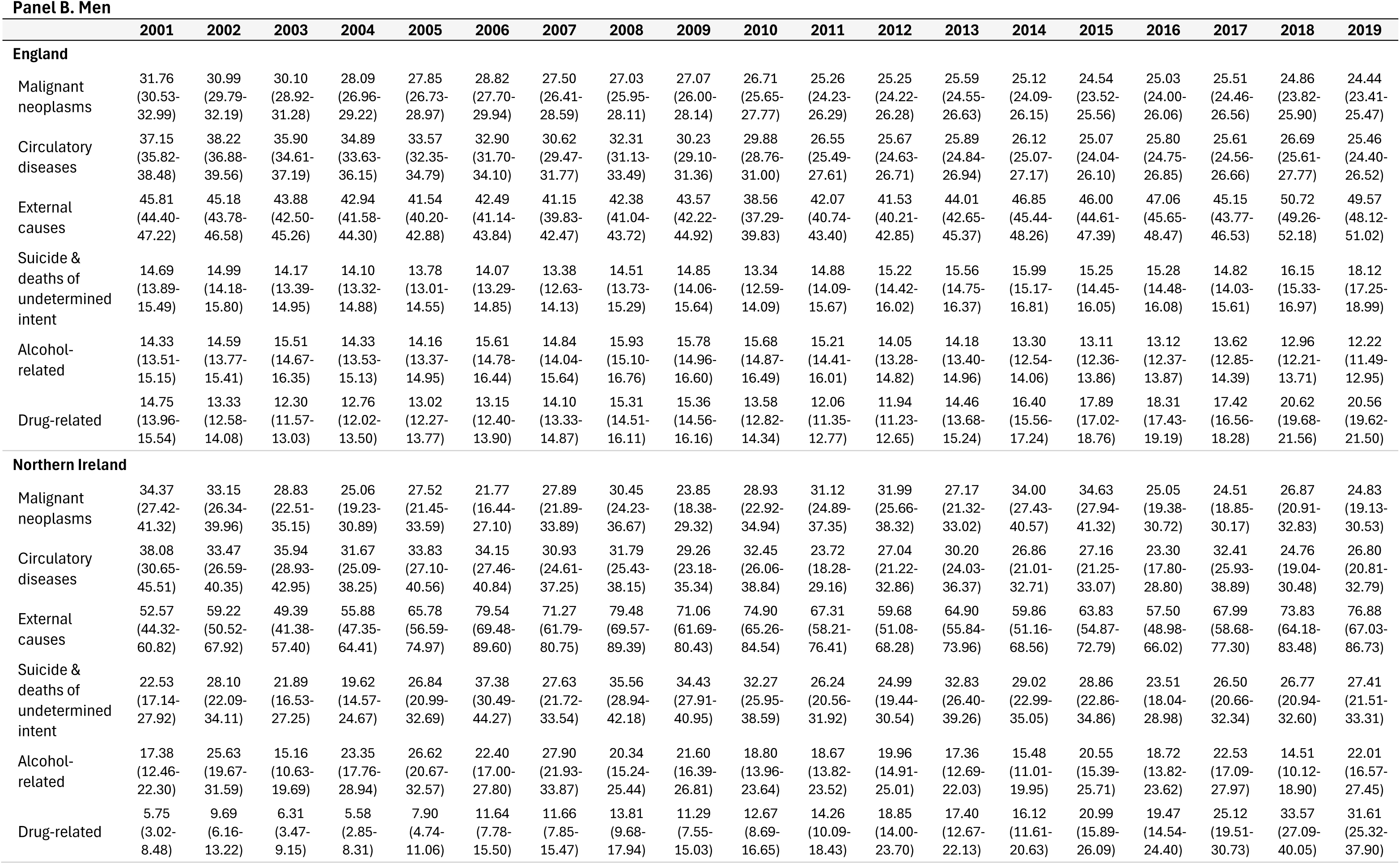
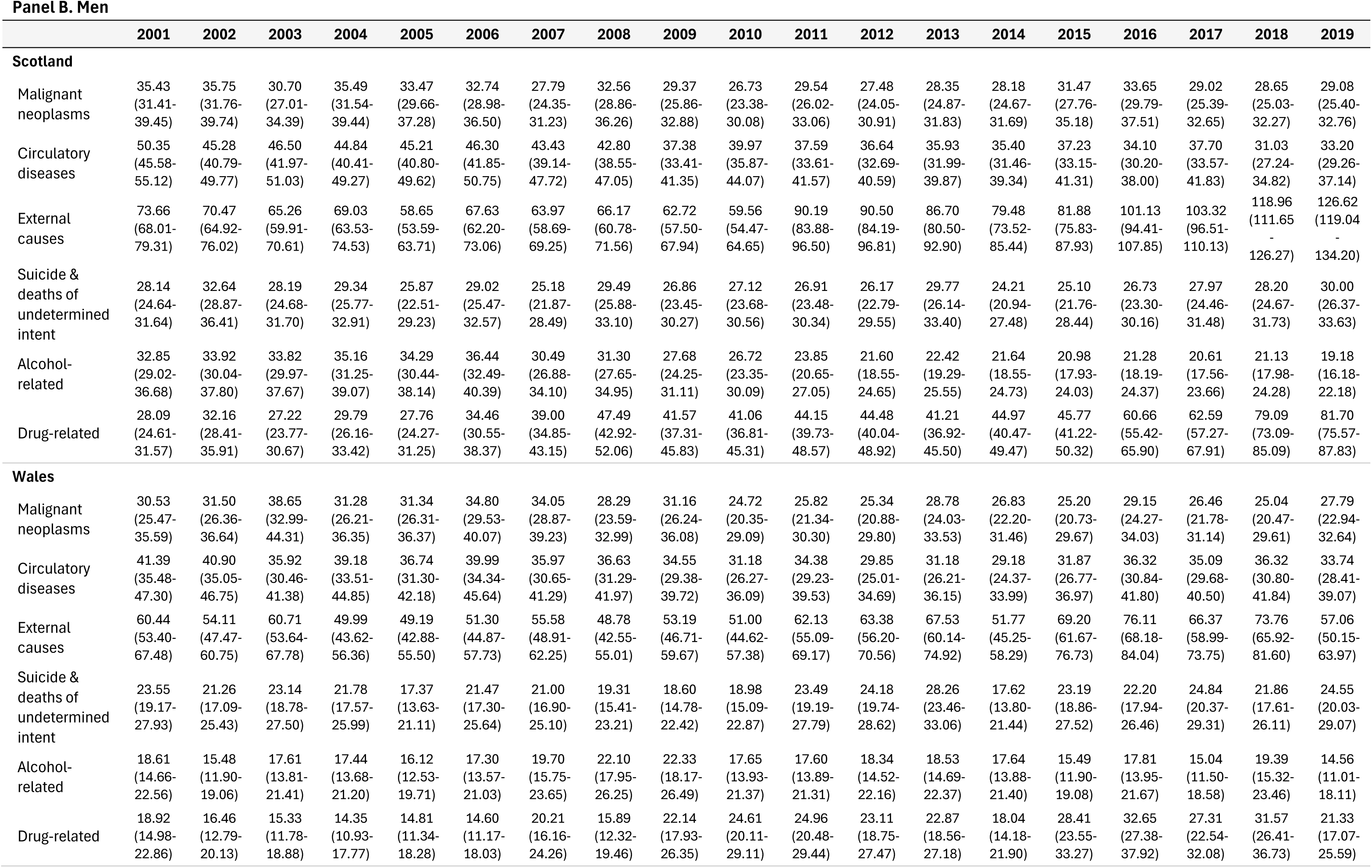
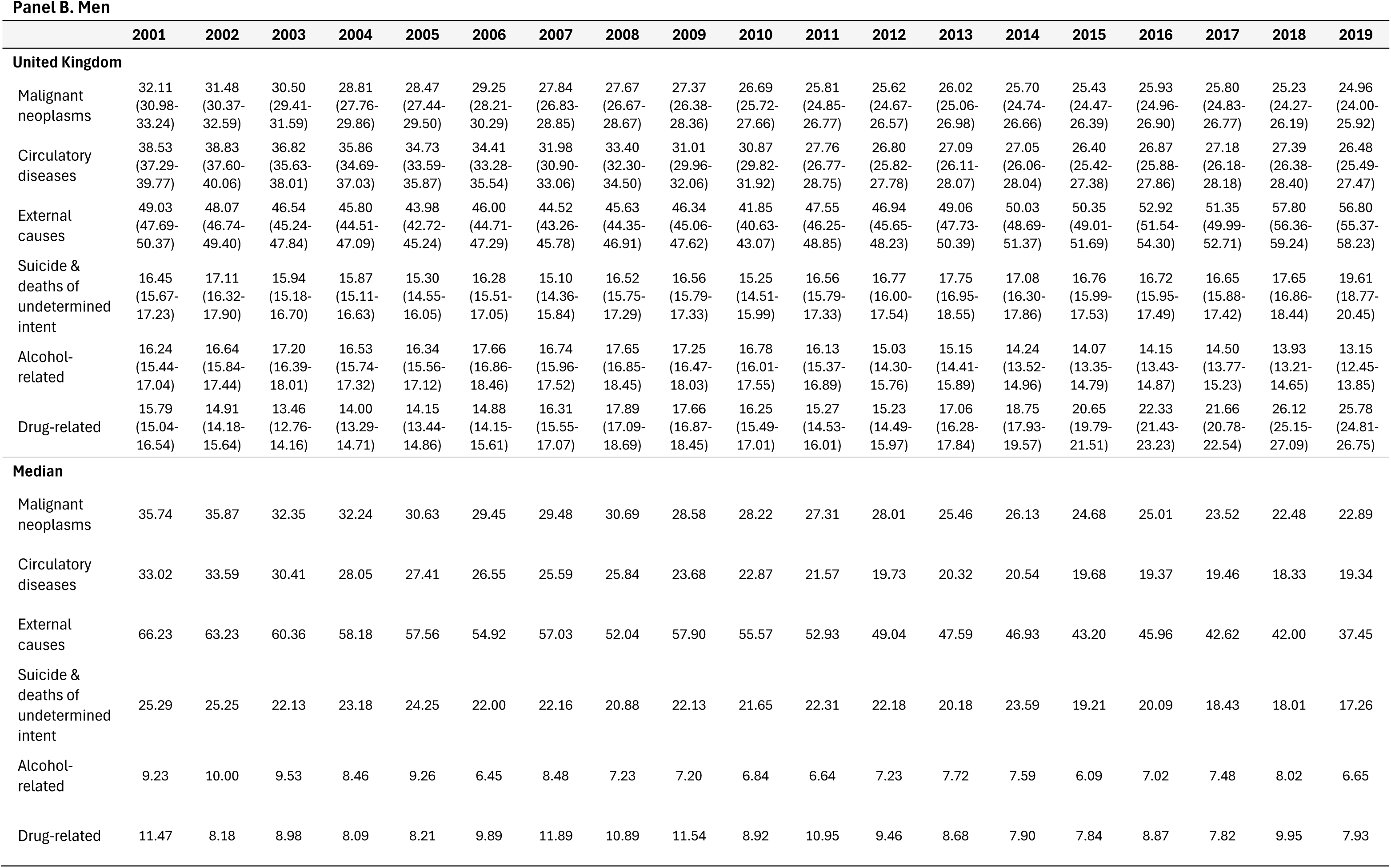
Cause-specific age-standardised mortality rates per 100,000 with 95% confidence intervals at ages 25-49 by year (2001-2021) for females (A) and males (B) in England, Wales, Northern Ireland, Scotland, and the UK, and the comparator median for the 21 peer countries.

**Table S5.**
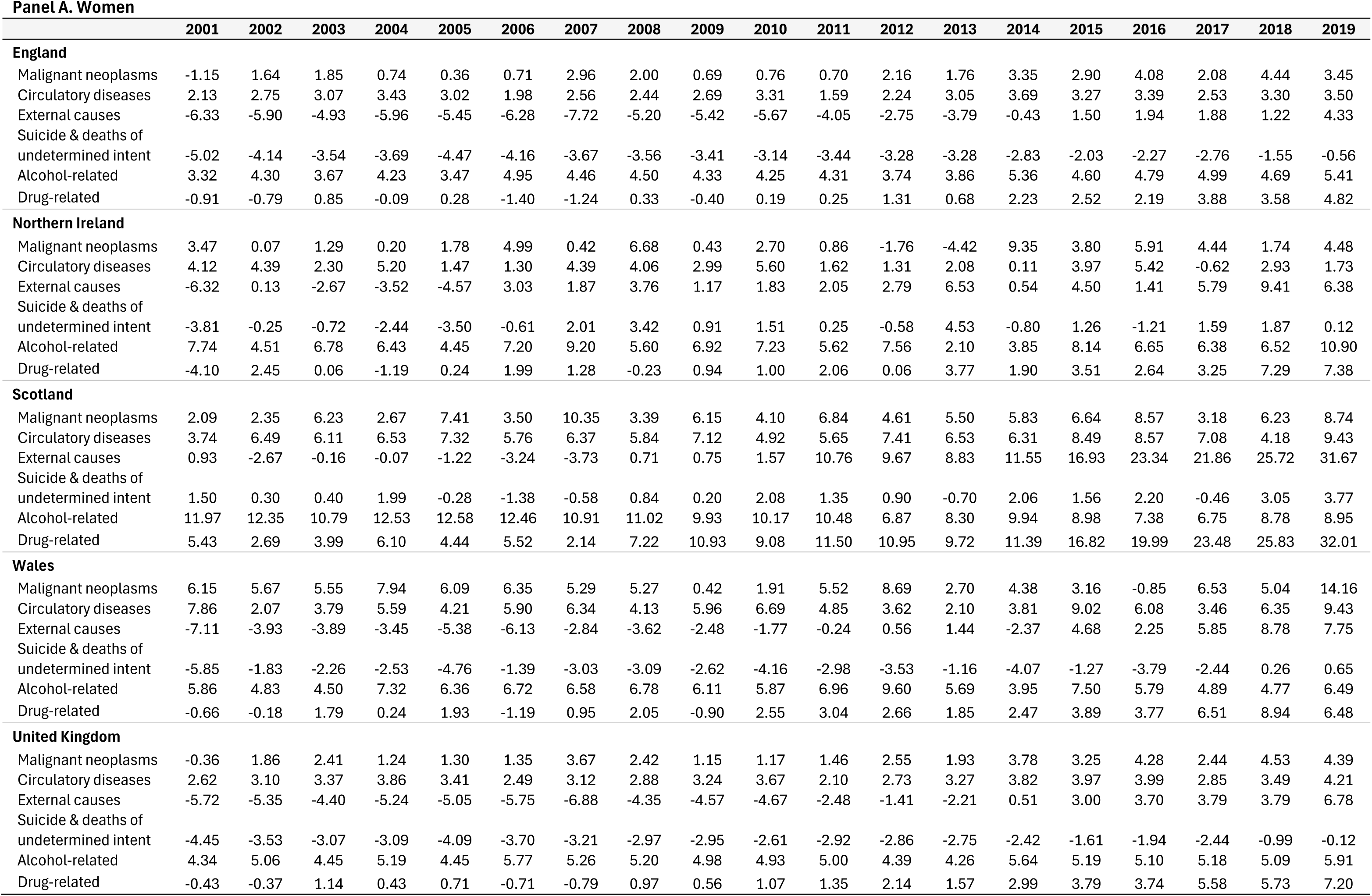
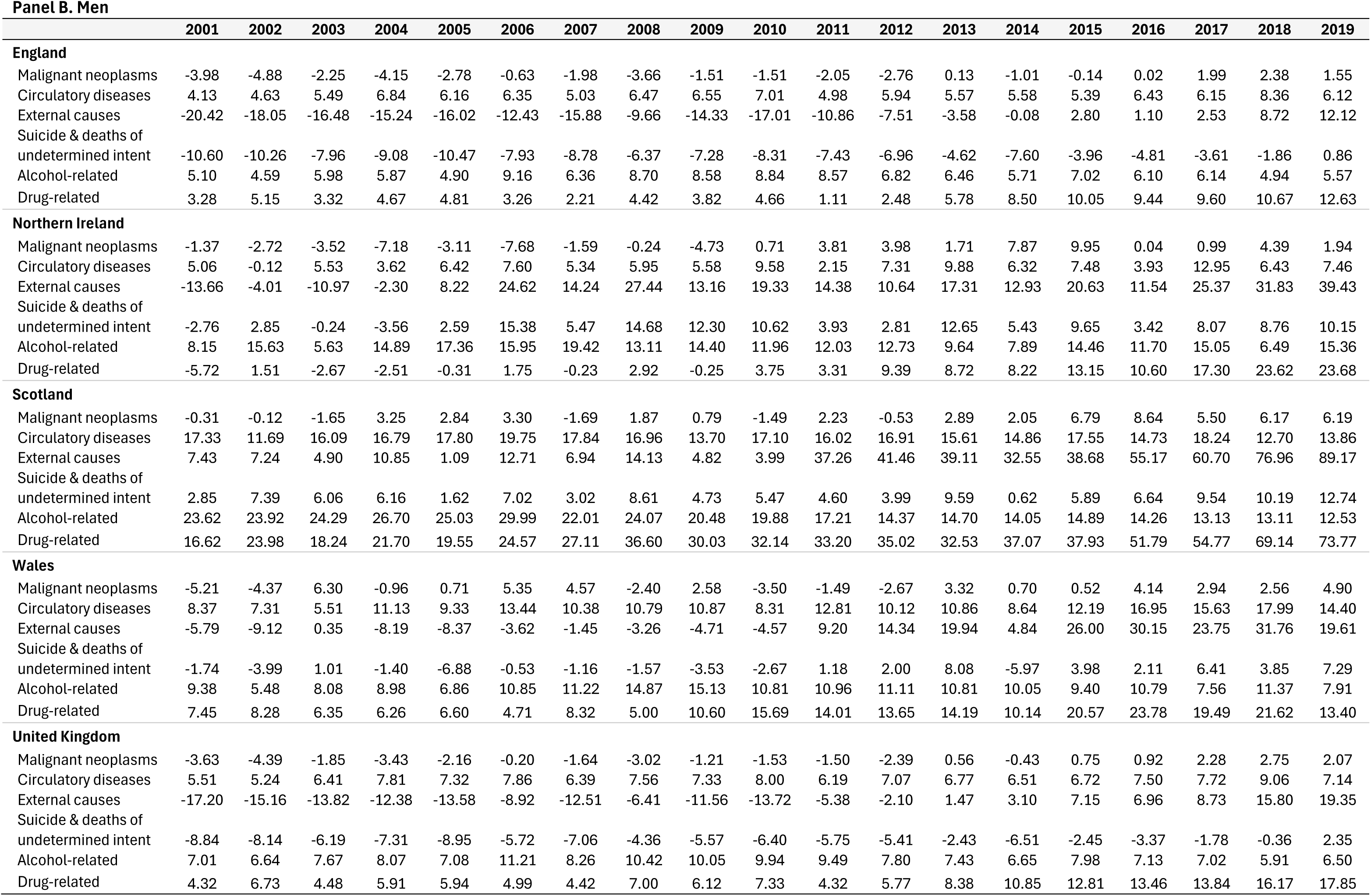
Difference in mortality rates per 100,000 at ages 25-49 years between the four constituent parts of the UK and the comparator median for 21 peer countries for selected causes by year (2001-19) and sex.

**Panel A. Women**
|  | 2001 | 2002 | 2003 | 2004 | 2005 | 2006 | 2007 | 2008 | 2009 | 2010 | 2011 | 2012 | 2013 | 2014 | 2015 | 2016 | 2017 | 2018 | 2019 |
| --- | --- | --- | --- | --- | --- | --- | --- | --- | --- | --- | --- | --- | --- | --- | --- | --- | --- | --- | --- |
| **England** |  |  |  |  |  |  |  |  |  |  |  |  |  |  |  |  |  |  |  |
| Malignant neoplasms | -1.15 | 1.64 | 1.85 | 0.74 | 0.36 | 0.71 | 2.96 | 2.00 | 0.69 | 0.76 | 0.70 | 2.16 | 1.76 | 3.35 | 2.90 | 4.08 | 2.08 | 4.44 | 3.45 |
| Circulatory diseases | 2.13 | 2.75 | 3.07 | 3.43 | 3.02 | 1.98 | 2.56 | 2.44 | 2.69 | 3.31 | 1.59 | 2.24 | 3.05 | 3.69 | 3.27 | 3.39 | 2.53 | 3.30 | 3.50 |
| External causes | -6.33 | -5.90 | -4.93 | -5.96 | -5.45 | -6.28 | -7.72 | -5.20 | -5.42 | -5.67 | -4.05 | -2.75 | -3.79 | -0.43 | 1.50 | 1.94 | 1.88 | 1.22 | 4.33 |
| Suicide & deaths of undetermined intent | -5.02 | -4.14 | -3.54 | -3.69 | -4.47 | -4.16 | -3.67 | -3.56 | -3.41 | -3.14 | -3.44 | -3.28 | -3.28 | -2.83 | -2.03 | -2.27 | -2.76 | -1.55 | -0.56 |
| Alcohol-related | 3.32 | 4.30 | 3.67 | 4.23 | 3.47 | 4.95 | 4.46 | 4.50 | 4.33 | 4.25 | 4.31 | 3.74 | 3.86 | 5.36 | 4.60 | 4.79 | 4.99 | 4.69 | 5.41 |
| Drug-related | -0.91 | -0.79 | 0.85 | -0.09 | 0.28 | -1.40 | -1.24 | 0.33 | -0.40 | 0.19 | 0.25 | 1.31 | 0.68 | 2.23 | 2.52 | 2.19 | 3.88 | 3.58 | 4.82 |
| **Northern Ireland** |  |  |  |  |  |  |  |  |  |  |  |  |  |  |  |  |  |  |  |
| Malignant neoplasms | 3.47 | 0.07 | 1.29 | 0.20 | 1.78 | 4.99 | 0.42 | 6.68 | 0.43 | 2.70 | 0.86 | -1.76 | -4.42 | 9.35 | 3.80 | 5.91 | 4.44 | 1.74 | 4.48 |
| Circulatory diseases | 4.12 | 4.39 | 2.30 | 5.20 | 1.47 | 1.30 | 4.39 | 4.06 | 2.99 | 5.60 | 1.62 | 1.31 | 2.08 | 0.11 | 3.97 | 5.42 | -0.62 | 2.93 | 1.73 |
| External causes | -6.32 | 0.13 | -2.67 | -3.52 | -4.57 | 3.03 | 1.87 | 3.76 | 1.17 | 1.83 | 2.05 | 2.79 | 6.53 | 0.54 | 4.50 | 1.41 | 5.79 | 9.41 | 6.38 |
| Suicide & deaths of undetermined intent | -3.81 | -0.25 | -0.72 | -2.44 | -3.50 | -0.61 | 2.01 | 3.42 | 0.91 | 1.51 | 0.25 | -0.58 | 4.53 | -0.80 | 1.26 | -1.21 | 1.59 | 1.87 | 0.12 |
| Alcohol-related | 7.74 | 4.51 | 6.78 | 6.43 | 4.45 | 7.20 | 9.20 | 5.60 | 6.92 | 7.23 | 5.62 | 7.56 | 2.10 | 3.85 | 8.14 | 6.65 | 6.38 | 6.52 | 10.90 |
| Drug-related | -4.10 | 2.45 | 0.06 | -1.19 | 0.24 | 1.99 | 1.28 | -0.23 | 0.94 | 1.00 | 2.06 | 0.06 | 3.77 | 1.90 | 3.51 | 2.64 | 3.25 | 7.29 | 7.38 |
| **Scotland** |  |  |  |  |  |  |  |  |  |  |  |  |  |  |  |  |  |  |  |
| Malignant neoplasms | 2.09 | 2.35 | 6.23 | 2.67 | 7.41 | 3.50 | 10.35 | 3.39 | 6.15 | 4.10 | 6.84 | 4.61 | 5.50 | 5.83 | 6.64 | 8.57 | 3.18 | 6.23 | 8.74 |
| Circulatory diseases | 3.74 | 6.49 | 6.11 | 6.53 | 7.32 | 5.76 | 6.37 | 5.84 | 7.12 | 4.92 | 5.65 | 7.41 | 6.53 | 6.31 | 8.49 | 8.57 | 7.08 | 4.18 | 9.43 |
| External causes | 0.93 | -2.67 | -0.16 | -0.07 | -1.22 | -3.24 | -3.73 | 0.71 | 0.75 | 1.57 | 10.76 | 9.67 | 8.83 | 11.55 | 16.93 | 23.34 | 21.86 | 25.72 | 31.67 |
| Suicide & deaths of undetermined intent | 1.50 | 0.30 | 0.40 | 1.99 | -0.28 | -1.38 | -0.58 | 0.84 | 0.20 | 2.08 | 1.35 | 0.90 | -0.70 | 2.06 | 1.56 | 2.20 | -0.46 | 3.05 | 3.77 |
| Alcohol-related | 11.97 | 12.35 | 10.79 | 12.53 | 12.58 | 12.46 | 10.91 | 11.02 | 9.93 | 10.17 | 10.48 | 6.87 | 8.30 | 9.94 | 8.98 | 7.38 | 6.75 | 8.78 | 8.95 |
| Drug-related | 5.43 | 2.69 | 3.99 | 6.10 | 4.44 | 5.52 | 2.14 | 7.22 | 10.93 | 9.08 | 11.50 | 10.95 | 9.72 | 11.39 | 16.82 | 19.99 | 23.48 | 25.83 | 32.01 |
| **Wales** |  |  |  |  |  |  |  |  |  |  |  |  |  |  |  |  |  |  |  |
| Malignant neoplasms | 6.15 | 5.67 | 5.55 | 7.94 | 6.09 | 6.35 | 5.29 | 5.27 | 0.42 | 1.91 | 5.52 | 8.69 | 2.70 | 4.38 | 3.16 | -0.85 | 6.53 | 5.04 | 14.16 |
| Circulatory diseases | 7.86 | 2.07 | 3.79 | 5.59 | 4.21 | 5.90 | 6.34 | 4.13 | 5.96 | 6.69 | 4.85 | 3.62 | 2.10 | 3.81 | 9.02 | 6.08 | 3.46 | 6.35 | 9.43 |
| External causes | -7.11 | -3.93 | -3.89 | -3.45 | -5.38 | -6.13 | -2.84 | -3.62 | -2.48 | -1.77 | -0.24 | 0.56 | 1.44 | -2.37 | 4.68 | 2.25 | 5.85 | 8.78 | 7.75 |
| Suicide & deaths of undetermined intent | -5.85 | -1.83 | -2.26 | -2.53 | -4.76 | -1.39 | -3.03 | -3.09 | -2.62 | -4.16 | -2.98 | -3.53 | -1.16 | -4.07 | -1.27 | -3.79 | -2.44 | 0.26 | 0.65 |
| Alcohol-related | 5.86 | 4.83 | 4.50 | 7.32 | 6.36 | 6.72 | 6.58 | 6.78 | 6.11 | 5.87 | 6.96 | 9.60 | 5.69 | 3.95 | 7.50 | 5.79 | 4.89 | 4.77 | 6.49 |
| Drug-related | -0.66 | -0.18 | 1.79 | 0.24 | 1.93 | -1.19 | 0.95 | 2.05 | -0.90 | 2.55 | 3.04 | 2.66 | 1.85 | 2.47 | 3.89 | 3.77 | 6.51 | 8.94 | 6.48 |
| **United Kingdom** |  |  |  |  |  |  |  |  |  |  |  |  |  |  |  |  |  |  |  |
| Malignant neoplasms | -0.36 | 1.86 | 2.41 | 1.24 | 1.30 | 1.35 | 3.67 | 2.42 | 1.15 | 1.17 | 1.46 | 2.55 | 1.93 | 3.78 | 3.25 | 4.28 | 2.44 | 4.53 | 4.39 |
| Circulatory diseases | 2.62 | 3.10 | 3.37 | 3.86 | 3.41 | 2.49 | 3.12 | 2.88 | 3.24 | 3.67 | 2.10 | 2.73 | 3.27 | 3.82 | 3.97 | 3.99 | 2.85 | 3.49 | 4.21 |
| External causes | -5.72 | -5.35 | -4.40 | -5.24 | -5.05 | -5.75 | -6.88 | -4.35 | -4.57 | -4.67 | -2.48 | -1.41 | -2.21 | 0.51 | 3.00 | 3.70 | 3.79 | 3.79 | 6.78 |
| Suicide & deaths of undetermined intent | -4.45 | -3.53 | -3.07 | -3.09 | -4.09 | -3.70 | -3.21 | -2.97 | -2.95 | -2.61 | -2.92 | -2.86 | -2.75 | -2.42 | -1.61 | -1.94 | -2.44 | -0.99 | -0.12 |
| Alcohol-related | 4.34 | 5.06 | 4.45 | 5.19 | 4.45 | 5.77 | 5.26 | 5.20 | 4.98 | 4.93 | 5.00 | 4.39 | 4.26 | 5.64 | 5.19 | 5.10 | 5.18 | 5.09 | 5.91 |
| Drug-related | -0.43 | -0.37 | 1.14 | 0.43 | 0.71 | -0.71 | -0.79 | 0.97 | 0.56 | 1.07 | 1.35 | 2.14 | 1.57 | 2.99 | 3.79 | 3.74 | 5.58 | 5.73 | 7.20 |

**Table S5. (continued)** **Panel B. Men**
|  | 2001 | 2002 | 2003 | 2004 | 2005 | 2006 | 2007 | 2008 | 2009 | 2010 | 2011 | 2012 | 2013 | 2014 | 2015 | 2016 | 2017 | 2018 | 2019 |
| --- | --- | --- | --- | --- | --- | --- | --- | --- | --- | --- | --- | --- | --- | --- | --- | --- | --- | --- | --- |
| **England** |  |  |  |  |  |  |  |  |  |  |  |  |  |  |  |  |  |  |  |
| Malignant neoplasms | -3.98 | -4.88 | -2.25 | -4.15 | -2.78 | -0.63 | -1.98 | -3.66 | -1.51 | -1.51 | -2.05 | -2.76 | 0.13 | -1.01 | -0.14 | 0.02 | 1.99 | 2.38 | 1.55 |
| Circulatory diseases | 4.13 | 4.63 | 5.49 | 6.84 | 6.16 | 6.35 | 5.03 | 6.47 | 6.55 | 7.01 | 4.98 | 5.94 | 5.57 | 5.58 | 5.39 | 6.43 | 6.15 | 8.36 | 6.12 |
| External causes | -20.42 | -18.05 | -16.48 | -15.24 | -16.02 | -12.43 | -15.88 | -9.66 | -14.33 | -17.01 | -10.86 | -7.51 | -3.58 | -0.08 | 2.80 | 1.10 | 2.53 | 8.72 | 12.12 |
| Suicide & deaths of<br>undetermined intent | -10.60 | -10.26 | -7.96 | -9.08 | -10.47 | -7.93 | -8.78 | -6.37 | -7.28 | -8.31 | -7.43 | -6.96 | -4.62 | -7.60 | -3.96 | -4.81 | -3.61 | -1.86 | 0.86 |
| Alcohol-related | 5.10 | 4.59 | 5.98 | 5.87 | 4.90 | 9.16 | 6.36 | 8.70 | 8.58 | 8.84 | 8.57 | 6.82 | 6.46 | 5.71 | 7.02 | 6.10 | 6.14 | 4.94 | 5.57 |
| Drug-related | 3.28 | 5.15 | 3.32 | 4.67 | 4.81 | 3.26 | 2.21 | 4.42 | 3.82 | 4.66 | 1.11 | 2.48 | 5.78 | 8.50 | 10.05 | 9.44 | 9.60 | 10.67 | 12.63 |
| **Northern Ireland** |  |  |  |  |  |  |  |  |  |  |  |  |  |  |  |  |  |  |  |
| Malignant neoplasms | -1.37 | -2.72 | -3.52 | -7.18 | -3.11 | -7.68 | -1.59 | -0.24 | -4.73 | 0.71 | 3.81 | 3.98 | 1.71 | 7.87 | 9.95 | 0.04 | 0.99 | 4.39 | 1.94 |
| Circulatory diseases | 5.06 | -0.12 | 5.53 | 3.62 | 6.42 | 7.60 | 5.34 | 5.95 | 5.58 | 9.58 | 2.15 | 7.31 | 9.88 | 6.32 | 7.48 | 3.93 | 12.95 | 6.43 | 7.46 |
| External causes | -13.66 | -4.01 | -10.97 | -2.30 | 8.22 | 24.62 | 14.24 | 27.44 | 13.16 | 19.33 | 14.38 | 10.64 | 17.31 | 12.93 | 20.63 | 11.54 | 25.37 | 31.83 | 39.43 |
| Suicide & deaths of<br>undetermined intent | -2.76 | 2.85 | -0.24 | -3.56 | 2.59 | 15.38 | 5.47 | 14.68 | 12.30 | 10.62 | 3.93 | 2.81 | 12.65 | 5.43 | 9.65 | 3.42 | 8.07 | 8.76 | 10.15 |
| Alcohol-related | 8.15 | 15.63 | 5.63 | 14.89 | 17.36 | 15.95 | 19.42 | 13.11 | 14.40 | 11.96 | 12.03 | 12.73 | 9.64 | 7.89 | 14.46 | 11.70 | 15.05 | 6.49 | 15.36 |
| Drug-related | -5.72 | 1.51 | -2.67 | -2.51 | -0.31 | 1.75 | -0.23 | 2.92 | -0.25 | 3.75 | 3.31 | 9.39 | 8.72 | 8.22 | 13.15 | 10.60 | 17.30 | 23.62 | 23.68 |
| **Scotland** |  |  |  |  |  |  |  |  |  |  |  |  |  |  |  |  |  |  |  |
| Malignant neoplasms | -0.31 | -0.12 | -1.65 | 3.25 | 2.84 | 3.30 | -1.69 | 1.87 | 0.79 | -1.49 | 2.23 | -0.53 | 2.89 | 2.05 | 6.79 | 8.64 | 5.50 | 6.17 | 6.19 |
| Circulatory diseases | 17.33 | 11.69 | 16.09 | 16.79 | 17.80 | 19.75 | 17.84 | 16.96 | 13.70 | 17.10 | 16.02 | 16.91 | 15.61 | 14.86 | 17.55 | 14.73 | 18.24 | 12.70 | 13.86 |
| External causes | 7.43 | 7.24 | 4.90 | 10.85 | 1.09 | 12.71 | 6.94 | 14.13 | 4.82 | 3.99 | 37.26 | 41.46 | 39.11 | 32.55 | 38.68 | 55.17 | 60.70 | 76.96 | 89.17 |
| Suicide & deaths of<br>undetermined intent | 2.85 | 7.39 | 6.06 | 6.16 | 1.62 | 7.02 | 3.02 | 8.61 | 4.73 | 5.47 | 4.60 | 3.99 | 9.59 | 0.62 | 5.89 | 6.64 | 9.54 | 10.19 | 12.74 |
| Alcohol-related | 23.62 | 23.92 | 24.29 | 26.70 | 25.03 | 29.99 | 22.01 | 24.07 | 20.48 | 19.88 | 17.21 | 14.37 | 14.70 | 14.05 | 14.89 | 14.26 | 13.13 | 13.11 | 12.53 |
| Drug-related | 16.62 | 23.98 | 18.24 | 21.70 | 19.55 | 24.57 | 27.11 | 36.60 | 30.03 | 32.14 | 33.20 | 35.02 | 32.53 | 37.07 | 37.93 | 51.79 | 54.77 | 69.14 | 73.77 |
| **Wales** |  |  |  |  |  |  |  |  |  |  |  |  |  |  |  |  |  |  |  |
| Malignant neoplasms | -5.21 | -4.37 | 6.30 | -0.96 | 0.71 | 5.35 | 4.57 | -2.40 | 2.58 | -3.50 | -1.49 | -2.67 | 3.32 | 0.70 | 0.52 | 4.14 | 2.94 | 2.56 | 4.90 |
| Circulatory diseases | 8.37 | 7.31 | 5.51 | 11.13 | 9.33 | 13.44 | 10.38 | 10.79 | 10.87 | 8.31 | 12.81 | 10.12 | 10.86 | 8.64 | 12.19 | 16.95 | 15.63 | 17.99 | 14.40 |
| External causes | -5.79 | -9.12 | 0.35 | -8.19 | -8.37 | -3.62 | -1.45 | -3.26 | -4.71 | -4.57 | 9.20 | 14.34 | 19.94 | 4.84 | 26.00 | 30.15 | 23.75 | 31.76 | 19.61 |
| Suicide & deaths of<br>undetermined intent | -1.74 | -3.99 | 1.01 | -1.40 | -6.88 | -0.53 | -1.16 | -1.57 | -3.53 | -2.67 | 1.18 | 2.00 | 8.08 | -5.97 | 3.98 | 2.11 | 6.41 | 3.85 | 7.29 |
| Alcohol-related | 9.38 | 5.48 | 8.08 | 8.98 | 6.86 | 10.85 | 11.22 | 14.87 | 15.13 | 10.81 | 10.96 | 11.11 | 10.81 | 10.05 | 9.40 | 10.79 | 7.56 | 11.37 | 7.91 |
| Drug-related | 7.45 | 8.28 | 6.35 | 6.26 | 6.60 | 4.71 | 8.32 | 5.00 | 10.60 | 15.69 | 14.01 | 13.65 | 14.19 | 10.14 | 20.57 | 23.78 | 19.49 | 21.62 | 13.40 |
| **United Kingdom** |  |  |  |  |  |  |  |  |  |  |  |  |  |  |  |  |  |  |  |
| Malignant neoplasms | -3.63 | -4.39 | -1.85 | -3.43 | -2.16 | -0.20 | -1.64 | -3.02 | -1.21 | -1.53 | -1.50 | -2.39 | 0.56 | -0.43 | 0.75 | 0.92 | 2.28 | 2.75 | 2.07 |
| Circulatory diseases | 5.51 | 5.24 | 6.41 | 7.81 | 7.32 | 7.86 | 6.39 | 7.56 | 7.33 | 8.00 | 6.19 | 7.07 | 6.77 | 6.51 | 6.72 | 7.50 | 7.72 | 9.06 | 7.14 |
| External causes | -17.20 | -15.16 | -13.82 | -12.38 | -13.58 | -8.92 | -12.51 | -6.41 | -11.56 | -13.72 | -5.38 | -2.10 | 1.47 | 3.10 | 7.15 | 6.96 | 8.73 | 15.80 | 19.35 |
| Suicide & deaths of<br>undetermined intent | -8.84 | -8.14 | -6.19 | -7.31 | -8.95 | -5.72 | -7.06 | -4.36 | -5.57 | -6.40 | -5.75 | -5.41 | -2.43 | -6.51 | -2.45 | -3.37 | -1.78 | -0.36 | 2.35 |
| Alcohol-related | 7.01 | 6.64 | 7.67 | 8.07 | 7.08 | 11.21 | 8.26 | 10.42 | 10.05 | 9.94 | 9.49 | 7.80 | 7.43 | 6.65 | 7.98 | 7.13 | 7.02 | 5.91 | 6.50 |
| Drug-related | 4.32 | 6.73 | 4.48 | 5.91 | 5.94 | 4.99 | 4.42 | 7.00 | 6.12 | 7.33 | 4.32 | 5.77 | 8.38 | 10.85 | 12.81 | 13.46 | 13.84 | 16.17 | 17.85 |

## Supplementary Methods

**Supplementary Table SM1.**
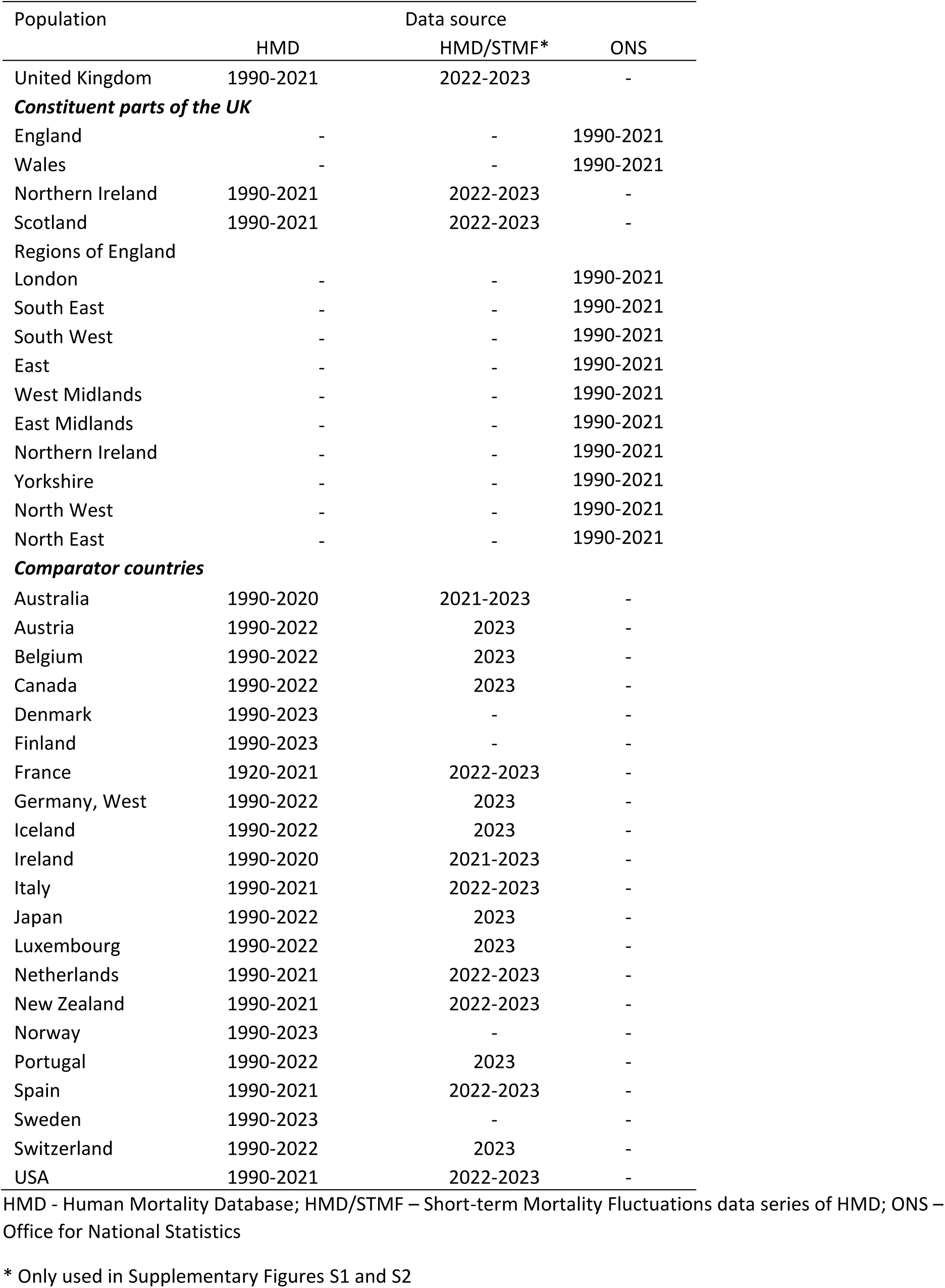
Populations and data sources for all-cause mortality analysis

Supplementary Methods Tables and Figures Supplementary Table SM1. Populations and data sources for all-cause mortality analysis
| Population | Data source |  |  |
| --- | --- | --- | --- |
|  | HMD | HMD/STMF* | ONS |
| United Kingdom | 1990-2021 | 2022-2023 | - |
| ***Constituent parts of the UK*** |  |  |  |
| England | - | - | 1990-2021 |
| Wales | - | - | 1990-2021 |
| Northern Ireland | 1990-2021 | 2022-2023 | - |
| Scotland | 1990-2021 | 2022-2023 | - |
| ***Regions of England*** |  |  |  |
| London | - | - | 1990-2021 |
| South East | - | - | 1990-2021 |
| South West | - | - | 1990-2021 |
| East | - | - | 1990-2021 |
| West Midlands | - | - | 1990-2021 |
| East Midlands | - | - | 1990-2021 |
| Northern Ireland | - | - | 1990-2021 |
| Yorkshire | - | - | 1990-2021 |
| North West | - | - | 1990-2021 |
| North East | - | - | 1990-2021 |
| ***Comparator countries*** |  |  |  |
| Australia | 1990-2020 | 2021-2023 | - |
| Austria | 1990-2022 | 2023 | - |
| Belgium | 1990-2022 | 2023 | - |
| Canada | 1990-2022 | 2023 | - |
| Denmark | 1990-2023 | - | - |
| Finland | 1990-2023 | - | - |
| France | 1920-2021 | 2022-2023 | - |
| Germany, West | 1990-2022 | 2023 | - |
| Iceland | 1990-2022 | 2023 | - |
| Ireland | 1990-2020 | 2021-2023 | - |
| Italy | 1990-2021 | 2022-2023 | - |
| Japan | 1990-2022 | 2023 | - |
| Luxembourg | 1990-2022 | 2023 | - |
| Netherlands | 1990-2021 | 2022-2023 | - |
| New Zealand | 1990-2021 | 2022-2023 | - |
| Norway | 1990-2023 | - | - |
| Portugal | 1990-2022 | 2023 | - |
| Spain | 1990-2021 | 2022-2023 | - |
| Sweden | 1990-2023 | - | - |
| Switzerland | 1990-2022 | 2023 | - |
| USA | 1990-2021 | 2022-2023 | - |

HMD - Human Mortality Database; HMD/STMF – Short-term Mortality Fluctuations data series of HMD; ONS – Office for National Statistics
\* Only used in Supplementary Figures S1 and S2

**Supplementary Table SM2.**
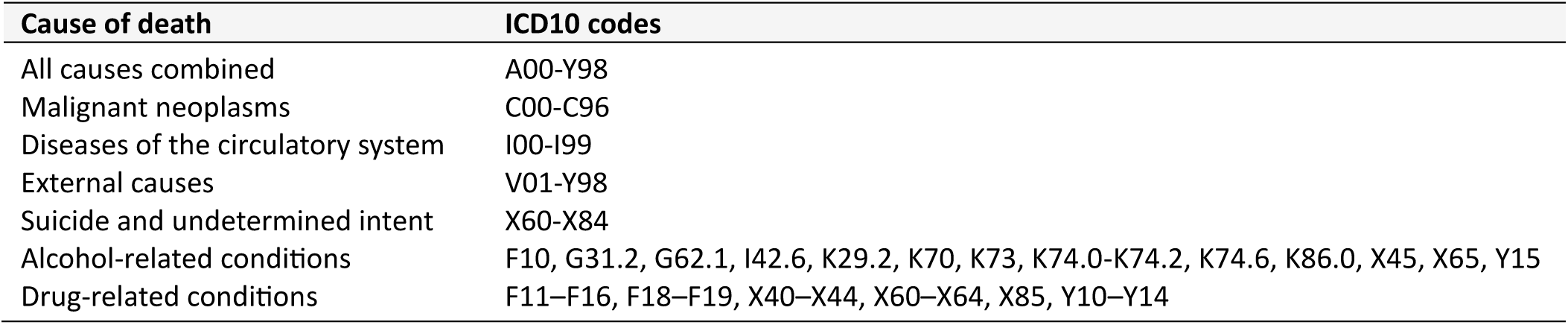
Selection of causes of death for the cause-of-death analysis

**Supplementary Figure SM1.**
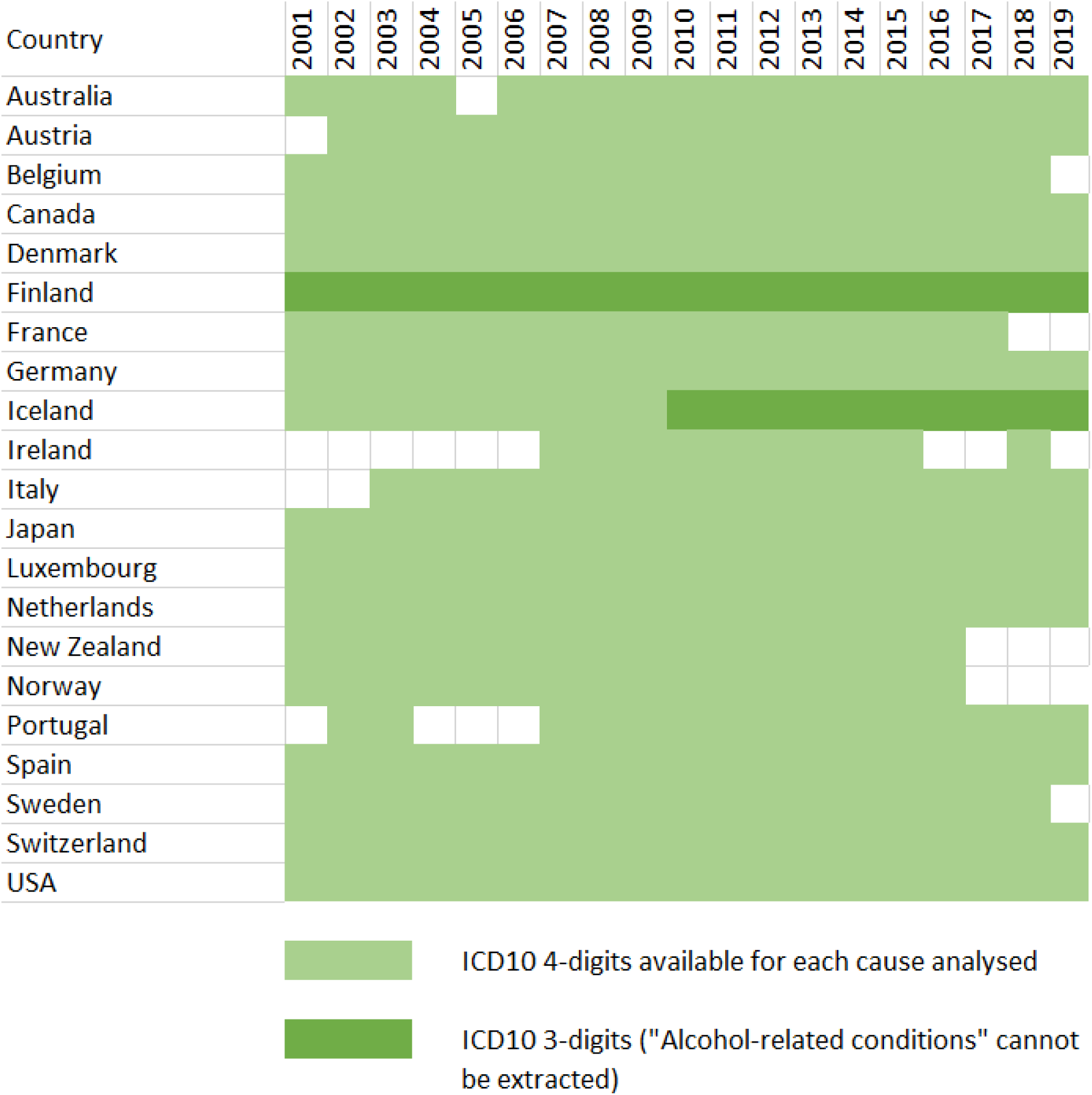
Data availability map for the WHO cause-specific mortality data used in this study

## Supplementary Methods Annex

### 1. Human Mortality Database (HMD)

The HMD data^1^ has unique strengths due to the strict comparability of data across time and space ensured by a comprehensive set of methods for the elimination of distortions caused by population re-enumerations at censuses and changes in data coverage as well as methods for treatment of mortality understatement at ages 80 and older.^2^

### 2. Data sources for England, Wales, Scotland and Northern Ireland

We obtained data on all-cause and cause-specific deaths and populations for England and Wales separately, together with the nine standard regions of England directly from the ONS.^3^ For each sex-year-age combination, the ONS all-cause death and population numbers were adjusted to ensure strict equivalence of the sum of England plus Wales and the sum of nine regions of England plus Wales to the HMD death and population numbers for England and Wales as a whole.

For Scotland and Northern Ireland, all-cause death and population numbers were taken directly from the HMD. For the cause-specific analyses, death and population numbers for Scotland were obtained from the National Records of Scotland (NRS) and for Northern Ireland from the Northern Ireland Statistics and Research Agency (NISRA). These numbers were adjusted for the strict equivalence of their sums to the corresponding HMD numbers.

### 3. Estimation of median values for 21 comparator countries

For the analysis of cause-specific mortality, we utilise different benchmark indicators than for the analysis of all-cause mortality. For all-cause mortality analyses, we use the ASDR of the median country as a comparison, while for cause-specific analyses, we use the median of country-specific ASDRs as a comparison. Details of the calculations and sensitivity analysis are provided below.

#### 3.1 All-cause mortality

First, we create a “median population” using age-specific death rates (25-49 years) of 21 comparator countries:

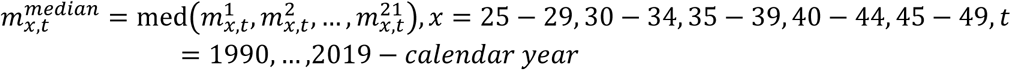

where 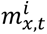 is death rate at age *x* in year *t* in country *i*.

At the next step, we calculate ASDR*^median^* as follows:

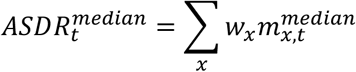

where 𝑤_𝑥_ are age-specific weights of the 2013 European standard population.

#### 3.2 Cause-specific mortality

We estimated the median ASDR (medASDR) as follows:

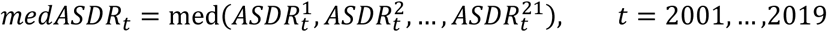

where 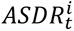 are country-specific age-standardized death rates:

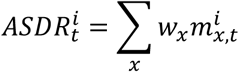

For the broad age group [a,b), the median ASDR is calculated as follows:

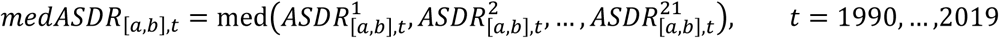

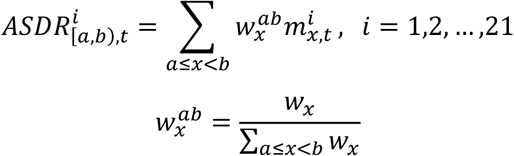

To ensure comparability with all-cause analysis which utilised ASDRs calculated using median age-specific rates, for every sex-year combination, we adjusted the cause-specific medians so that their sum equaled the all-cause median. The adjustment factors varied from 1 to 1.09 being mostly lower than 1.05.

#### 3.3 Sensitivity analysis: ASDR^median^ vs. medASDR

Theoretically, ASDR*^median^* might significantly deviate from medASDR. In our case, the difference between these two indicators is minor. The bar plot in the Figure below shows distribution of relative difference (in %) between these two indicators by age group for the selected 21 countries for the period 1990 to 2019 for males and females.

**Figure:**
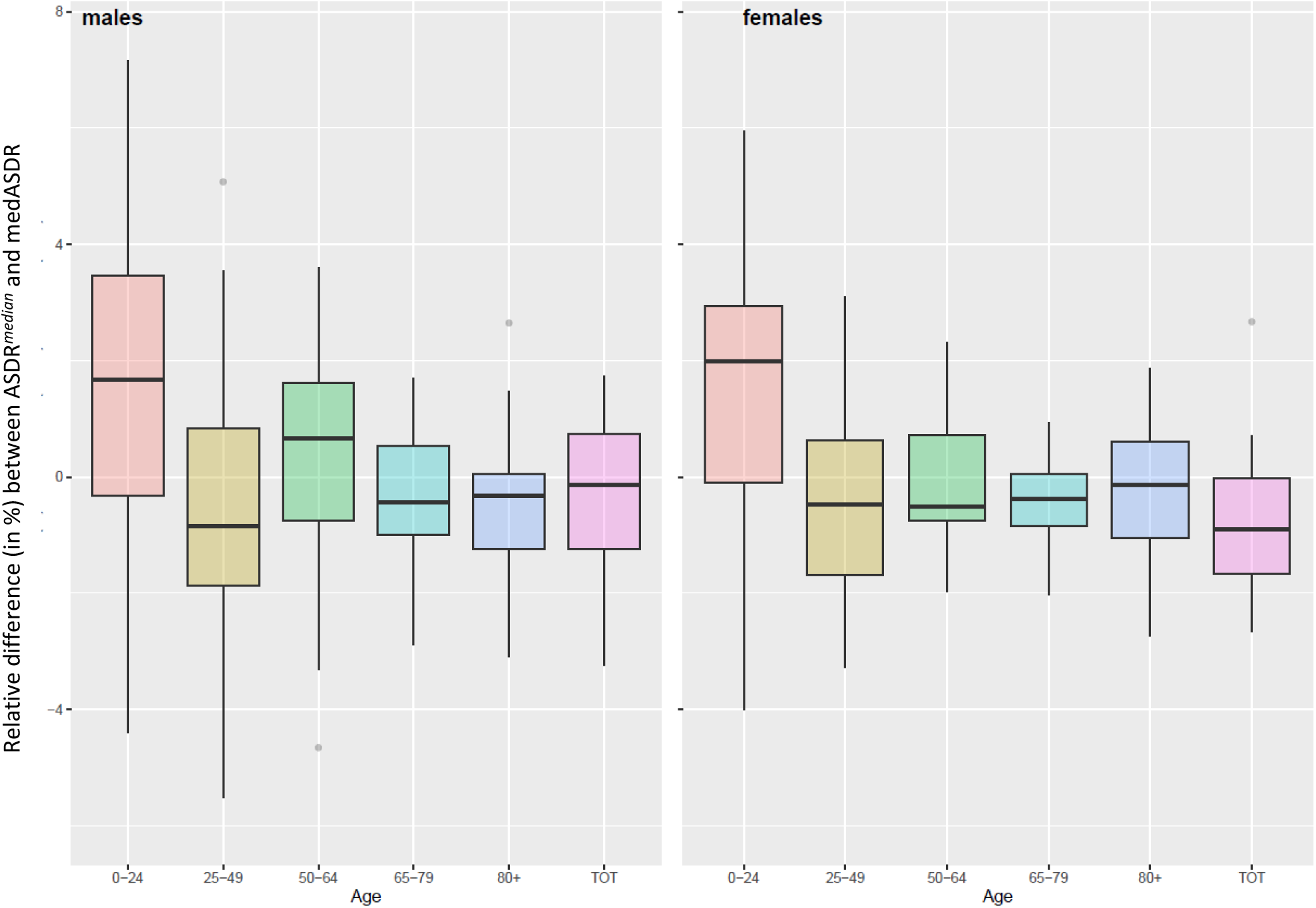
Distribution of relative difference (in %) between ASDR*^median^*and medASDR by age group for the selected 21 countries for the period 1990 to 2019 for males (left panel) and females (right panel).

### 4. Excess deaths

Excess deaths were calculated as the difference between the observed number of deaths and the number of deaths that would occur if mortality were equal to the median rates:

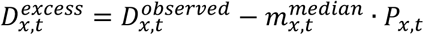

where *x* and *t* denote age group *x* and year *t*, 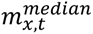 is the 21-country median death rate at age *x* in year *t*, and 𝑃_𝑥,𝑡_ is population of the UK (or respective region) in year *t* and age group *x*.

The excess years of life lost were calculated by multiplying the excess deaths by the WHO standard values of potentially achievable (frontier) life expectancy^4^ :

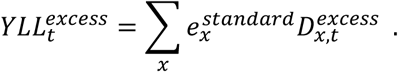

### 5. Coding of cause of death in the UK from 2001 and inconsistencies in cause-of-death series

The principal limitation in comparing cause-specific mortality across time and space is variability in practices of medical diagnostics and approaches to determining the underlying cause of death.^5^ These differences are generally related to variations in medical technologies, healthcare infrastructure, and established medical practice. Significant inconsistencies in the cause of death series may be produced by updates of the 10th revision of the International Classification of Diseases (ICD10).^6^ These updates are modifications of the WHO rules for cause-of-death coding recommended by the WHO for implementation by national statistical agencies during the ICD10 period.

#### 5.1 Automated coding software changes and ICD10 updates

ICD10 was used in the whole of the UK from 2001. It was introduced in 2001 in England and Wales by the Office for National Statistics (ONS) and in Northern Ireland by the Northern Ireland Statistics and Research Agency (NISRA). In Scotland, it was introduced in 2000 by the National Records of Scotland (NRS).

Automated coding with the Mortality Medical Data System (MMDS) software, developed in the U.S., was utilized by ONS and NRS until 2010 to select and code the underlying cause of death using medical death certificates as an input. In Northern Ireland, manual coding was used until 2016. Since then, ONS has performed the automated selection of the underlying cause of death for Northern Ireland on behalf of NISRA.

In 2011, ONS and NRS implemented a new version of the MMDS software to account for the ICD10 updates accumulated by that time. To quantify the impact of the introduction of ICD10 v2010 instead of ICD10 v2001, ONS and NRS completed bridge coding studies in which the same sets of deaths were independently coded using the “old” and the “new” versions of ICD10. The ONS study detected impacts of the ICD10 update such as: a 32% increase in deaths from mental and behavioral disorders, a 5% decrease in deaths from diseases of the circulatory system, a 21% decrease in deaths from diseases of the genitourinary system, and a 2% increase in deaths from diseases of the respiratory system.^7^ The Scottish bridge coding study revealed a 21% increase in deaths from external causes, a 5% decrease in deaths from diseases of the respiratory system, a 14% decrease in deaths from metal and behavioral disorders, and a 16% increase in diseases of the nervous system.^8^

The MMDS was subsequently replaced by the European IRIS Software – an automatic system for coding multiple causes of death and selecting the underlying cause of death.^9^ The transition was made in 2014 for England and Wales^10^ and in 2017 for Scotland.^11^ The new software adopted the official ICD10 updates up to 2013. Iris Software has been used to code Northern Irish data on causes of death since 2016.

The transition to the IRIS software did not produce ruptures as evident as those between 2010 and 2011. However, the bridge-coding studies performed by ONS and NRS indicate that trends in some causes of death were still moderately affected. In particular, both ONS and NRS reported that the software change produced an increase in deaths from mental and behavioral disorders (+7% in England and Wales, +6.2% in Scotland) compensated by a decrease in respiratory diseases (-2.5% in England and Wales, -4.8% in Scotland).

The WHO does not provide information about the implementation of the ICD10 updates by the member states and also about their use of automated coding systems. Therefore, changes in cause-of-death coding and their potential effects on cause-specific mortality in the 21 comparator countries are unknown.

#### 5.2 Specific changes in coding rules

As the previous section showed, distinct inconsistencies in the cause-of-death series were produced by changes in rules and practices of coding within ICD10. For our analyses of mortality at ages 25-49 years,, the most significant of these was related to changes in coding of drug-related deaths in 2011. The change resulted in some of the relevant deaths being transferred from mental and behavioural disorders to accidental poisoning. However, this change has not affected our analyses as we created a consistent category of drug-related deaths that accounts for this change in coding by combining together all relevant categories.

## References

1. Bor J, Stokes AC, Raifman J, et al. Missing Americans: Early death in the United States-1933-2021. PNAS Nexus 2023; 2(6): pgad173.

2. Leon DA, Jdanov DA, Shkolnikov VM. Trends in life expectancy and age-specific mortality in England and Wales, 1970–2016, in comparison with a set of 22 high-income countries: an analysis of vital statistics data. The Lancet Public Health 2019; **4**(11): e575-e82.

3. Stuckler D, Reeves A, Loopstra R, Karanikolos M, McKee M. Austerity and health: the impact in the UK and Europe. Eur J Public Health 2017; 27(suppl_4): 18-21.

4. Camacho C, Webb RT, Bower P, Munford L. Risk factors for deaths of despair in England: An ecological study of local authority mortality data. Soc Sci Med 2024; 342: 116560.

5. Case A, Deaton A. Mortality and morbidity in the 21(st) century. Brookings Pap Econ Act 2017; 2017: 397–476.

6. Dowd JB, Angus C, Zajacova A, Tilstra AM. Comparing trends in mid-life ’deaths of despair’ in the USA, Canada and UK, 2001-2019: is the USA an anomaly? BMJ Open 2023; **13**(8): e069905.

7. Max Planck Institute for Demographic Research (Germany), University of California Berkeley (USA), French Institute for Demographic Studies (France). HMD. Human Mortality Database. http://www.mortality.org (accessed 11/9/2024.

8. Islam N, Jdanov DA, Shkolnikov VM, et al. Effects of covid-19 pandemic on life expectancy and premature mortality in 2020: time series analysis in 37 countries. BMJ 2021; 375: e066768.

9. Office for National Statistics. Death registrations by cause group and population counts by sex, age group, and region, England and Wales: 1981 to 2021. 2023.

10. World Health Organisation. WHO Mortality Database (accessed 9/1/24). 2024.

11. Keyfitz N. Sampling variance of standardized mortality rates. Hum Biol 1966; 38(3): 309–17.

12. Pechholdova M, Camarda CG, Mesle F, Vallin J. Reconstructing Long-Term Coherent Cause-of-Death Series, a Necessary Step for Analyzing Trends. Eur J Popul 2017; 33(5): 629–50.

13. R Core Team. R: A Language and Environment for Statistical Computing, Vienna, Austria; 2023. Available from: https://www.R-project.org/. In: . editor.

14. Minton J, Shaw R, Green MA, Vanderbloemen L, Popham F, McCartney G. Visualising and quantifying ’excess deaths’ in Scotland compared with the rest of the UK and the rest of Western Europe. J Epidemiol Community Health 2017; 71(5): 461–7.

15. Polizzi A, Dowd JB. Working-age mortality is still an important driver of stagnating life expectancy in the United States. Proc Natl Acad Sci U S A 2024; 121(4): e2318276121.

16. Ho JY. Mortality under age 50 accounts for much of the fact that US life expectancy lags that of other high-income countries. Health Aff (Millwood) 2013; 32(3): 459–67.

17. Dowd JB, Doniec K, Zhang L, Tilstra A. US exceptionalism? International trends in midlife mortality. Int J Epidemiol 2024; 53(2).

18. Timonin S, Leon DA, Banks E, Adair T, Canudas-Romo V. Faltering mortality improvements at young-middle ages in high-income English-speaking countries. Int J Epidemiol 2024; 53(5).

19. Raleigh V. Trends in life expectancy in EU and other OECD countries : Why are improvements slowing? . Paris, 2019.

20. Ho JY, Hendi AS. Recent trends in life expectancy across high income countries: retrospective observational study. BMJ 2018; 362: k2562.

21. Harper S, Riddell CA, King NB. Declining Life Expectancy in the United States: Missing the Trees for the Forest. Annual review of public health 2021; 42: 381–403.

22. Shanahan L, Hill SN, Gaydosh LM, et al. Does Despair Really Kill? A Roadmap for an Evidence-Based Answer. Am J Public Health 2019; 109(6): 854–8.

23. McCartney G, McMaster R, Popham F, Dundas R, Walsh D. Is austerity a cause of slower improvements in mortality in high-income countries? A panel analysis. Soc Sci Med 2022; 313: 115397.

24. Hiam L, Harrison D, McKee M, Dorling D. Why is life expectancy in England and Wales ’stalling’? J Epidemiol Community Health 2018.

25. Seaman R, Walsh D, Beatty C, McCartney G, Dundas R. Social security cuts and life expectancy: a longitudinal analysis of local authorities in England, Scotland and Wales. J Epidemiol Community Health 2023; 78(2): 82–7.

26. Vodden A, Holdroyd I, Bentley C, et al. Evaluation of the national governmental efforts between 1997 and 2010 in reducing health inequalities in England. Public Health 2023; 218: 128–35.

27. Ramsay J, Minton J, Fischbacher C, et al. How have changes in death by cause and age group contributed to the recent stalling of life expectancy gains in Scotland? Comparative decomposition analysis of mortality data, 2000-2002 to 2015-2017. BMJ Open 2020; **10**(10): e036529.

28. Toffolutti V, Suhrcke M. Does austerity really kill? Econ Hum Biol 2019; 33: 211–23.

29. Steel N, Bauer-Staeb CMM, Ford JA, et al. Changing life expectancy in European countries 1990–2021: a subanalysis of causes and risk factors from the Global Burden of Disease Study 2021. The Lancet Public Health 2025.

30. Office for National Statistics. Death Certification Reform: A Case Study on the Potential Impact on Mortality Statistics, England and Wales. https://assets.publishing.service.gov.uk/media/5a7c699fe5274a5590059b16/dcp171778_288141.pdf (Last accessed 3/3/25), 2012.

## References

1. Max Planck Institute for Demographic Research (Germany), University of California Berkeley (USA), French Institute for Demographic Studies (France). HMD. Human Mortality Database [Available from: http://www.mortality.org] accessed 11/9/2024.

2. Barbieri M, Wilmoth JR, Shkolnikov VM, et al. Data Resource Profile: The Human Mortality Database (HMD). Int J Epidemiol 2015;44(5):1549–56. doi: 10.1093/ije/dyv105 [published Online First: 2015/06/26]

3. Office for National Statistics. Death registrations by cause group and population counts by sex, age group, and region, England and Wales: 1981 to 2021, 2023.

4. Martinez R, Soliz P, Caixeta R, et al. Reflection on modern methods: years of life lost due to premature mortality-a versatile and comprehensive measure for monitoring non-communicable disease mortality. Int J Epidemiol 2019;48(4):1367–76. doi: 10.1093/ije/dyy254

5. Murray CJ, Dias RH, Kulkarni SC, et al. Improving the comparability of diabetes mortality statistics in the U.S. and Mexico. Diabetes Care 2008;31(3):451–8. doi: 10.2337/dc07-1370 [published Online First: 20071024]

6. World Health Organisation. List of Official ICD-10 Updates [Available from: https://www.who.int/standards/classifications/classification-of-diseases/list-of-official-icd-10-updates] accessed 11/9/24.

7. Office for National Statistics. Results from the ICD–10 v2010 bridge coding study. Statistical Bulletin 1 [Available from file:///C:/Users/encddleo/Downloads/statsbulletinbccorrected14feb2013_tcm77-214005pdf], 2011.

8. National Records of Scotland. Changes to the coding of causes of death between 2010 and 2011. Vital Events – Deaths – Background Information [Available at https://wwwnrscotlandgovuk/files/statistics/vital-events/changes-to-coding-of-causes-of-death-between-2010-2011pdf], 2011.

9. Iris Institute. Iris software [Available from: https://www.bfarm.de/EN/Code-systems/Collaboration-and-projects/Iris-Institute/Iris-software/_node.html] accessed 11/9/24.

10. Office for National Statistics. Impact of the Implementation of IRIS Software for ICD-10 Cause of Death Coding on Mortality Statistics, England and Wales 2014 [Available from: https://www.ons.gov.uk/peoplepopulationandcommunity/birthsdeathsandmarriages/deaths/bulletins/impactoftheimplementationofirissoftwareforicd10causeofdeathcodingonmortalitystatisticsenglandandwales/2014-08-08] accessed 11/9/24.

11. National Records of Scotland. The Impact of the Implementation of IRIS Software for ICD-10 Cause of Death Coding on Mortality Statistics in Scotland [Available at https://www.nrscotland.gov.uk/files/statistics/vital-events/impact-of-implementation-iris-for-icd.pdf], 2017.

